# Multiomic analysis reveals cellular, transcriptomic and epigenetic changes in intestinal pouches of ulcerative colitis patients

**DOI:** 10.1101/2023.11.11.23298309

**Authors:** Yu Zhao, Ran Zhou, Bingqing Xie, Cambrian Y Liu, Martin Kalski, Candace M Cham, Zhiwei Jiang, Jason Koval, Christopher R Weber, David T Rubin, Mitch Sogin, Sean Crosson, Mengjie Chen, Jun Huang, Aretha Fiebig, Sushila Dalal, Eugene B Chang, Anindita Basu, Sebastian Pott

## Abstract

Total proctocolectomy with ileal pouch anal anastomosis is the standard of care for patients with severe ulcerative colitis. We generated a cell-type-resolved transcriptional and epigenetic atlas of ileal pouches using scRNA-seq and scATAC-seq data from paired biopsy samples of the ileal pouch and the ileal segment above the pouch (pre-pouch) from patients (male=4, female=2), and paired biopsies of the terminal ileum and ascending colon from healthy individuals (male=3, female=3) serving as reference. Our study finds previously uncharacterized populations of absorptive and secretory epithelial cells within the pouch but not the pre-pouch. These pouch-specific enterocytes express a subset of colon-specific genes, including *CEACAM5* and *CD24*. However, compared to normal colonocytes, expression of these genes is lower, and these enterocytes also express inflammatory and secretory genes while maintaining expression of some ileal-specific genes. This cell-type-resolved transcriptomic and epigenetic atlas of the ileal pouch establishes a reference for investigating pouch physiology and pathology.

## Introduction

Patients with medically refractory Ulcerative Colitis (UC) often undergo ileal pouch anal anastomosis (IPAA), which removes the colon in its entirety and thus the organ affected by UC^1^. An ileal pouch (pouch) established from the terminal segment of the ileum restores continence and improves quality of life^1^. Establishment of the pouch in UC patients is frequently associated with histological changes of the mucosa involving both the epithelium and immune cells^2,3^, including various degrees of inflammation (neutrophil infiltration) and villus atrophy^3,4^. Furthermore, up to 50% of patients will develop severe inflammation of the pouch (pouchitis) within 1-2 years following surgery^5,6^. In most cases, acute pouchitis can be managed with antibiotic treatments and biologics; however, about 60% of these patients subsequently develop chronic pouchitis^7^. The etiology for pouchitis remains unclear, hampering efforts to develop specific treatment strategies^8^. Notably, histological changes of the mucosa alone are not predictive for developing pouchitis^4,9^.

Transcriptional characterization of newly established ileal pouches for UC patients using bulk RNA-seq revealed a colon-like expression profile in pouch compared to pre-pouch samples^10,11^. However, bulk RNA-seq is unable to provide insights into the cellular composition of the pouch epithelium or detect cell-type-specific gene expression differences between pouch and pre-pouch, and the only single cell RNA-seq study of pouch tissue was limited in scope, focusing only on immune cells (CD45+)^12^.

Therefore, it remains unclear whether the ileum-derived mucosa of the pouch undergoes changes in cell type composition and how gene expression changes in specific cell types after pouch functionalization. Addressing these questions is an important first step to understanding the cellular processes that shape this unique tissue and a necessary basis for studies aiming to identify molecular characteristics predisposing patients to pouchitis.

Here we use single cell RNA-seq (scRNA-seq) and single cell assay for transposase-accessible chromatin with sequencing (scATAC-seq) to generate a cell-type-resolved atlas of the UC pouch mucosa and to obtain transcriptional and epigenetic profiles of component cell types. We find and characterize absorptive and secretory cell populations unique to UC pouch, but not pre-pouch. These pouch-specific enterocytes are in part characterized by the expression of a set of colon-specific genes. However, compared to normal colonocytes, expression of these genes is lower, and these enterocytes also express inflammatory and secretory genes while maintaining expression of some ileal-specific genes. By leveraging published longitudinal bulk RNA-seq data, we find evidence that these cells are present in most UC pouches regardless of the development of pouchitis. Using scATAC-seq we identify regulatory features and transcription factors associated with pouch-specific enterocytes. Together, this cell-type-resolved transcriptomic and epigenetic atlas of the ileal pouch provides a resource for investigating pouch physiology and pathology.

## Results

### Single-cell RNA-seq profiling reveals cellular composition of ileal pouch tissue

We aimed to develop a cellular atlas for ileal pouches in UC patients post-IPAA surgery, focusing on delineating their cell types and comparing these with pre-pouch samples from the same patients and healthy tissue samples from control subjects. To this end, we performed scRNA-seq (10x Genomics) on paired biopsy samples from the pouch and pre-pouch from 6 UC-IPAA patients with long established pouches (mean=17 yrs post-surgery, range=9-31 yrs). Using paired samples from the same patients allowed us to use the pre-pouch regions as ‘internal control’ for each patient with the pre-pouch representing the terminal ileum and therefore the tissue from which the pouch was derived. This design aimed to reduce the impact of external and inter-individual variation. While histopathological evaluation indicated the presence of enteritis in the pouch samples, all individuals presented without clinical symptoms of overt pouchitis, this observation agrees with a previous study assessing UC pouch mucosa^4^. Bulk RNA-seq detected increased expression of colon-specific marker genes in pouch compared to pre-pouch samples^10^, and we therefore included biopsies from the TI and AC of 6 healthy individuals undergoing routine screening as reference for healthy tissues and to assess the nature of cellular changes in the pouch (Fig. 1a, Supplementary Fig. 1a, Supplementary Data 1). After quality-control filtering, we obtained 103,772 cells from all scRNA datasets combined (Fig. 1b).

**Fig. 1.**
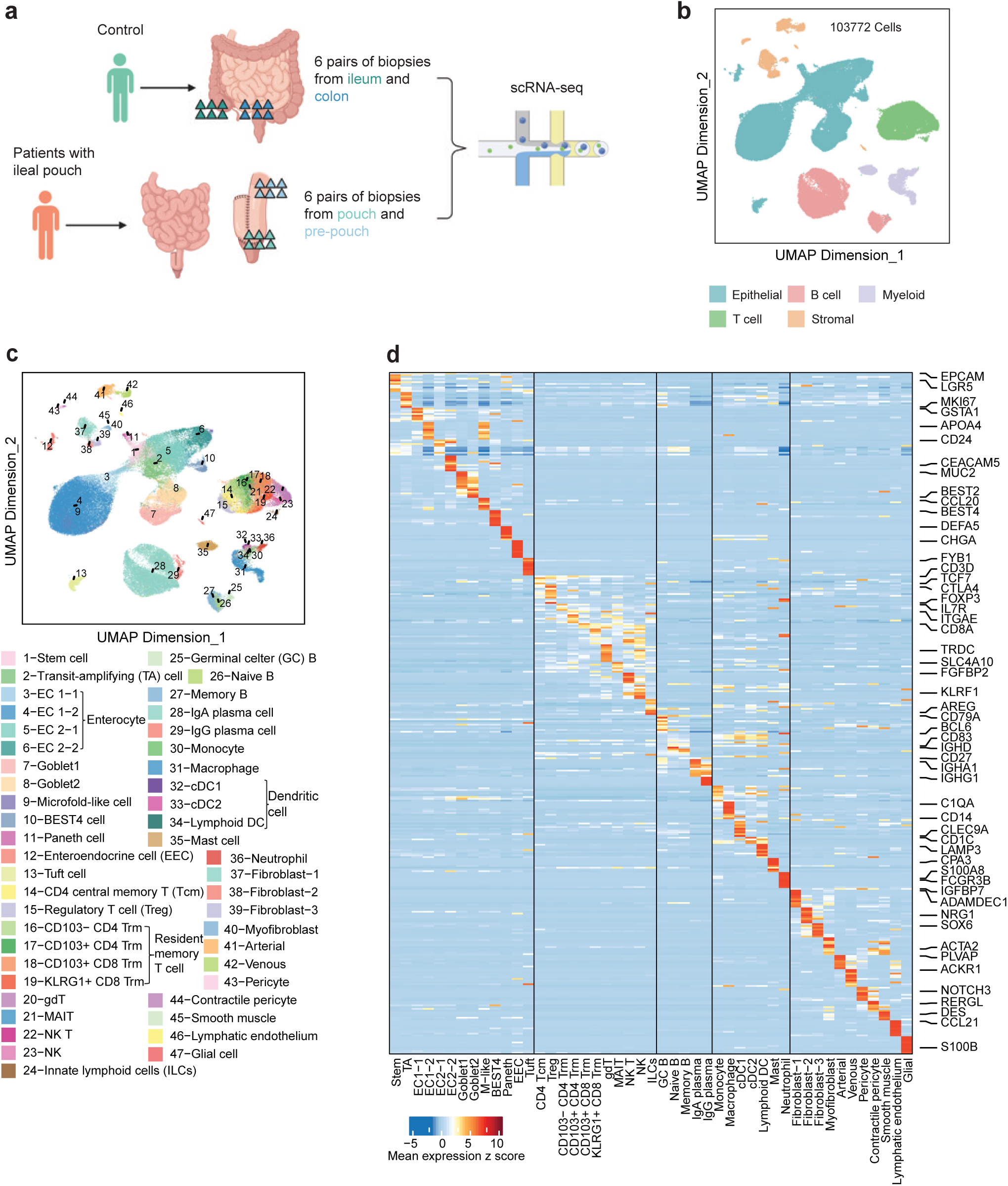
O**v**erview **for the scRNA-seq profiling of 24 human intestinal biopsies from 4 different anatomical regions. a** Schematic of biopsy sampling strategy for scRNA-seq. We profiled 6 pairs of terminal ileum (TI) and ascending colon (AC) biopsies from 6 healthy individuals, and 6 pairs of pre-pouch (PP) and pouch (POU) biopsies from 6 different healthy IPAA patients. Parts of the image were created in BioRender. ZHAO, Y. (2024) https://BioRender.com/b75p140. **b** UMAP dimensionality reduction of 5 major cell lineages in all 24 samples. **c** High resolution UMAP showing 47 cell types in 24 biopsies. **d** Heatmap of top 10 differentially expressed genes and representative cell marker for each cell type within a lineage. Mean expression in each cell type was scaled as z score in each row across all cell types.

Using community-based clustering and marker-gene analysis we identified 5 major clusters comprising cells of the following lineages: epithelial (59,751 cells, marked by *EPCAM*), T-cells (16,389 cells, *CD3D*), B-cells (17,317 cells, *CD79*), myeloid cells (5,375 cells, *TYROBP*), and stromal cells (4,940 cells, *IGFBP7*) (Supplementary Fig. 1c, d). Each individual and sample contributed to all cell lineages (Supplementary Fig. 1b).

We further separated each of the major lineages into multiple cell-types. By combining computational annotation that leveraged previously published data with manual annotation employing well-defined marker genes, we annotated 47 cell types across all lineages (Fig. 1c, d, Supplementary Fig. 2-5, see Methods for detailed description of classification).

We identified all major cell types previously reported for the absorptive and secretory epithelial lineages (Fig. 1c, d, Supplementary Fig. 6, 7)^13^, including stem cells (*LGR5*, *ASCL2*), transit amplifying cells (TA cells) (*MKI67*), absorptive epithelial cells and secretory cells, such as goblet cells (*MUC2*), and Paneth cells (*DEFA5*). Several absorptive and secretory cell populations from the TI and AC clustered separately based on their sampling location. To distinguish these ileal and colonic cell populations we denoted the Enterocyte cell clusters as EC1 (ileal) and EC2 (colon), and similarly identified Goblet1 and Stem1 (ileum) and Goblet2 and Stem2 (colon) clusters. Furthermore, we distinguished progenitor and mature cells within the EC1 and EC2 cell clusters and denoted them as EC1-1 (*SI*, *GSTA2*) and EC2-1 (*CD24*, *CA2*) and EC1-2 (*SLC15A1*, *APOA4*) and EC2-2 (*CEACAM5*), respectively (Fig. 1d).

All samples contained a range of adaptive and innate immune cells representing T-cell, B-cell, and myeloid lineages (Fig. 1c, d, Supplementary Fig. 8-10). T cells were further clustered into CD4, CD8, and gamma-delta T cells. CD4 T cells included central memory (*CCR7*), regulatory (*FOXP3*), CD103- and CD103+ resident memory (CD69+, Supplementary Fig. 3) T cells. CD8 T cells include CD103- and CD103+ resident memory T cells. B cells readily separated into germinal center B cells (*BCL6*), naive B cells (*IGHD*), plasma B cells (*SDC1*) and memory/effector B cells (*CD27*) (Fig. 1c, d). Innate immune cells included macrophages (*CD163*), monocytes (*FCN1*), dendritic cells (*CLEC9A*, *CD1C*), mast cells (*CPA3*), and neutrophils (*FCGR3B*) (Fig. 1c, d). Stromal and other cell types constituted 4.8% (4,940 out of 103,772) of cells. We distinguished several fibroblast populations and identified small clusters of the following cell types: endothelial cells from arteries (*EFNB2*) and veins (*ACKR1*), pericytes (*NOTCH3*), smooth muscle cells (*DES*), lymphatic endothelial cells (*LYVE1*), and glial cells (*S100B*) (Supplementary Fig. 11).

These data constitute the first cellular atlas of UC pouch and pre-pouch tissues, enabling us to compare the cellular and transcriptional changes between pouch and pre-pouch regions and identify similarities and differences to TI and AC reference samples from individuals without IBD history.

### Profound differences in the cellular composition of the epithelial lineages between pouch and pre-pouch

To systematically assess the cellular identities and composition of UC pouches, we used data from all 4 regions (pouch, pre-pouch, AC, TI). We noticed stark differences in cellular composition of the epithelial lineage between pouch and pre-pouch samples, with pouch samples featuring two distinct clusters each of absorptive and secretory cells (Fig. 2). After integration of all samples using Harmony^14^ (Methods), a sizeable proportion of epithelial cells from the pouch clustered with cells from colon samples suggesting some transcriptional similarity. Pouch samples featured two different cell populations for enterocyte, goblet, and stem cells, corresponding to EC1 and EC2, Goblet1 and Goblet2, and Stem1 and Stem2, respectively. (Fig. 2b, c, Supplementary Fig. 12). We observed this compositional change in all 6 individuals, suggesting that partial colonic metaplasia might be a common feature of UC pouches. In contrast, the proportion of these colon-like cell populations was low in pre-pouch samples (Fig. 2c, Supplementary Fig. 12) and their overall cellular composition was similar to TI.

**Fig. 2.**
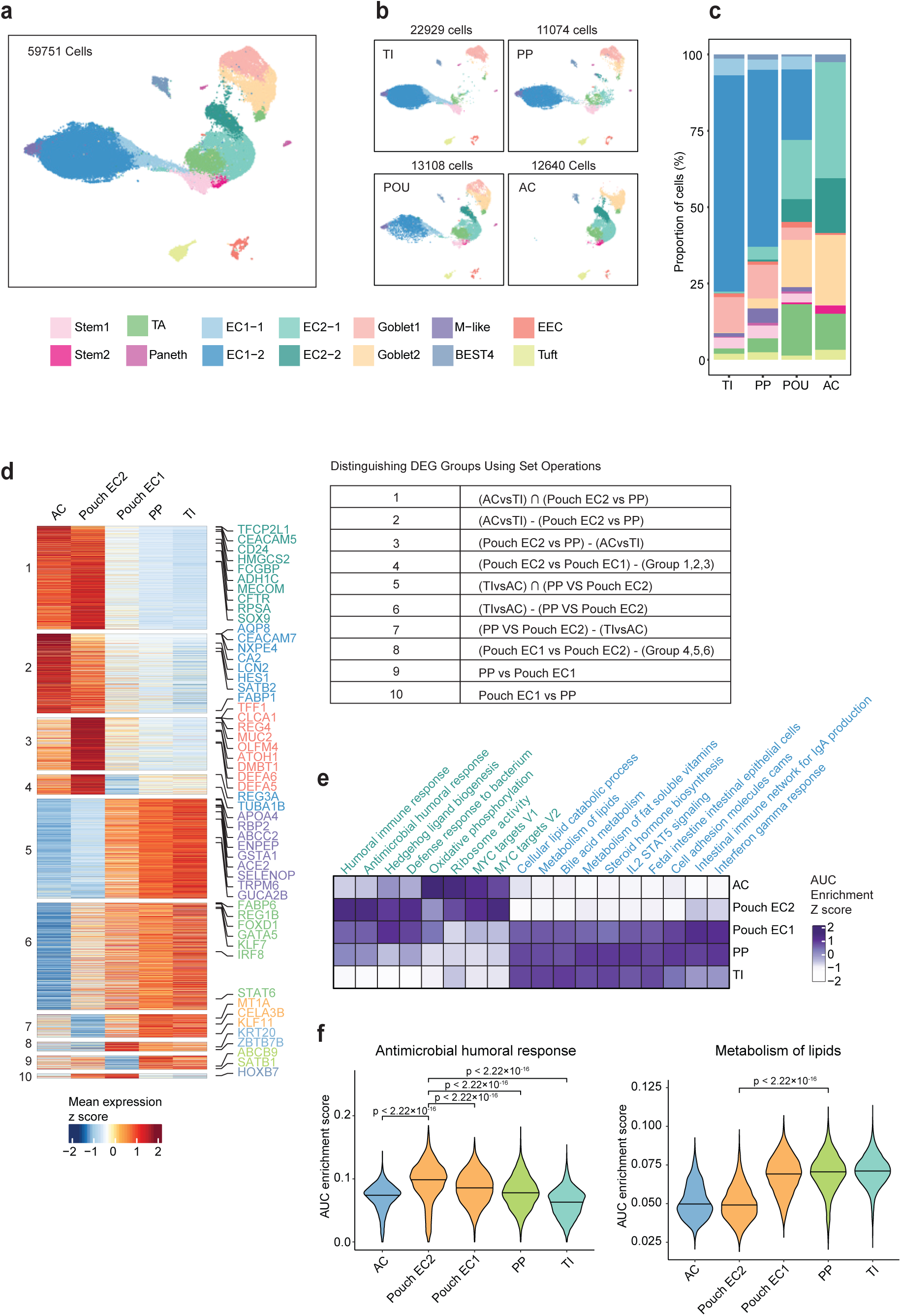
I**d**entification **of both ileum-like enterocytes and colon-like enterocytes in pouch. a** Harmony corrected UMAP of 59,751 cells in epithelial lineage. There are two different populations of enterocytes: ileum-like EC1 and colon-like EC2. EC1-2 and EC2-2 are more mature than EC1-1 and EC2-1 respectively. **b** Harmony corrected UMAP of epithelial cells split based on the four different anatomical regions. **c** Proportions of different cell types in each region. **d** Enterocytes are split into 5 groups: AC EC2, TI EC1, pre-pouch EC1, pouch EC1, and pouch EC2. The DEGs were further grouped into 10 different clusters by the set operations of different contrasts, as summarized in the table. Union of sets (∪) Intersection of sets (∩) Difference of sets (–). **e** Heatmap showing pathway enrichment score in 5 enterocytes groups. The score is scaled across enterocyte groups in each column. **f** AUC enrichment score for antimicrobial humoral response and lipid metabolism related pathway activity. The analysis was conducted between single cells from two cell populations, with each single cell considered a biological replicate. The number of cells in each group is as follows: AC (7,072), pouch EC2 (3,523), pouch EC1 (3,581), PP (7,327), TI (17,633). Statistical significance was assessed using a two-sided Wilcoxon rank-sum test. Notably, all significance tests labeled in the two panels yielded p-values below 2.22×10^−16^, corresponding to the limit of floating-point precision in R. Source data are provided as a Source Data file.

In addition, we observed smaller proportional shifts in other epithelial cell types. The proportion of TA cells across lineages was higher in pouch compared to pre-pouch samples (14% vs 4.8%, p-value=0.0087), and this difference was also observed in AC compared to TI (12% vs 1.9%, p-value=0.0022). The proportion of TA cells in pre-pouch and TI samples differed slightly (4.8% vs 1.9%, p-value=0.041) (Supplementary Fig. 6). The proportion of M-like cells was significantly higher in pouch and pre-pouch compared to AC and elevated compared to TI (Supplementary Fig. 6). We detected Paneth cells in TI, pre-pouch, and pouch but not in AC (Supplementary Fig. 6).

### Pouch-specific changes in non-epithelial cell types

Given the profound cellular changes in the pouch epithelium, we also tested whether similar changes were observed in the immune and stromal compartments. We noted that the changes in the immune compartment were less uniform across patients compared to the epithelial lineage. For example, while we observed an increase in the proportion of IgG plasma B-cells in 3 patients, a change previously observed in UC patients^15^, the proportion of these cells in the remaining patients was indistinguishable from pre-pouch and adjacent tissues. Samples from pouch and pre-pouch contained a slightly higher proportion of CD103+ CD4 Trm cells compared to AC (13%, 11% vs 4.3%, p-value=0.015, 0.0043, respectively), while the proportion of KLRG1+ CD8 Trm was higher in AC, pouch, and pre-pouch compared to TI. Of note, NK T cells were specifically increased in pouch samples compared to pre-pouch samples (1.8%, 0.4%, p-value=0.0087) as were regulatory T cells (13%, 5.1%, p-value=0.0043) (Supplementary Fig. 8). These differences suggest compositional changes of immune cells in pouch samples, possibly reflecting the tissue origin (CD103+ CD4 Trm) and disease status (IgG plasma cells, NK T cells).

Among stromal cell types, we found a significantly higher proportion of the fibroblast-2 (NRG1+) population in the pouch (3.4%) compared to pre-pouch. This population is almost absent in pre-pouch and TI samples (0.2% and 0.3%, respectively) and highest in AC (13%). We complemented these analyses by obtaining spatial transcriptomics data from pouch, pre-pouch, and control samples (Methods). We identified cells expressing NRG1 located underneath the epithelial layer in pouch and AC samples (Supplementary Data 2). In comparison, fewer NRG1 expressing cells were observed in pre-pouch and TI in this location (Supplementary Data 2). These observations thus indicated a remodeling of the stromal niche in UC pouch.

In comparison to the observed changes in the pouch epithelium, the differences in the non-epithelial clusters are smaller and more variable across samples. Increasing the number of cells and samples would improve power to detect differences in the immune and stromal lineages. In the remainder of this study, we focused our analysis on the characterization of the partial metaplasia phenotype observed in epithelial cell types in the pouch.

### Differential expression in pouch enterocytes suggested partial colonic metaplasia

Our clustering analysis suggested the presence of a colon-like enterocyte population in UC pouches but not in pre-pouch regions. However, global similarities between EC2 cells in the pouch and AC based on clustering and embedding following computational integration with Harmony do not directly imply that cells from these regions are identical, and expression of specific genes might differ substantially. We thus compared and characterized the transcriptional programs of enterocytes in the pouch (EC1 and EC2) and compared these differences to expression differences between similar enterocyte populations from pre-pouch and reference samples. We complemented this analysis with a direct comparison of EC2 population in pouch and AC.

First, we detected a substantial number of DEGs between pouch EC2 and EC1 (1,240 genes up in EC2 and 952 genes up in EC1, false discovery rate (FDR) < 0.05, log2 fold-change (logFC) > 1) and between AC (EC2) and TI (EC1) from healthy individuals (1,883 genes up in AC, 2,141 genes up in TI). Notably, a substantial proportion of these DEGs were shared across these two comparisons (836 genes upregulated in pouch EC2 vs pouch EC1 and AC vs TI, and 770 genes upregulated between pouch EC1 vs pouch EC2 and TI vs AC). This analysis suggested that EC1 and EC2 in the pouch recapitulate at least a portion of the expression differences observed between enterocytes from colon and ileum. We also noted lower expression of some typical markers of colonic epithelial cells in pouch EC2 cells (e.g., CA2) (Supplementary Data 3). We further confirmed these common expression differences between EC1 and EC2 cell types through pairwise comparisons across all other conditions and samples (Fig. 2d, Supplementary Fig. 12- 14). Note that the limited numbers of Goblet1 and 2 and Stem1 and 2 cells in the pouch did not allow us to compare these populations directly within the pouch samples. Likewise, assessment of DEGs in EC2 cells in the pre-pouch was not possible due to the exceedingly small number of cells detected per individual. Comparison of aggregate expression profiles of pre-pouch enterocytes suggested similarity with EC1 populations in TI and pouch (Supplementary Fig. 13, 14).

Second, we asked whether the sets of DEGs between populations of EC1 and EC2 cells were enriched for specific cellular functions. DEGs associated with EC2 in the pouch were enriched for immune and antimicrobial pathways and shared enrichment for proliferation associated terms and oxidative phosphorylation with enterocytes from AC (Fig. 2e, Supplementary Data 4). We found that DEGs associated with pouch EC1 populations were strongly enriched in lipid and bile acid metabolism terms (Fig. 2e, f). This comparison suggested that compared to pouch EC1, pouch EC2 cells appear to lack some of the metabolic functions associated with the ileal epithelium while acquiring some functions also enriched in AC enterocytes.

Third, we directly compared pouch to AC EC2 cells and identified 1,442 differentially expressed genes between EC2 cells from these locations (770 up in EC2 pouch, 672 up in AC EC2, FDR < 0.05, logFC > 1) (Supplementary Fig. 12d, Supplementary Data 5). Among the most strongly up- regulated genes in the pouch were genes with known ileal expression and function, including *FABP6*, *APOA4* and *APOB*. In contrast, *AQP8*, *HMGCS2*, *SELENBP1*, *SATB2* and *CA2* showed increased expression in AC EC2 compared to pouch EC2. In addition, among genes upregulated in pouch EC2 compared to AC EC2, we noticed several genes previously reported as part of an expression signature in UC/IBD epithelium, including *REG1A*^16^, *DMBT1*^17,18^, *LYZ*^19^, and *REG4*^20^. This finding indicates that while the global transcriptional identity of pouch EC2 cells resembles that of colonic EC2 cells, they maintain substantial differences in their expression profiles which likely reflect the different tissues of origin, differing microbial and metabolic environments, or disease status.

Among DEGs in EC2 between pouch and AC, we noted higher expression of several IBD-related genes in the pouch. Motivated by this observation, we compared the set of DEGs between AC EC2 and Pouch EC2 to a previous scRNA-seq study of UC samples^21^. We found concordant overlap between a subset of pouch-specific EC2 genes and UC-associated genes from Smillie *et al.*^21^ (81 out of 770 pouch EC2 genes) (Methods). Conversely, AC-specific EC2 genes were enriched for genes down-regulated in colonic epithelium of UC patients in that study (29 out of 672 AC EC2 genes) (Fig. 3a, Supplementary Data 5). This comparison suggested a UC-like inflammatory signature in pouch EC2 cells, highlighting a potentially persistent inflammatory environment. While these markers were differentially expressed between AC EC2 and Pouch EC2 cells from all individuals, it is worthwhile noting that expression of these markers was variable between individuals (Fig. 3a). These expression differences between Pouch and AC samples were not affected by differences in composition of control and UC populations (Supplementary Fig. 15, 16). We also identified similar inflammation-related changes between pouch and AC Goblet2 cells (Fig. 3b, Supplementary Data 5). Of note, a number of DEGs, including LCN2, REG1, DMBT1 were recently associated with distinct epithelial cell populations in active Crohn’s disease^22^.

**Fig. 3.**
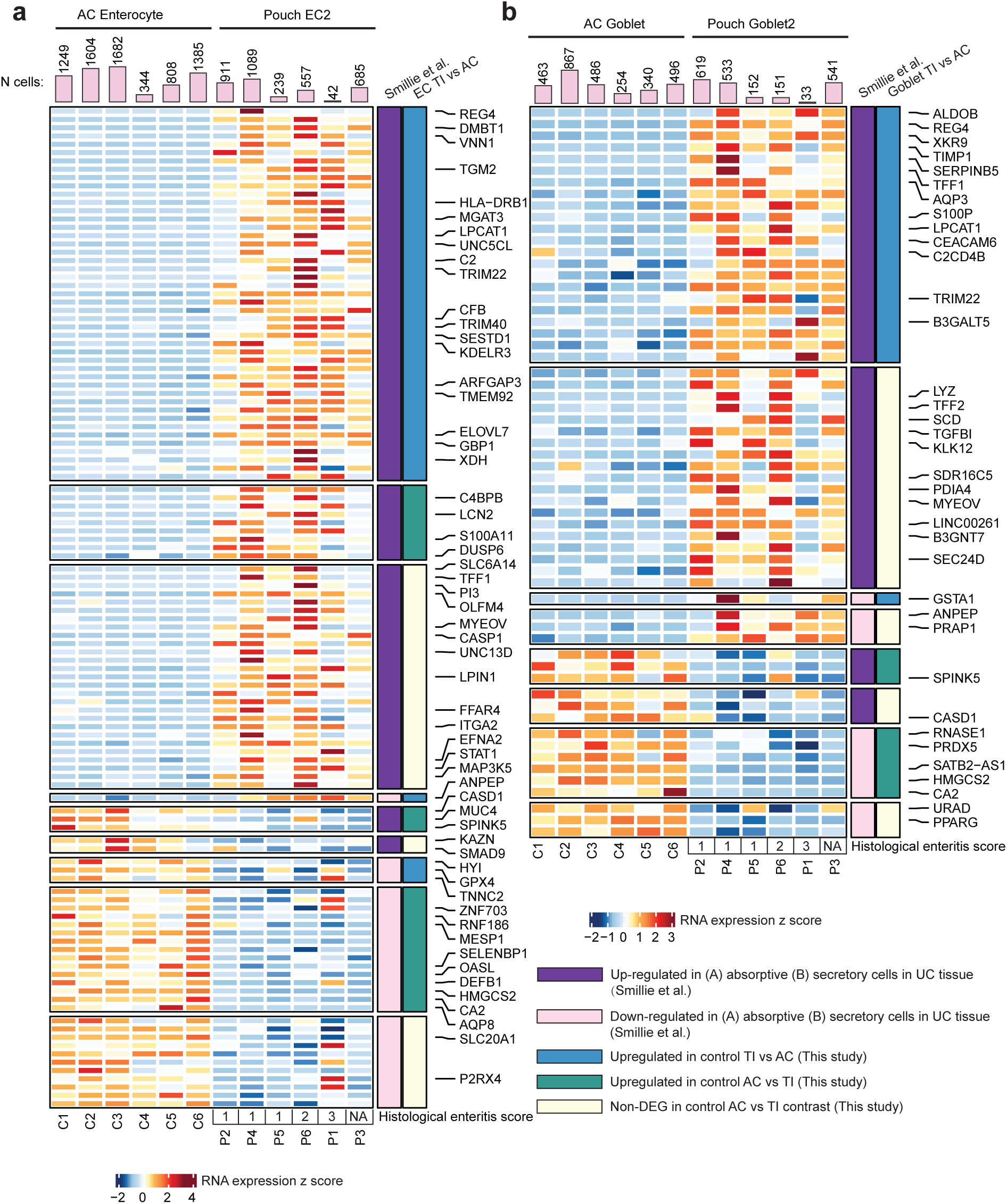
H**e**atmap **showing key differentially expressed genes between cell populations. a** pouch EC2 vs AC EC2. **b** pouch Goblet2 and AC Goblet. Selected genes were reported in a previous study (Smillie et al.)^21^. Pink bars on top are the number of cells in each sample of the corresponding cell type. Expression is scaled in each row across all samples. The last row shows the histological enteritis score of the sample, as also shown in Fig. 4b.

Together, these analyses established the presence of distinct absorptive and secretory enterocyte populations in UC pouches compared to pre-pouches and suggested that the pouch epithelium undergoes partial metaplasia. These comparisons also highlight expression of inflammation-associated genes which were previously identified as part of epithelial IBD signatures suggesting that ongoing inflammation in UC pouches without overt pouchitis affects epithelial programs.

### Histopathology confirmed presence of colon-like cells in pouch epithelium

We sought to validate the metaplastic phenotype and characterize the spatial distribution of EC1 and EC2 cells in the pouch epithelium. We obtained tissue sections from pouch and pre-pouch samples from 5 out of 6 individuals profiled with scRNA-seq, for histopathological assessment and immunohistochemistry (IHC) (Fig. 4a, Supplementary Data 6). In addition to mild to severe enteritis (Fig. 4b), we observed differences of the epithelial architecture in pouch biopsies, including villus atrophy, in agreement with previous reports^3^.

**Fig. 4.**
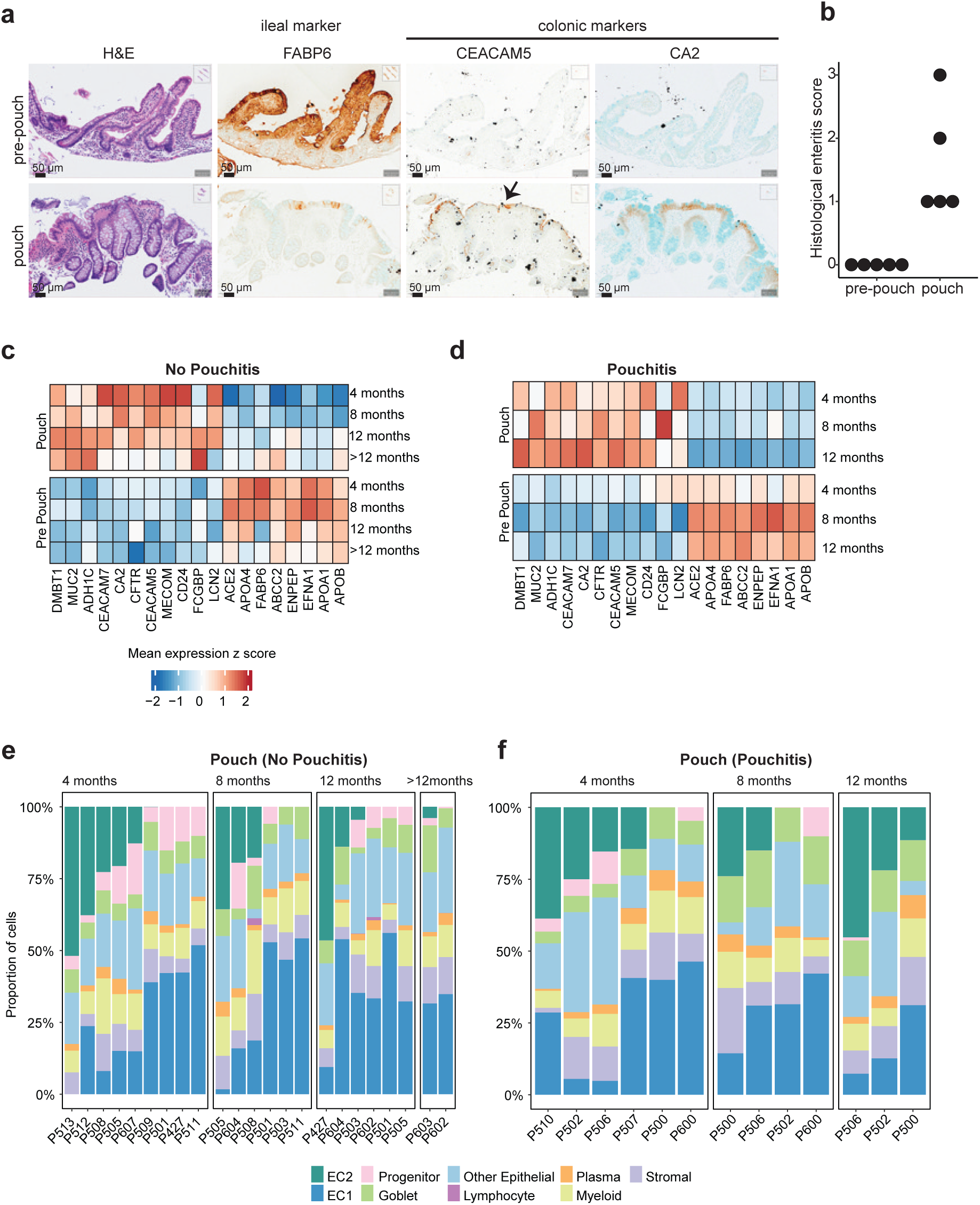
V**a**lidation **of a metaplastic enterocyte population in UC pouches. a** Representative immunohistochemistry images from pre-pouch and pouch biopsies for H&E, FABP6, CEACAM5, CA2 staining. We found evidence of FABP6 or APOA4 expression, small-intestinal markers, in 5/5 pouch samples. 4/5 pouch samples showed expression of CEACAM5 (colonic marker), and 1/5 pouch samples showed expression of CA2 (colonic marker). Experiments to titrate antibodies and validate specificity of labeling were performed on ileal or ascending colonic biopsies collected from 2 patients (Supplementary Data 6). **b** Histological enteritis score of pre-pouch and pouch samples. **c, d** Average gene expression in the bulk RNA-seq data from healthy pouches (**c**) patients and in pouches from patients who developed pouchitis (**d**). Expression for each gene is scaled across pre-pouch and pouch. **e, f** Cell type proportions predicted from bulk RNA-seq data in healthy pouches (**e**) and in pouches from patients who developed pouchitis (**f**). Source data are provided as a Source Data file.

Assessment of sections from pre-pouch biopsies showed staining for the ileal enterocyte marker FABP6 and complete absence of colonic markers CEACAM5 and CA2 (Fig. 4a, Supplementary Data 6). In contrast, expression of FABP6 was greatly diminished in pouch samples and confined to a small subset of cells. In addition, we observed expression of CEACAM5 and CA2 in a subset of epithelial cells from pouch samples (Fig. 4a). CEACAM5 was consistently detected in pouch samples on RNA and protein level, CA2 was only detected in some individuals. These observations validated our scRNA-seq findings by confirming the expression of colonic markers in a subset of epithelial cells, while showing diminished expression of ileal marker genes.

In addition, we performed spatial transcriptomics on sections from two pairs of pre-pouch and pouch samples and controls using the Xenium platform from 10x Genomics targeting 422 genes (Methods). To assess differences between pouch and pre-pouch samples, we delineated spatial domains through the integration of spatial transcriptomic data with our single cell reference dataset using IRIS^23^ (Methods). This domain-level analysis identified two domains enriched for mature enterocytes and revealed an upregulation of EC2-associated genes in pouch samples compared to pre pouch samples within these epithelial domains (Supplementary Data 7). These data also detected strong expression of inflammation-associated markers (e.g. *LCN2, DMBT1*), within the pouch. At the level of individual cells we detected expression of EC1 and EC2 markers in epithelial cells and in agreement with the domain analysis, several EC2 markers, including CEACAM5 and CD24 appeared more highly expressed in pouch samples (Supplementary Data 7). We detected the expression of ileal marker genes was detected across pouch epithelial cells (e.g. APOA4, FABP6) albeit at lower levels than in pre-pouch, which confirms the scRNA-seq findings.

### Pouch metaplasia occurs early after functionalization and independently of subsequent inflammation

We observed colon-like EC2 cells in all our pouch samples; however, our cross-sectional study was limited to six patients with well-established pouches and without clinical symptoms. To generalize our observation, we took advantage of a previously generated bulk RNA-seq dataset^10^ which reported a colon-like expression profile in pouch samples. These data allowed us to validate the expression of EC2 markers in pouch samples, to address when EC2 cells appear after pouch functionalization, and to ask whether their presence is associated with to the development of pouchitis. This longitudinal study included 19 patients with newly established pouches, sampled at three time-points within the first year after UC IPAA surgery (4-mo, 8-mo, 12-mo post-functionalization); 6 out of the 19 individuals were subsequently clinically diagnosed with pouchitis^10^.

We first assessed the expression of EC1- and EC2-specific markers obtained with our scRNA-seq data in bulk samples from pre-pouch and pouch regions. EC2 markers showed increased expression in bulk samples from pouches, while EC1 markers were increased in pre-pouch samples (Fig. 4c, d, Supplementary Data 6). We then used the cell types identified in UC pouch individuals (from pouch and pre-pouch samples) with our scRNA-seq data as reference to deconvolve the bulk RNA-seq data from Huang *et al.* using MuSiC^24^ (Methods). This approach allowed us to infer the constituent cell types and estimate their respective proportions in each bulk sample. We found that the presence of EC2 cells was predicted for the majority of pouch samples (58%, 21/36) (Fig. 4e, f) and in only 3 out of 35 (9%) pre-pouch samples (Supplementary Data 6). We detected EC2 cells (>1%) in 14 out of 19 individuals in at least one time point. The fraction of EC2 cells in pouch samples (4.0% - 51.9%) was generally inversely correlated to the fraction of EC1 cells (Spearman correlation coefficient = -0.82, p-value = 7.4e-10) in the same sample.

EC2 cells were detected in pouch samples at the earliest time point (4-mo) and there was no significant difference in the proportion of samples or the proportion of cells per sample across timepoints (Fig. 4e, f). Similarly, the EC2 cell proportional difference between samples from healthy individuals and those diagnosed with pouchitis is not significant (Mann-Whitney test, p-value = 0.3775) (Fig. 4e, f), suggesting that the presence of EC2 cells alone is not causing pouchitis. Together these data suggest the presence of colon-like EC2 enterocytes in most pouches, providing a cellular basis for the previously reported colon-like expression within pouch samples.

### Integrative scATAC-seq profiling in ileal pouch and neighboring tissues

To identify key transcription factors, gene regulatory elements, and target genes associated with pouch metaplasia, we measured chromatin accessibility using scATAC-seq^25^. To minimize biological and technical variation, scATAC-seq was performed on cells from the same 24 biopsies used for scRNA-seq (Methods).

After quality-control filtering, we retained 88,595 cells, (sample mean = 3,691, range: 1,704- 5,524). Using dimensionality reduction and clustering following previously established procedures, including correcting effects from individuals^26^, we identified cells from all major lineages (Epithelium, T cells, B cells, Myeloid, and Stromal/others) (Supplementary Fig. 17-21). We assigned cell-type labels to each cell by computationally transferring cluster labels from scRNA-seq and observed that with few exceptions, specific cell-type labels mapped to discrete locations in the embedding, suggesting good correspondence between the modalities (Fig. 5a, Methods). To assess the accuracy of our cell-type annotations, we used gene activity scores as a proxy for gene expression^26^ (Methods) and compared the scores for marker genes identified by scRNA-seq across clusters. This comparison further supported our assignment (Supplementary Fig. 22a). Finally, we confirmed that cell-type proportions identified separately by each modality (scRNA-seq and scATAC-seq) were similar for each sample (Supplementary Fig. 22b).

**Fig. 5.**
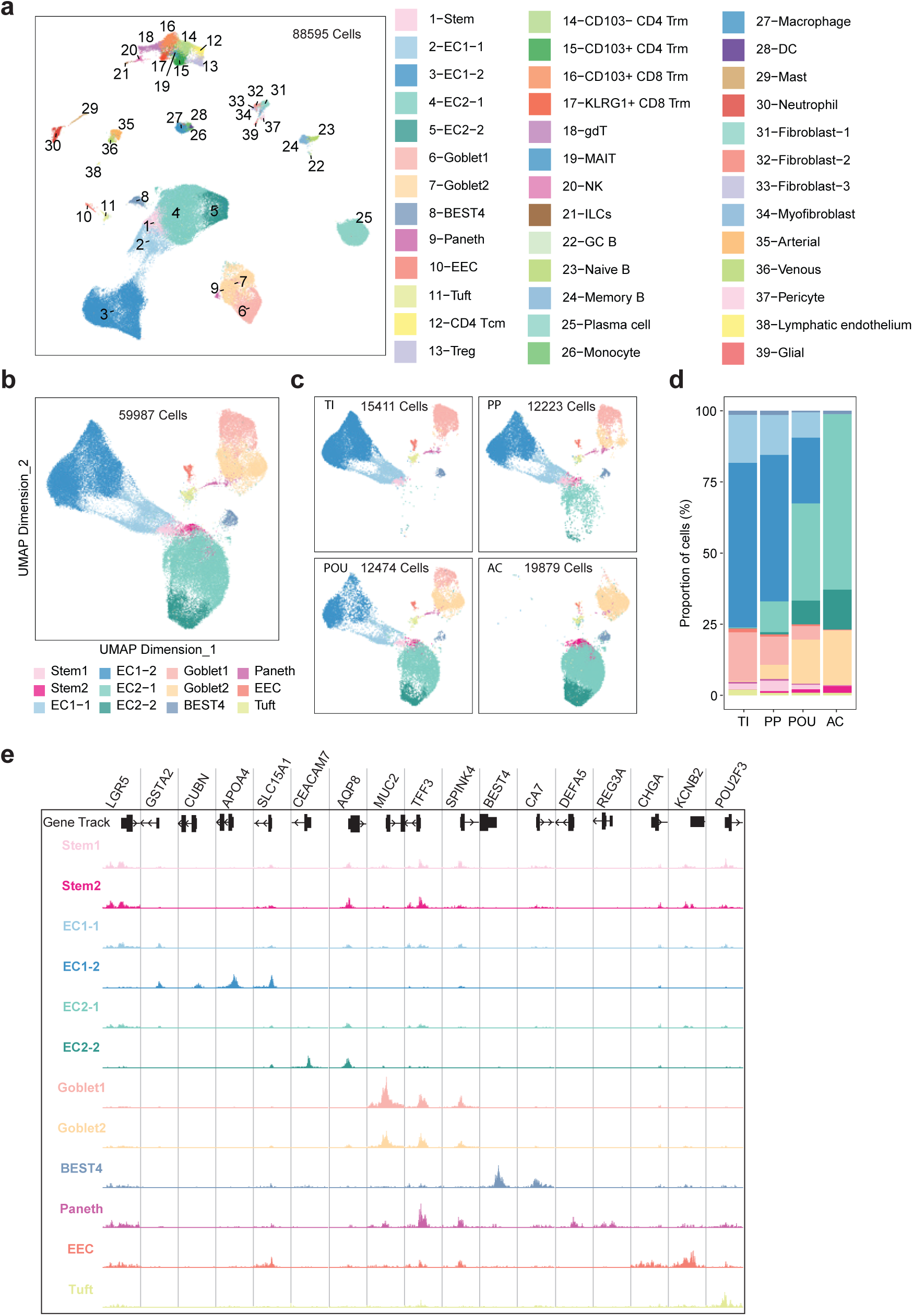
s**c**ATAC**-seq profiling of the same group of 24 biopsies studied in scRNA-seq. a** Harmony corrected UMAP and clustering based on single-cell chromatin accessibility identifies 39 distinct cell types from 88,595 cells across all lineages. **b** Harmony corrected UMAP of the epithelial lineages includes all cell types identified in scRNA-seq analysis. **c** Epithelial UMAP split based on four sampling locations. **d** Proportions of cell types in each region. **e** Normalized accessibility in promoter regions of cell- type-specific marker genes. Source data are provided as a Source Data file.

Focusing on cells from the epithelial lineage alone, we retained 59,987 cells across all samples, including 12,474 cells from pouch samples (Fig. 5b-d). Similar to scRNA-seq, we detected absorptive cells (EC1-2, EC2-2, BEST4), secretory cells (Goblet1, Goblet2, Paneth, EEC, Tuft) and progenitor cells (Stem, EC1-1, EC2-1). We excluded TA cells and M-cells from annotation since these were more ambiguous, likely due to differences between gene scores and RNA expression for informative markers of these cell types (Methods). Loci harboring marker genes for the annotated cell types showed highly specific chromatin accessibility (Fig. 5e).

We confirmed the presence of both ileum-like (EC1) and colon-like enterocyte (EC2) populations in the pouch by comparing clusters from different sampling locations (Fig. 5c). As observed with scRNA-seq, the proportion of EC2 cells was significantly increased in pouch compared to pre-pouch (42.47% vs 11.6%) (Fig. 5d, Supplementary Fig. 17b). These data confirmed the presence of EC2 cells in the pouch using an independent, chromatin-based assay and thus providing the basis to identify and characterize regulatory regions and transcription factors associated with colonic metaplasia in UC pouches.

### Identification of regulatory regions and sequence motifs associated with colon-like cells in the pouch

To identify cis-regulatory elements (CREs) in each epithelial cell type, we identified significantly accessible regions (peaks) in each cell-type cluster separately (Methods). We identified a union set of 262,177 peaks across all clusters with the number of peaks per cluster ranging from 37,127 to 150,000 (Supplementary Fig. 17d). While most peaks were shared across multiple cell types, we observed significant variation in chromatin accessibility across cell types (Fig. 6a). Enrichment analysis within cell-type-specific peaks identified motifs of known master transcription factors (TFs) for these cell types and lineages (Fig. 6b). For example, motifs for HNF4A/G were strongly enriched in ileal-like EC1 cells, while motifs for CDX1/2 were most strongly enriched in colon-like EC2 and progenitor cells (Fig. 6b). This analysis established the union set of candidate CREs in the epithelial lineage and their use across cell types and helped infer sequence motifs associated with cell-type-specific TFs.

**Fig. 6.**
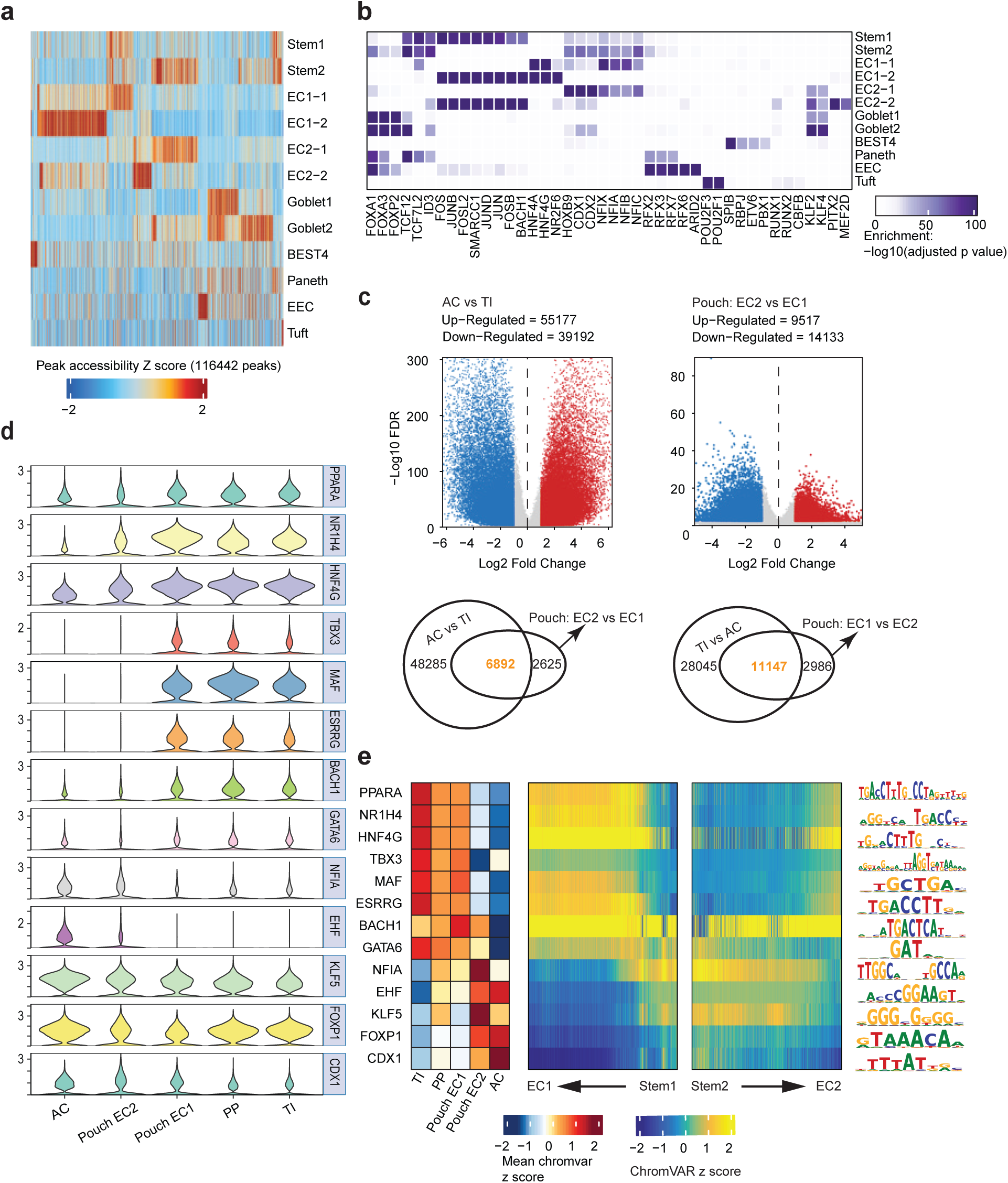
I**d**entification **of marker peaks and transcription factors (TFs) in each cell type in epithelial lineage**. **a** Marker peaks for each cell type in epithelial lineage. Accessibility is scaled in each column across cell types. **b** Transcription factors enriched in each cell type. Enrichment was assessed using the hypergeometric test (one-sided), with p-values adjusted using the Bonferroni correction to account for multiple comparisons. **c** Differentially accessible regions in AC EC2 vs TI EC1 and pouch EC2 vs pouch EC1. **d** Stacked violin plot shows the RNA expression of different TFs in each enterocyte group. **e** Heatmap shows the scaled mean ChromVar TF activity score in each enterocyte group. The score is scaled in each row across enterocyte groups (left). TF activity in EC1 lineage and EC2 lineage along inferred differentiation pseudotime (middle, right).

We then used these candidate CREs to identify putative regulators of EC2 cells in the pouch. Analogous to our identification of differentially expressed genes between EC1 and EC2 cells (Fig. 2d), we performed pairwise comparisons between EC1 and EC2 cell populations from pouch, pre-pouch, TI, and AC (Methods). We identified 120,569 differentially accessible regions (DARs) in at least one comparison (FDR < 0.01, logFC > 1) (Fig. 6c, Supplementary Fig. 23a) and 23,650 DARs between EC1 and EC2 cells in the pouch (14,133 up in EC1, 9,517 up in EC2). In contrast to scRNA-seq, we found substantially more DARs between AC and TI compared to Pouch EC1 and EC2. Nevertheless, this observation provided evidence for epigenetic differences between the two enterocyte cell types in the pouch and suggested plasticity within the epithelial lineages. Of note, 6,892 (72.4%) DARs in pouch EC2 vs EC1 populations were also identified as DARs when comparing EC2 and EC1 cells in AC and TI (Fig. 6c, Supplementary Fig. 23a). Similarly, the comparison of EC1 and EC2 cells in the pouch yielded 11,147 (78.9%) DARs also found when comparing TI to AC enterocytes (Fig. 6c, Supplementary Fig. 23a). This suggests that differences between EC2 and EC1 cells in the pouch were indeed associated with regulatory regions that broadly differentiated AC from TI. However, we also detected 36,848 DARs in total between EC2 cells in the pouch and AC mirroring the results of our DEG analysis and suggested substantial differences in chromatin accessibility between EC2 cells from these two tissues (Supplementary Fig. 5d, Supplementary Fig. 23a).

These sets of DARs were enriched for a range of motifs for TFs involved in epithelial differentiation as well as regulation of inflammatory processes (Supplementary Fig. 23b). To identify candidate transcription factors associated with these enriched motifs, we computed the correlation between integrated RNA transcript levels and motif accessibility within the epithelial lineage (Methods) (Supplementary Fig. 23c) and retained TF-motif pairs with a correlation > 0.5 (activating TF), and correlation < -0.5 (repressive TF)^27^. Expression (Fig. 6d, Supplementary Fig. 23d) and motif accessibility (Fig. 6e, Supplementary Fig. 23d, e) reflected differential activity in EC1 and EC2 cell types for most TFs.

Among the identified TFs, CDX1 and 2, NFIA, EHF, and KLF5 showed higher expression and motif accessibility in EC2 cells from both Pouch and AC (Fig. 6d, e). HNF4G, PPARA, GATA6, JUND were among the TFs associated with EC1 cells in the TI, pouch, and pre-pouch. Several of these TFs we identified in EC1 and EC2 cells in UC pouches, were previously identified in a multimodal single-cell atlas of the human intestine as having preferential activity in the ileum and colon, respectively^28^. We identified these TFs based on transcript levels and correlated enrichment of their annotated motifs within accessible chromatin. We confirmed that these motifs are occupied in epithelial cells clusters by performing TF footprint analysis (Methods, Supplementary Fig. 24). Finally, our spatial transcriptomics data provided additional evidence for expression of these TFs within epithelial cells in pouch and pre-pouch samples (Supplementary Data 8).

By comparing accessibility of these TF motifs along pseudotime-based differentiation trajectories from stem cells into EC1 and EC2 cells, respectively, we observed cell-type-specific activation pattern for these TFs during epithelial differentiation (Fig. 6e, Supplementary Fig. 25, Supplementary Data 9).

Notably, TF expression and motif enrichment appeared to be more heterogeneous in pouch EC2 cells compared to AC EC2 cells. Pouch EC2 cells also maintained activity of several TFs associated with EC1 cells, including BACH1 (Fig. 6e). Motif enrichment and expression of NFIA was more strongly associated with pouch EC2 highlighting differences in TF activity compared to AC. NFIA was identified in progenitor cells of early colonocytes in our data and by Hickey *et al*^28^. In contrast to AC EC2, NFIA motif activity remained high in pouch EC2 cells, possibly reflecting their less mature state.

Together, these TFs represent a key part of the gene regulatory network associated with the two transcriptionally distinct enterocyte populations observed in the pouch epithelium and provide evidence for the activation of a colon-like regulatory network in pouch EC2 cells.

### Activation of a colon-like GRN underlies transcriptional identity of EC2 cells in the pouch

To identify putative target genes of identified cell-type-specific TFs in the pouch epithelium and characterize the regulatory network defining EC2 cells, we linked CREs to genes using co-accessibility of peaks with integrated expression^26^ (Methods). We linked 31,808 peaks to 6,312 genes across EC1 and EC2 populations, identifying a large set of putative targets (Fig. 7a). We observed a strong enrichment of DARs associated with DEG compared to non-DEGs (Supplementary Fig. 26). For example, distal regulatory elements linked to the colon-specific marker gene CA2 gained chromatin accessibility in pouch EC2 cells (Fig. 7b). In contrast, accessibility at CREs linked to ileal marker gene APOA4 was lower in pouch EC2 cells compared to EC1 cells in pouch, pre-pouch and TI (Supplementary Fig. 27).

**Fig. 7.**
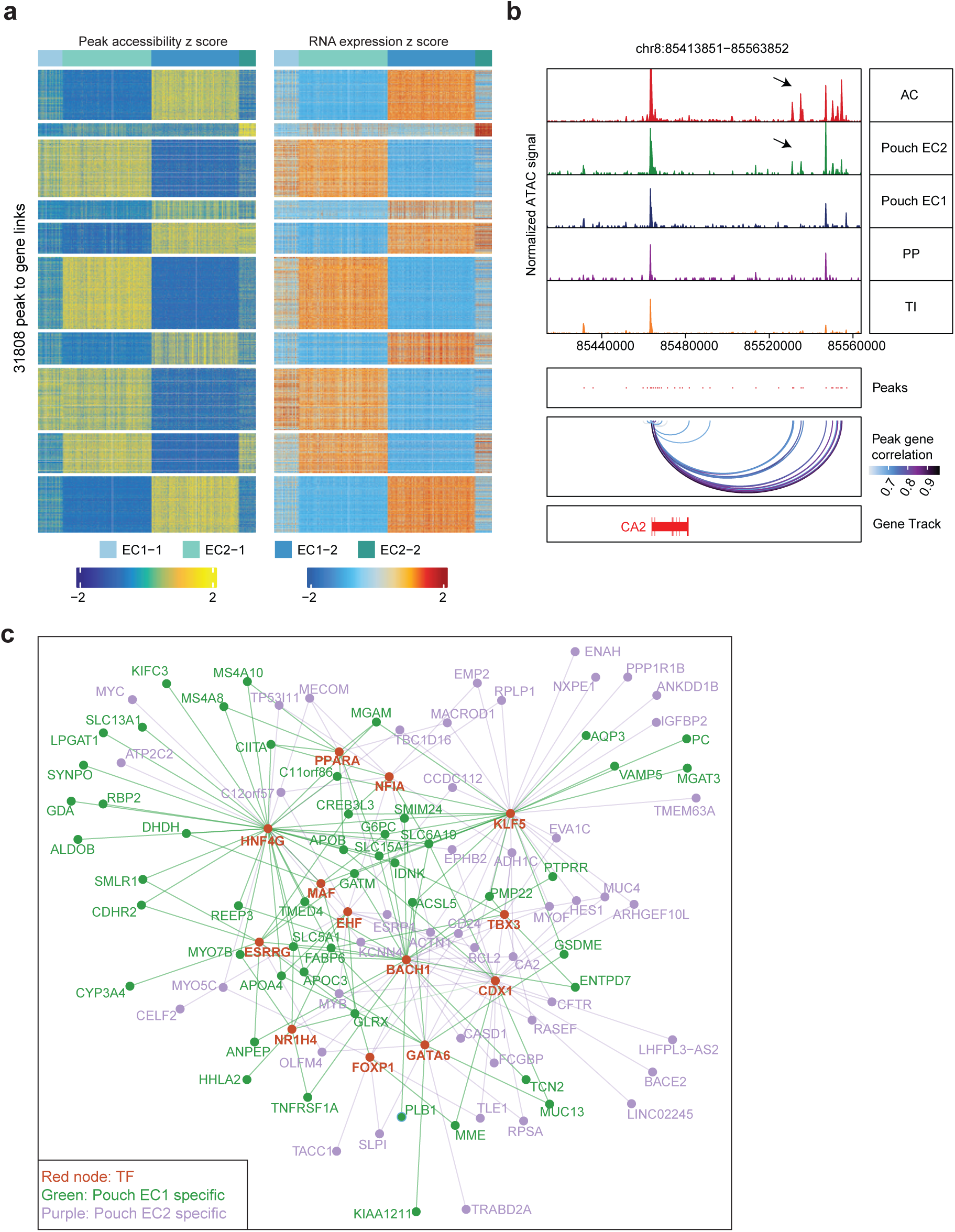
C**o**nstruction **of TF-gene regulatory network in pouch enterocytes by peak-gene linkages based on multiomic analysis. a** 31,808 peak-gene links are identified based on the correlation of gene expression and peak accessibility with correlation score cutoff>0.6. **b** Peaks highly correlated with CA2 gene expression show higher accessibility in colon and pouch EC2 enterocytes (highlighted by arrows). **c** TF-gene regulatory network shows that the TFs (red nodes) are dominating different groups of genes in pouch EC1 and EC2.

We complemented this identification of target genes using FigR to identify domains of regulatory chromatin (DORCs)^29^ which might harbor key target genes and facilitate the identification of regulatory factors. Using this metric, we ranked the genes by the number of associated peaks and identified 765 DORC-associated genes within the epithelial lineage. These putative target genes included both regulatory genes (e.g. ETS2, GATA5, KLF6) and genes associated with cellular function (e.g. OLFM4, MUC2, RBP2) (Supplementary Data 10).

To summarize the regulatory mechanisms underlying EC1 and EC2 cells, we constructed a partial gene regulatory network by collating information from motif enrichment, TF expression, co-accessibility links, DORC scores, and differential gene expression (Methods). This regulatory network highlighted that the DEGs in the two pouch enterocytes populations are targeted by groups of overlapping TFs with differing activities in EC1 and EC2 cells (Fig. 7c). Our data therefore suggested that the colon-like transcriptional program in pouch EC2 cells is the result of the increased activation of TFs predominantly associated with the colon. For example, genes associated with colon function (CA2, CD24, CFTR) are induced in pouch EC2 cells compared to EC1 (Supplementary Fig. 13b). While we showed that this cell type shares regulatory features (CREs and TFs) with enterocytes from the colon and undergoes similar differentiation dynamics, we note that compared to enterocytes from the ascending colon the acquisition of a colonic transcriptional and epigenomic features is incomplete. In addition, we identified several genes associated with ileal function that are still expressed at significant levels in EC2 cells in the pouch compared to colon (e.g., *FABP6*, *APOA4*, *APOB*) (Supplementary Fig. 12d).

## Discussion

To our knowledge, this study is the first single-cell atlas for ileal pouch mucosa in UC patients post-IPAA surgery, leveraging a single-cell multiomic approach. Histological changes associated with UC pouch mucosa are well-documented^3,4^, but the molecular basis of the cellular changes remains unclear^3^. Our study contributed to the understanding of these changes in the pouch by measuring the differences in cellular composition and cell-type-specific gene expression in the pouch compared to pre-pouch and control samples from AC and TI.

Through multiomic analysis, we identified and characterized partial colonic metaplasia of epithelial cell types in UC pouches and provided insights into the mechanisms underlying their differentiation. We identified transcriptional regulators active in colon-like pouch cells that were previously associated with colonic epithelial cells and epithelial progenitors, including CDX1, CDX2^30,31^, EHF^32^ and NFIA^28^. Activity of these TFs was correlated with the expression of genes with colon-specific functions including metabolic (CA2, HMGCS2) and anti-microbial (CEACAM5) activity. While CEACAM5 was expressed in EC2 cells from all samples, CA2 expression was lower compared to AC and only observed in some pouch samples. We also observed an increase in the expression of SATB2 in a subset of pouch EC2 cells. SATB2 is necessary for colonocyte differentiation, and its overexpression is sufficient to convert ileal epithelium into colonic epithelium in mice^31^. However, SATB2 was expressed at lower levels and more heterogeneously in pouch EC2 cells compared to AC EC2. In contrast to AC EC2, pouch EC2 cells also maintained expression of a subset of genes with generally ileal-specific gene expression pattern (APOA4, APOB, FABP6) (Supplementary Data 5), and appeared less mature compared to AC EC2. In addition, pouch EC2 and Goblet2 cells showed evidence of a transcriptional signature enriched for immune and secretory functions. Similar expression changes in epithelial cells have been observed in chronic inflammatory conditions in human IBD and animal models^21,22,33,34^.

Bulk RNA-seq analyses have revealed distinct gene expression shifts indicative of ’regionalization’ in CD^35^, and it will be interesting to understand whether this is indicative of metaplasia in these CD cases.

Changes in gene expression of the pouch mucosa had previously been observed in bulk RNA-seq data^11^. Our study linked these changes to a particular cell population and comparing our cell-type-resolved data with bulk data provided evidence for the presence of EC2 cells in the majority of pouches. This analysis also suggested that the presence of EC2 cells alone was not linked to development of pouchitis. It is possible that the presence of EC2 cells is unrelated to pouchitis and a consequence of establishing pouches. Alternatively, EC2 cells may be a necessary, but not the sole, contributor to development of pouchitis. Familial adenomatous polyposis patients frequently undergo the same procedure to reduce risk of colon cancer yet reportedly rarely develop pouchitis^9^, which suggests that establishment of a pouch alone does not render patients susceptible to inflammation.

Our data underscored the adaptability of the intestinal epithelium in adult tissues, yet the exact progenitor source for the colon-like metaplastic epithelial cells remains to be determined. The differentiation into colon-like and ileum-like enterocyte populations is most pronounced within the pouch’s mature secretory and absorptive cells. However, stem and progenitor populations in pouches diverged into distinct populations as well, and co-clustered closely with their mature counterparts in ileum and colon, suggesting an early stage of differentiation fate. Our analysis integrated cells from different tissues using Harmony which was necessary to account for inter-individual variation. This process facilitated the grouping of broadly similar cells and allowed for joint annotation of cell type clusters within a shared embedding of patient and reference samples. However, this process potentially removed non-technical variation between cell types from patient and controls (e.g. contributed by inflammation) and the resulting embedding might overemphasize similarities between EC2 cells in pouch and AC. Our DEG analysis addressed this concern by directly comparing expression patterns across different cell clusters and identifying both shared and different sets of genes.

While this study provides high-resolution insights into transcriptomic and regulatory signatures using primary human tissues, there are several limitations. The samples were obtained from a small cross-sectional cohort consisting of six individuals. This limitation guided our focus towards epithelial cell types that were both prevalent and exhibited consistent effects across the cohort. The study’s focus does not imply that changes in other cell types, especially in stromal lineage, are unimportant but rather reflects our study’s limited capacity to explore them comprehensively. For example, identification of colon-enriched NRG1+ fibroblasts^30^ in the pouch might suggest the development of a colon-like niche in the pouch. Additionally, single cell genomics data lacks spatial context which is important to study the cellular niches that contribute to the observed cellular changes in the pouch. We generated a small spatial transcriptomics dataset for validation and provided evidence for the expression of key marker genes in pouch and pre-pouch samples. However, these data do not have sufficient power to be used for discovery and the comprehensive characterization of cellular domains and niches.

Nevertheless, these data are a valuable resource for further cellular and molecular studies of IPAA. Our results suggest that UC pouches provide a unique system to study intrinsic and extrinsic cues contributing to epithelial plasticity and how modulation of epithelial identity relates to disease risks in the pouch and IBD in general.

## Methods

### Ethics statement

This study was conducted under the translational core research program protocol (15573A), approved by the Institution Review board of University of Chicago. Licensed gastroenterologists collected biopsy samples and clinical data from patients identified as UC-pouch and non-IBD control subjects. Biopsy sample and patients’ demographic data were obtained from all participants with signed informed consent. Samples and demographic data were anonymized.

### Patient information

Non-IBD controls were consented and recruited by the time of routine colonoscopy. UC-pouch patients were consented and recruited at follow-up pouchoscopy. Non-IBD controls were patients without a history of IBD, an immune related disease, an ongoing infection in intestines, a history of Familial adenomatous polyposis, a history of primary digestive tract malignancies, a history of metastatic malignancies in digestive tract, and symptoms of digestive disorders. The UC-pouch patients had a clinical diagnosis of ulcerative colitis, have a history of ileal pouch-anal anastomosis and were reported free of active disease via macroscopic assessment from a physician during the pouchoscopy. Demographic data of patients are provided in Supplementary Data 1.

### Tissue biopsy sample collection

All biopsies were collected in locations determined by scoping gastroenterologists. Non-IBD controls had up-to-six biopsies obtained from each location during colonoscopy at two separate anatomic regions (terminal ileum and ascending colon). UC-pouch patients had up-to-six biopsies obtained from each region, at two regions (pre-pouch and pouch), during pouchoscopy. In short, mucosal linings were obtained using biopsy forceps following standardized care. Biopsied mucosal tissues from each region were immersed in Advanced DMEM/F-12 [Thermo Fisher 12634010] in an Eppendorf tube and placed on ice for short-term preservation and transfer. The wait times between biopsy and sample processing were under two hours.

### Sample processing

Single cells from mucosal biopsies were acquired using a modified version of previously published protocols^21,36,37^. In brief, intact mucosal tissues were rinsed in 1.5 ml ice-cold PBS [Thermo Fisher 10010023] six times. Mucosal tissues from the same regions were combined and minced by iris scissors. Minced tissues were transferred to 2 ml fresh chelator solution (HBSS Ca/Mg-Free [Thermo Fisher 14175095], 10 mM EGTA [Fisher Scientific 50-255-956], 10 mM HEPES [Thermo Fisher 15630080], and 10% FBS [Gemini Bio 100-106]). The tissues were incubated at 37°C for 15 minutes with end-over-end rotation, then cooled on ice for 10 minutes. The epithelium was separated from lamina propria by mechanical agitation (shaking 20 times). Visualizing epithelial sheets in the solution, the supernatant was collected and spun down at 300xg for 5 minutes at 4°C. The remnant tissue chunks were rinsed in ice-cold PBS and incubated in 2 ml enzyme solution (HBSS Ca/Mg-Free [Thermo Fisher 14175095], 10 mM HEPES [Thermo Fisher 15630080], 200 μg/ml Liberase TM [Millipore Sigma 5401127001] and DNase I [Gold Bio D-301- 100]) at 37°C for 20 minutes. The spun-down epithelium was resuspended and incubated in 2 ml TrypLE express [Thermo Fisher 12605010], at 37°C for 5 minutes. Both incubations were applied with end-over-end rotation. Tissues were gently triturated with a P1000 pipette after incubations and no visible tissue chunks remained after trituration. Cells were filtered through a 40 μM cell strainer [Falcon/VWR 21008–949]. After filtration, epithelial cells and cells from lamina propria were combined and spun down at 300xg for 5 minutes at 4°C. Cell pellet from each region were resuspended in 200 μl cold wash buffer (HBSS Ca/Mg-Free [Thermo Fisher 14175095] and 10% FBS [Gemini Bio 100-106]). Cell suspension was then diluted by 2 ml lysis buffer [Miltenyi Biotec 130-094-183] and sat for 3 minutes at room temperature to eliminate red blood cells. Cells in suspensions were spun at 300g for 5 minutes at 4°C. Cell pellets were resuspended in 200 μl suspension buffer (PBS and 0.04% BSA [Miltenyi Biotec 130-091-376]) and assessed for density and viability with Trypan Blue [Thermo Fisher 15250061]. Only cell suspensions with >85% viability were used for downstream single cell assays. The detailed processing is fully described here (https://www.protocols.io/view/preparation-of-single-cell-suspensions-from-human-q26g7bxqklwz/v4).

### scRNA library preparation and sequencing

Single cells were loaded onto chip G per the manufacturer’s protocol for the Chromium Single Cell 3’ Library (v3.1) [10X Genomics 1000121]. In short, single cells were partitioned into Gel Beads in Emulsion (GEMs) in Chromium controller. GEMs were incubated for cell lysis and barcoded reverse transcription, followed by cDNA amplification. The cDNAs were then enzymatically fragmented and indexed. Approximately 8,300 single cells were loaded to a channel with a target recovery for 5,000 cells. Libraries were sequenced on Illumina NovaSeq platform.

### scATAC library preparation and sequencing

Single nuclei were isolated from single cell suspensions following the manufacturer’s protocol for the Chromium Single cell ATAC (v1.1). Briefly, single nuclei were transposed before loading to chip H. Transposed nuclei were partitioned into GEMS in Chromium controller. GEMs were incubated for barcoding, followed by fragment indexing. Approximately 8,000 single nuclei were transposed and loaded to a channel with a target recovery for 5,000 nuclei. Libraries were sequenced on Illumina Novaseq platform.

### scRNA-seq preprocessing and quality control

scRNA-seq gene expression raw sequencing data were processed using the Cell Ranger software v7.0.1 and the 10X human transcriptome GRCh38-2020-A as the reference. The expected number of cells were auto-estimated in Cell Ranger.

SoupX was used with default parameters for the estimation and removal of cell free mRNA contamination for each sample separately^38^. Clustering information from Cell Ranger output was loaded to estimate the contamination fraction. R (v4.1.0) and Seurat (v4.2.0) were used to pool single cell counts and for downstream analyses^39–43^. Cells were filtered for more than 500 UMIs, fewer than 20,000 UMIs, more than 200 unique genes, fewer than 6,000 unique genes, less than 50% of reads mapping to mitochondrial genes and less than 40% of reads mapping to ribosomal genes and a ‘log10GenesPerUMI’ score higher than 0.7 (to remove contamination with low complexity cell types like red blood cells). Genes expressed in fewer than 120 cells were filtered out (average 5 cells per sample in all 24 samples). Doublet score was calculated using Scrublet (v0.2.3)^44^, and a cut-off score of 0.2 was applied to exclude doublets. The cells with unexpected co-expression of different lineage makers such as CD3D and EPCAM were also excluded as doublets. Gene expression for each cell was normalized and log transformed. Cell cycle scores (G2/M phase, S phase) were calculated using the expression of cell cycle genes listed in Seurat (v4.2.0). Cell cycle score (the difference between the G2M and S phase scores), the percentage of mitochondrial reads and unique molecular identifiers (UMIs) were regressed out before scaling the data.

### scRNA-seq dimensionality reduction, clustering analysis and visualization

We restricted the expression matrix to the 2000 most highly variable genes identified by fitting the mean-variance relationship and high-quality cells noted above as input for principal component analysis (PCA). Principal components were corrected using Harmony (v0.1.0)^14^ to remove patient-specific effects. This strategy facilitated joint clustering and annotation of cells from all samples in a single embedding. Cells were clustered using the Louvain algorithm^45^ (resolution 0.3–1.5) for modularity optimization using the kNN graph as input. Cell clusters were visualized in Uniform Manifold Approximation and Projection (UMAP) embedding^46^.

The union dataset was divided into different lineages based on clustering analysis and marker gene expression. Cells from epithelial, T cell, B cell, myeloid, stromal lineages were subset for further analysis. For each lineage, we repeated the process of lineage-specific QC, dimensionality reduction, principal component correction, Louvain clustering and UMAP visualization as described above. Cell identities for cell subtypes in each lineage were assigned based on markers outlined in the following section.

### Cell-type annotation based on scRNA-seq data

Cell-type clusters were annotated based on marker genes previously reported as markers of intestinal cell populations^13^.

Epithelial lineage cells were defined on the basis of *EPCAM* and *FABP1* expression. Stem cells had specific expression of *LGR5* and *SMOC2*. Transit-amplifying cells were classified based on the highest expression of *MKI67*, *TOP2A* and *TUBA1B*. Enterocyte clusters with ileum functional signatures include EC1-1 (*SI*, *GSTA2*) and EC1-2 (*SI*, *SLC15A1*, *APOA4*, *CUBN*). Enterocytes in EC1-1 cluster is at an earlier differentiation stage than EC1-2, with higher *OLFM4* expression. Enterocyte clusters with colonic signatures include EC2-1 (*CD24*, *CA2*) and EC2-2 (*CD24*, *CA2*, *CEACAM5*). EC2-1 cluster is at an earlier differentiation stage than EC2-2, with higher *OLFM4* expression. Goblet clusters were identified based on high *MUC2* and *TFF3* expression. Goblet2 cluster is a dominating type of goblet cells in the colon with higher *BEST2* and *KLK1* expression compared to Goblet1. Paneth cells (*DEFA5*, *ITLN2*, *REG3A*) and Microfold-like cells (*CCL20*) are observed in ileum, pre-pouch and pouch. BEST4 enterocytes, enteroendocrine cells (*CHGA*, *KCNB2*, *RIMBP2*) and tuft cells (*POU2F3*, *FYB1*) are identified in all four regions.

T cell lineage cells were defined based on positive *CD3D* expression. CD4 T cells had expression of *CD4*, but not *CD8A*, and vice versa for CD8 T cells. Tissue resident memory T cells were characterized by high *CD69* expression. CD4 tissue resident memory T cells were further divided based on *ITGAE* (encodes CD103) expression. CD103^-^CD4 T cells had higher *IL7R* expression, while CD103^+^CD4 T cells had higher *IL17A* and *IL26* expression. CD8 tissue resident memory T cells include KLRG1 positive and negative clusters, indicating different cytotoxic functions. Central memory T cells had the highest *LEF1*, *TCF7* and *CCR7* expression. Regulatory T cells were characterized by high *CTLA4* and unique *FOXP3*, *IL2RA* expression. Two TRDC^high^ clusters were classified as gamma-delta T cells (ENTPD1, GZMA^high^) and NK cells (NCR1^+^KLRF1^+^CD3D^-^) respectively. NK T cells expressed *CD3D*, *FGFBP2*, as well as NK genes *NCR1*, *KLRF1*. Mucosal-associated invariant T (MAIT) cells showed high *SLC4A10* and *NCR3* expression. Innate lymphoid cells (ILCs) (*PRKG1*, *PCDH9*, *AFF3*, *AREG*, *IL1R1*, *IL23R*, *KIT*) are a diverse group of immune cells, but we were not able to further sub-cluster ILCs in this atlas.

B cell lineage cells were defined based on high *CD79A* and *MS4A1* expression, and plasma cell clusters had high *XBP1* and *IGHA1* expression. *BCL6* expression defines germinal center B cells, which is crucial for somatic hypermutation and class switch recombination. Naïve B cells had the highest expression of immunoglobulin heavy chains D (*IGHD*). Memory B cells were characterized by positive *CD27* expression and negative *IGHD*. Class switch IgA and IgG plasma cells showed expression of the *XBP1*, and immunoglobulin heavy chains A and G, respectively.

Myeloid lineage cells were defined based on high expression of *TYROBP* and *CPA3*. Monocytes and macrophages both are *CD14* and *C1QA* positive. Monocytes had high expression of *VCAN*, *FCN1* and *EREG*, and macrophages had highest expression of *CD163* and *MMP12*. Among dendritic cells (DCs) we observed cDC1 (*CLEC9A*) and cDC2 (*CD1C*), and lymphoid DCs (*LAMP3*). Mast cells were identified by *CPA3*, *KIT* and *TPSB2*. Neutrophils were classified by high expression of *FCGR3B* (encodes Fc gamma receptor 3B).

Stromal lineage cells were defined based on high expression of *IGFBP7* and *COL3A1*. Fibroblasts include three main populations, fibroblast1 (*ADAMDEC1*, *CCL11*, *CCL13*), fibroblast2 (*NRG1*, *NPY*, *PTGS1*), fibroblast3 (SOX6 high). Endothelial cells (*PECAM1*) include arterial endothelium population (*HEY1*, *EFNB2*), venous endothelium population (*ACKR1*) and lymphatic endothelium population (*PROX1*, *LYVE1*, *CCL21*). Pericytes were identified by high *NOTCH3* expression, and a contractile pericyte subset were characterized by high *PLN*, *RERGL* and *KCNAB1* expression. Myofibroblasts and smooth muscle cells both show high actin (*ACTA2*) and transgelin (*TAGLN*) expression, while Myofibroblasts lack smooth muscle marker desmin (*DES*). Glial cells were characterized by *S100B* and *NRXN1*.

### Marker gene detection and differential expression analysis

In order to account for patient-specific effects when calculating the differentially expressed genes in enterocytes across regions, we used a linear mixed model (model formula = cluster + (1|Patient_ID)) implemented in variancePartition (v1.21.1)^47,48^ to quantify gene expression attributable to individual patients. We verified that other confounding factors are not contributing a significant amount of variance, and we did not include additional factors in the linear model. The cell number threshold to create pseudo-bulk cell clusters was set considering the size of the cell type population (enterocyte groups: 200 cells, goblet cells: 30 cells, stem cell: 30 cells). Differentially expressed genes in different cell type clusters were identified by the ‘FindAllMarkers’ function (Wilcoxon Rank Sum test with default parameters) in Seurat. The other pairwise differential tests were performed by the ‘FindMarkers’ function (Wilcoxon Rank Sum test with default parameters).

### Gene set enrichment analysis

Gene set categories (H, C2, C5, C7 and C8) implemented in Molecular Signatures Database (MSigDB) were pulled and examined for each list of differentially expressed genes using msigdbr package (v7.5.1). R package ‘org.Hs.eg.db’ (v3.14.0) was used to map gene identifiers. Gene set enrichment analyses were performed using the ‘clusterProfiler’ R package with the ‘GSEA’ R function (pvalueCutoff = 0.05, pAdjustMethod = “fdr”, minGSSize = 10, maxGSSize = 5000)^49^.

### Enrichment score of signature gene sets

‘Area Under Curve’ (AUC) was used to calculate the enrichment of a gene set for each cell, as implemented in AUCell (v1.16.0)^50^. For each cell, the genes are ranked from highest to lowest value. The genes with the same expression value are shuffled. Therefore, genes with expression ‘0’ are randomly sorted at the end of the ranking. The AUC value is representing the fraction of genes from the pathway within the top 5% (aucMaxRank=0.05) of all the genes in our dataset.

### Immunostaining of human biopsy

Biopsies matched to the patients assessed in single-cell RNA-seq experiments were obtained from the pathology archives at UChicago Medicine under an IRB-approved protocol. Serial sections were prepared from the paraffin-embedded material. Immunohistochemical staining was performed as previously described^51^. Briefly, sections were deparaffinized and rehydrated, heat/pressure-treated for 20 min with citrate buffer (pH 6), bleached with 0.3% hydrogen peroxide for 30 minutes, and blocked with horse serum. Primary antibodies were prepared in 0.3% Triton-X and 4% horse serum in phosphate-buffered saline (PBS) and incubated with the tissue overnight at 4C. Primary antibodies used were rabbit anti-APOA4 (1:1,000, Sigma HPA001352), rabbit anti-FABP6 (1:200, Sigma HPA012601), mouse anti-CA2 (Santa Cruz Biotechnology sc- 48351), and mouse anti-CEACAM5 (R&D Systems MAB41281). After washing with PBS, sections were exposed to horseradish peroxidase-conjugated secondary antibodies (Immpress kit, Vector Labs) for 2 h at room temperature, and signal was developed for 2-10 minutes with a diaminobenzidine (DAB) kit (Vector Labs). Tissue was counterstained with methyl green prior to dehydration, clearing, and coverslipping. A whole-slide scanner (Olympus) mounted with a 20X objective was used to obtain images of the staining. Analysis was performed with QuPath^52^.

### Histology evaluation and quantitation of enteritis score

Histological assessment of hematoxylin-and-eosin stained biopsy sections was performed by a clinical pathologist (C.W.) blinded to the slide labels. The scoring method used was based on previous assessment^53^ of colonic inflammation and mucosal damage. The score ranged from 0 (no abnormalities) to 3 (severe damage), with intermediate grading defined as follows:

| Score | Category | Description |
| --- | --- | --- |
| 0 | Quiescent or Inactive | No active inflammation (no intraepithelial neutrophils) |
| 1 | Mild Active | Presence of intraepithelial neutrophils |
| 2 | Moderate Active | Presence of crypt abscesses |
| 3 | Severe Active | Erosions or ulcers |

### Bulk-RNA data deconvolution

We used cell type specific gene expression from our scRNA-seq data to characterize cell type compositions from bulk RNA-seq data (GSE81266), following the pipeline described in MuSiC (Multi-subject Single-cell Deconvolution, v 1.0.0) package^24^, with default parameters.

### scATAC analysis workflow preprocessing and quality control

scATAC-seq raw sequencing data were processed using the Cell Ranger ATAC v2.0.0, and the 10X human genome GRCh38-2020-A-2.0.0 as the reference. Further filtering was conducted using ArchR (v1.0.3)^26^. Only high-quality individual cells were kept for downstream analysis (doublet filter ratio = 2.0, minimum fragments per cell = 2,000, minimum TSS enrichment score = 6).

### Dimensionality reduction and clustering analysis

We used ArchR to convert the fragments file into a 500bp tile matrix of fragment counts and generated a gene score matrix using the default model (model 42) with default parameters, taking into account chromatin accessibility within a gene body as well as proximally and distally from the TSS. We performed latent semantic indexing (LSI) on the tile matrix and retained the top 100 LSI vectors (‘addIterativeLSI’ function, 6 iterations including 5 different clustering resolutions: 0.5, 1, 1.5, 2, 2). The first LSI vector was removed because of high correlation with sequencing depth (correlation score > 0.75). The LSI vectors were further corrected Harmony (v0.1.0)^14^ using PatientID as variable. The remaining 99 LSI vectors were used to create the UMAP (nNeighbors = 30, minDist = 0.2). Cell clusters were identified using the ‘addClusters’ function with multiple different resolutions.

### Annotation of scATAC cells

The scRNA-seq expression matrix was integrated with the scATAC-seq gene score matrix using the ‘addGeneIntegrationMatrix’ function from ArchR. It identified the corresponding cells across datasets using Seurat’s mutual nearest neighbors algorithm, thus transferring the labels from scRNA data to scATAC data. The labels in scATAC data were further curated based on specific marker gene score distribution. A notable exception were TA cells which did not have a clear corresponding cell type in scATAC-seq. We interpreted this as a consequence of the annotation of TA cells based on the expression of cell cycle genes which might not show strong corresponding changes in chromatin accessibility at their genomic loci. We therefore removed TA designation from scATAC-seq and used progenitor/immature instead. In addition, M-like cells which represent a small proportion of cells in the scRNA-seq data (<1.7%) and which we transcriptionally distinguished from enterocytes solely by expression of CCL20 showed a diffuse distribution, so we removed this label.

### Accessible chromatin peak calling

We merged cells within each cell group for the generation of pseudo-bulk replicates and then merged these replicates into a single insertion coverage file. The ‘addReproduciblePeakSet’ function was used to get insertions from coverage files, call peaks, and merge peaks to get a “Union Reproducible Peak Set” (peaksPerCell = 500, maxPeaks = 150,000, minCells = 50, extsize = 150, cutOff = 0.05, extendSummits = 250).

### Differential accessible region (DAR) analysis

To identify cell-type specific marker peaks, a single-cell insertion count matrix was created using the function ‘addPeakMatrix’ in ArchR. To perform DA, we used ‘getMarkerFeatures’ (Wilcoxon rank-sum test, maxCells = 20,000). To control for technical variation, we accounted for “TSSEnrichment” and ”log10(nFrags)” when selecting a matched null group for marker feature identification (FDR < 0.01, logFC > 1).

### Motif annotation and TF motif enrichment

We used 870 human motif matrices from the CisBP^54^ database to annotate each of the peaks in the union peak set for each lineage separately by the ‘addMotifAnnotations’ function. The hypergeometric test was used to assess the enrichment of the number of times a motif overlaps with a given set of peaks, compared to random expectation (adjusted p-value < 0.001). The top 20 transcription factor motifs significantly enriched for each cell type were filtered based on top 3,000 broadly expressed genes (ranked by the proportion of cells expressing this gene). Tn5 bias-subtracted TF footprinting analysis was performed using the getFootprints function in ArchR for pseudo-bulk aggregates of cells in each cell type and tissue location.

### ChromVAR deviation score

We computed background peaks controlling for total accessibility and GC-content by the function ‘addBgdPeaks’ and used the function ‘addDeviationsMatrix’ function to calculate the TF *ChromVAR* activity score. The TFs for each enterocyte group were further further filtered based on positive or negative TF activity-expression correlation across enterocyte cell types.

### Trajectory analysis

The trajectory analysis was performed using the ‘addTrajectory’ function in ArchR by specifying the cluster order in two enterocyte lineages (EC1 lineage: Stem1, EC1-1, EC1-2; EC2 lineage: Stem2, EC2-1, EC2-2). We visualized trajectory-dependent changes of TF activity and gene expression along differentiation trajectories by ordering cells by pseudotime.

### Domain of regulatory chromatin analysis

The domains of regulatory chromatin (DORCs) were identified using the standard workflow introduced in FigR (v0.1.0)^29^. The integrated gene expression matrix and peak matrix were used to infer cis-regulatory interactions. Here the integrated gene expression matrix was obtained from the multiomic integration analysis introduced above. The minimum number of interactions (cutoff=7) was used to determine a significant DORC gene.

### Gene regulatory network construction

Highly correlated links between gene expression and region accessibility were identified by ‘addPeak2GeneLinks’ (corCutOff = 0.6), as candidate cis-regulatory elements (cCREs). The core transcription factors were first identified by the TF enrichment analysis of different groups of differentially accessible regions between enterocyte groups. These groups of TFs were then filtered for highly expressed genes. Finally, only the most significantly positively and negatively TF regulators in the same TF family were preserved as nodes in the network. We filtered the gene targets for the top 100 most significantly linked genes, the well-known functional genes (within top 300 most significantly linked genes), and top 100 DORC genes (within top 300 most significantly linked genes).

### Xenium 10x spatial transcriptomics

Samples were treated following the Xenium 10x Genomics spatial transcriptomics protocol (CG000749). FFPE samples were sectioned onto a Xenium slide by the Human Tissue Resource Center at the University of Chicago. Xenium data were generated at the NuSeqCore at Northwestern University using a set of 422 probes. 322 probes from the pre-designed Xenium Human Colon Gene Expression Panel Kit (1000642) and 100 probes designed to target pouch relevant genes (Supplementary Data 11). Raw data were processed using Xenium Analyzer. Cell segmentation was performed with the Xenium Cell Segmentation Add-on Kit (1000662).

### Spatial domain determination vis IRIS

The xenium 10x spatial transcriptomics data of two pouch and two pre-pouch slices from the same individual were loaded in R version (v4.3.1) using Seurat (v5.1.0)^39–43^ and analyzed with IRIS (v1.0)^23^. In our scRNA-seq data, we merged cell type labels for enterocytes, goblets, T cells, B cells, plasma cells, myeloid cells, stromal cells and endothelial cells. scRNA count matrices were then extracted and used to construct IRIS objects using the function createIRISObject, with minCountGene set to 20 and minCountSpot set to 5. Function IRIS_spatial was called to isolate main spatial domains with numCluster set to 5. Differentially expressed genes were identified using the function FindMarkers from Seurat with default parameters.

## Data availability

The scRNA-seq and scATAC-seq pouch data analyzed in this study have been deposited in the GEO database under accession code GSE242087. The reference terminal ileum and ascending colon samples were deposited in the GEO database under accession code GSE266616 (scRNA) and GSE273194 (scATAC). The one-to-one correspondence of sample records in the GEO database is detailed in Supplementary Data 12. The processed scRNA-seq gene count matrix, lineage-specific scATAC-seq peak matrix and cell type annotation are available at Zenodo doi: 10.5281/zenodo.14165540. Source data are provided with this paper.

## Code availability

Code to reproduce data processing and analysis are available at: https://github.com/EthanZhaoChem/Code_documentation_2023_IBD_pouch_paper

## Supporting information

description of supplementary data files

## Acknowledgements

This work was enabled through grants from the National Institutes of Health R35GM142986 (to S.P.), RC2DK122394, T32DK007074, P30DK042086, a grant from the GI Research Foundation (to E.B.C.), and a grant from The Leona M. and Harry B. Helmsley Charitable Trust to the University of Chicago. This work was completed in part with resources provided by the University of Chicago Research Computing Center. We thank The University of Chicago Human Tissue Resource Center (RRID:SCR_019199), especially Xin Jiang for their assistance with preparation of tissue sections. We thank the NUSeq Core at Northwestern University, especially Jennifer Wai for supporting the generation of Xenium data. We thank Christian Jones and Sarah Sumner for assistance with editing.

## Author contributions

E.B.C., A.B. and S.P. conceived the study and supervised this project. Y.Z. performed data analysis. R.Z. performed experiments and generated data. C.M.C., J.K. contributed to sample collection and experiments. B.X., Z.J. and M.C. contributed to data analysis. C.R.W. performed histological assessment of tissue sections. C.Y.L. and M.K. performed IHC and microscopy. D.T.R., M.S., S.C., J.H., A.F., S.D. contributed to study design and data interpretation. S.P. and Y.Z. wrote the manuscript with input from all authors.

