## supplemental figures for "Multiomic analysis reveals cellular, transcriptomic and epigenetic changes in intestinal pouches of ulcerative colitis patients"

**Supplementary Fig. 1** | **Data overview. a** Metadata of the patients involved in this study. **b** Number of cells in each biopsy sample that have been analyzed after QC in scRNA-seq analysis. **c** UMAP split by regions highlights that the largest cross-region differences lie in epithelial lineage. **d** Expression of markers for different cell lineages. Color indicates scaled mean expression in each lineage, dot size represents the proportion of cells expressing that marker. Source data are provided as a Source Data file.

**Supplementary Fig. 2** | **Expression of markers for different cell types in epithelial lineage.** The color shows scaled mean expression in each cell type. The dot size represents the proportion of cells expressing that marker.

**Supplementary Fig. 3** | **Expression of markers for different cell types in T cell lineage.** The color shows scaled mean expression in each cell type. The dot size represents the proportion of cells expressing that marker.

**Supplementary Fig. 4** | **Expression of markers for different cell types in (a) B cell lineage (b) myeloid lineage.** The color shows scaled mean expression in each cell type. The dot size represents the proportion of cells expressing that marker.

**Supplementary Fig. 5** | **Expression of markers for different cell types in stromal cell lineage.** The color shows scaled mean expression in each cell type. The dot size represents the proportion of cells expressing that marker.

**Supplementary Fig. 6** | **scRNA-seq data. a** Epithelial UMAPs showing the expression of markers for epithelial stem cells (LGR5), transit-amplifying cells (MKI67), early stage enterocytes (OLFM4), ileum-like enterocytes (FABP6), colon-like enterocytes (CEACAM5). **b** Pouch enterocytes are split into two populations: pouch EC1 and pouch EC2. **c** Violin plots for FABP6 and CEACAM5 expression in 5 enterocyte groups. The analysis was conducted between single cells from two cell populations, with each single cell considered a biological replicate. The number of cells in each group is as follows: AC (7,072), pouch EC2 (3,523), pouch EC1 (3,581), PP (7,327), TI (17,633). Statistical significance was assessed using a two-sided Wilcoxon rank-sum test. **d** Cell proportion in epithelial lineages. A two-sided Wilcoxon rank-sum test was performed to compare samples from different tissue regions, with each sample representing a biological replicate. "ns" indicates $p > 0.05$. **e** UMAP of all epithelial cells without batch effect removal, colored by patient ID and biopsy location, respectively. **f** UMAP of all epithelial cells after batch effect removal, colored by patient ID and biopsy location, respectively. **g** UMAP of epithelial cells from pouch and pre-pouch after batch effect removal, colored by biopsy location. **h** UMAP of epithelial cells from pouch and pre-pouch after batch effect removal, colored by cell type annotation. Source data are provided as a Source Data file.

**Supplementary Fig. 7 | Gene expression validation.** The z-score of the average expression of each gene in (**a, b**) Enterocytes or (**c**) Goblets per sample was calculated and plotted across samples for each gene. All highlighted genes in Fig. 2d, Fig. 3a and Fig. 3b were plotted to demonstrate the consistency between the reference dataset **a** Kong et al.[1] or (**b, c**) Smillie et al.[2] and our study. Source data are provided as a Source Data file.

**Supplementary Fig. 8 | scRNA data. a** UMAP shows cell types in T cell lineage. **b** The proportion of different cell types in different regions. **c** Cell types that show proportional differences across regions. A two-sided Wilcoxon rank-sum test was performed to compare samples from different tissue regions, with each sample representing a biological replicate. "ns" indicates $p > 0.05$. **d** The cell type proportions in each sample. Source data are provided as a Source Data file.

**Supplementary Fig. 9 | scRNA data. a** UMAP shows cell types in B cell lineage. **b** The proportion of different cell types in different regions. **c** The cell type that shows proportional differences across regions. A two-sided Wilcoxon rank-sum test was performed to compare samples from different tissue regions, with each sample representing a biological replicate. "ns" indicates $p > 0.05$. **d** The cell type proportions in each sample. Source data are provided as a Source Data file.

**Supplementary Fig. 10 | scRNA data. a** UMAP shows cell types in myeloid cell lineage. **b** The proportion of different cell types in different regions. **c** The cell type proportions in each sample. Source data are provided as a Source Data file.

**Supplementary Fig. 11 | scRNA data. a** UMAP shows cell types in stromal cell lineage. **b** The proportion of different cell types in different regions. **c, d** Cell types that show proportional differences across regions. A two-sided Wilcoxon rank-sum test was performed to compare samples from different tissue regions, with each sample representing a biological replicate. "ns" indicates $p > 0.05$. **e** The cell type proportions in each sample. **f** Average gene expression from bulk RNA-seq data of pouch and prepouch biopsies without overt pouchitis at the time of pouchoscopy (Huang et al.[3]). A two-sided Wilcoxon rank-sum test was used to compare gene expression between different sample groups, with each sample treated as a biological replicate. **g** A Spearman correlation analysis was performed to evaluate the relationship between *NRG1* expression and *CEACAM5* expression in pouch biopsies without overt pouchitis at the time of pouchoscopy (Huang et al.[3]). Source data are provided as a Source Data file.

**Supplementary Fig. 12 | scRNA-seq data. a** The proportions of epithelial cell types in each sample. Volcano plot shows the top differentially expressed genes comparing different enterocyte groups: (**b**) PP enterocytes vs pouch EC2, (**c**) TI vs AC, (**d**) pouch EC2 vs AC, (**e**) pouch Goblet2 vs AC Goblet. A linear mixed model, accounting for patient-specific effects, was used to evaluate both upregulation and downregulation (two sided test) of genes (Methods). P-values were adjusted for multiple comparisons using the false discovery rate (FDR) method. Source data are provided as a Source Data file.

**Supplementary Fig. 13** | **Differential expression between select epithelial cell clusters. a** Number of differentially expressed genes that are shared across different contrasts. **b** Volcano plot shows the top differentially expressed genes comparing pouch EC2 vs pouch EC1. In (**a, b**), a linear mixed model, accounting for patient-specific effects, was used to evaluate both upregulation and downregulation (two sided test) of genes (Methods). P-values were adjusted for multiple comparisons using the false discovery rate (FDR) method.

**Supplementary Fig. 14** | **scRNA data.** Volcano plot shows the top differentially expressed genes comparing different enterocyte groups (**a**) PP vs TI, (**b**) PP vs pouch EC1. In (**a, b**), a linear mixed model, accounting for patient-specific effects, was used to evaluate both upregulation and downregulation (two sided test) of genes (Methods). P-values were adjusted for multiple comparisons using the false discovery rate (FDR) method. (**c**) Differentially expressed genes comparing all EC2 vs EC1. **d** Number of differentially expressed genes that are shared across contrasts in different cell types. In **(c, d)**, differential expression was assessed using a two-sided Wilcoxon rank-sum test, with p-values adjusted for multiple comparisons using the Bonferroni correction.

**Supplementary Fig. 15** | A larger pool of control samples (GSE266616) were used to examine the gene expression variance across different cell type-sample pseudo bulk clusters. Top principle components (explained variance > 1%) calculated from the pseudo-bulk wise gene expression matrix were plotted to highlight the significant role that cell type differences play in genetic variance.

**Supplementary Fig. 16** | **a** We extended Figure 3 with a larger pool of control samples to show the consistent gene expression pattern across different ethnic backgrounds. **b** Z scores of average gene expression from bulk RNA-seq data of healthy pouches (Huang et al.[3]) were plotted to confirm the significant gene expression differences between the pouch and prepouch enterocytes identified in our scRNA study. Source data are provided as a Source Data file.

**Supplementary Fig. 17** | **scATAC data. a** The number of cells in each biopsy sample that have been analyzed after QC in scATAC-seq analysis. **b** The epithelial cell type proportions in each sample. **c** Gene scores for marker genes of different epithelial subpopulations overlayed onto UMAP of epithelial cells. **d** The number of peaks that were initially called in each cell type. Source data are provided as a Source Data file.

**Supplementary Fig. 18** | **scATAC data. a** UMAP shows different cell types in T cell lineage. **b** The proportion of different cell types in different regions. **c** The cell type proportions in each sample. **d** Marker peaks for each cell type. **e** Enriched TF motifs in each cell type. Enrichment was assessed using the hypergeometric test (one-sided), with p-values adjusted using the Bonferroni correction to account for multiple comparisons. Source data are provided as a Source Data file.

**Supplementary Fig. 19** | **scATAC data. a** UMAP shows different cell types in B cell lineage. **b** The proportion of different cell types in different regions. **c** The cell type proportions in each sample. **d** Marker peaks for each cell type. **e** Enriched TF motifs in each cell type. Enrichment was assessed using the hypergeometric test (one-sided), with p-values adjusted using the Bonferroni correction to account for multiple comparisons. Source data are provided as a Source Data file.

**Supplementary Fig. 20** | **scATAC data. a** UMAP shows different cell types in myeloid cell lineage. **b** The proportion of different cell types in different regions. **c** The cell type proportions in each sample. **d** Marker peaks for each cell type. **e** Enriched TF motifs in each cell type. Enrichment was assessed using the hypergeometric test (one-sided), with p-values adjusted using the Bonferroni correction to account for multiple comparisons. Source data are provided as a Source Data file.

**Supplementary Fig. 21** | **scATAC data. a** UMAP shows different cell types in stromal cell lineage. **b** The proportion of different cell types in different regions. **c** The cell type proportions in each sample. **d** Marker peaks for each cell type. **e** Enriched TF motifs in each cell type. Enrichment was assessed using the hypergeometric test (one-sided), with p-values adjusted using the Bonferroni correction to account for multiple comparisons. Source data are provided as a Source Data file.

**Supplementary Fig. 22** | **a** Gene score of each cell type in scATAC-seq clusters. The markers are the same group of genes as scRNA analysis. **b** Sample wise within lineage cell type proportion in each region. The cell groups with fewer than 50 cells are labeled with '-' to indicate low confidence value.

**Supplementary Fig. 23** | **scATAC data. a** Number of differentially accessible peaks that are shared across different contrasts. **b** Enriched TF motifs in each differential test. Enrichment was assessed using the hypergeometric test (one-sided), with p-values adjusted using the Bonferroni correction to account for multiple comparisons. **c** TFs are categorized into activators and repressors. **d** The normalized gene expression and scaled average chromVAR score in 5 enterocyte groups. **e** The logos of repressive TFs.

**Supplementary Fig. 24** | Tn5 bias-subtracted normalized transcription factor footprinting.

**Supplementary Fig. 25** | **scATAC data. a** Differentially accessible peaks comparing Stem2 and Stem1 and enriched TF motifs. **b** Differentially accessible peaks comparing Goblet2 and

Goblet1 and enriched TF motifs. In (**a, b**), differential accessibility was assessed using a two-sided Wilcoxon rank-sum test, with p-values adjusted for multiple comparisons using the false discovery rate (FDR) method. Enrichment was assessed using the hypergeometric test (one-sided), with p-values adjusted using the Bonferroni correction to account for multiple comparisons.

**Supplementary Fig. 26** | **Multiomic analysis.** For genes with at least one linked peak (Methods), the number of linked peaks per gene was calculated and plotted as a distribution. Genes with a higher number of linked peaks exhibit a larger proportion of differentially expressed genes (DEGs, purple) compared to those with fewer linked peaks. A linear mixed model, accounting for patient-specific effects, was used to evaluate DEGs (two-sided test; Methods). P-values were adjusted for multiple comparisons using the false discovery rate (FDR) method. DEGs were selected in Supplementary Data 3 from all five comparison groups, with criteria of FDR $< 0.05$ and $|logFC| > 1$. Source data are provided as a Source Data file.

**Supplementary Fig. 27** | **Multiomic analysis. a** Peaks highly correlated with *APOA4* expression show higher accessibility in terminal ileum, pre-pouch and pouch EC1 enterocytes. **b** Peaks highly correlated with MAF gene expression show higher accessibility in terminal ileum, pre-pouch and pouch EC1 enterocytes. **c** Peaks highly correlated with *SATB2* expression show higher accessibility in ascending colon and pouch EC2 enterocytes.

Supplementary Figure 1

**a**

| Individual ID | Biopsies | Years after surgery |
|---|---|---|
| P1 | Paired pouch and pre-pouch | 14 |
| P2 | Paired pouch and pre-pouch | 9 |
| P3 | Paired pouch and pre-pouch | 23 |
| P4 | Paired pouch and pre-pouch | 31 |
| P5 | Paired pouch and pre-pouch | 10 |
| P6 | Paired pouch and pre-pouch | 14 |
| C1 | Paired ileum and colon | NA |
| C2 | Paired ileum and colon | NA |
| C3 | Paired ileum and colon | NA |
| C4 | Paired ileum and colon | NA |
| C5 | Paired ileum and colon | NA |
| C6 | Paired ileum and colon | NA |

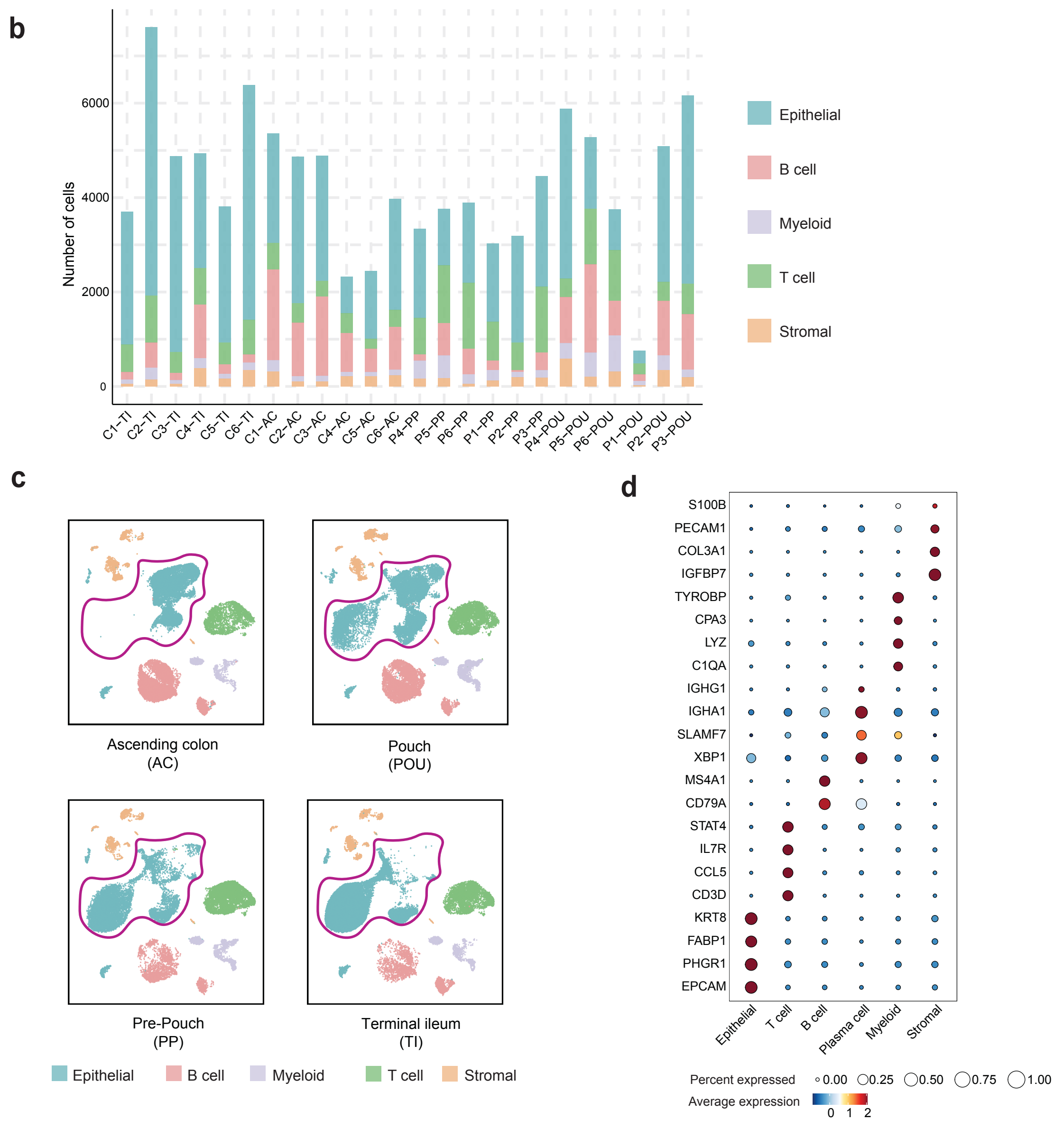

Supplementary Figure 2

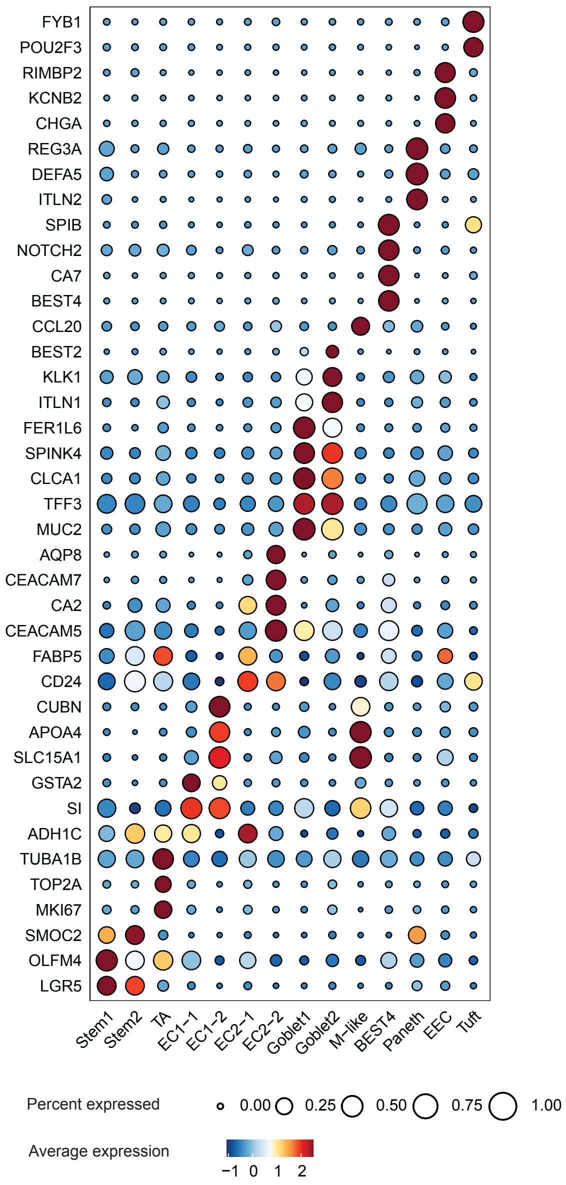

Supplementary Figure 3

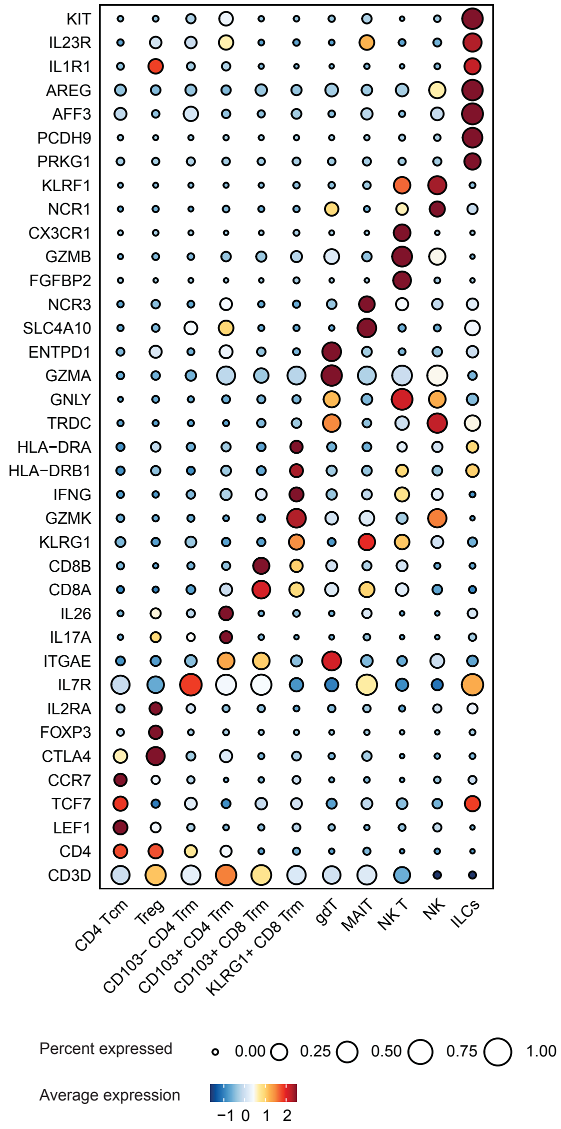


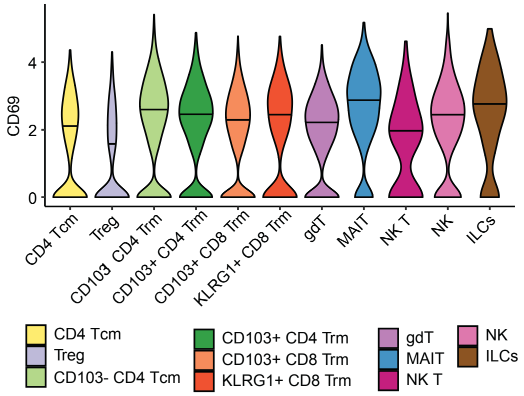

### Supplementary Figure 4

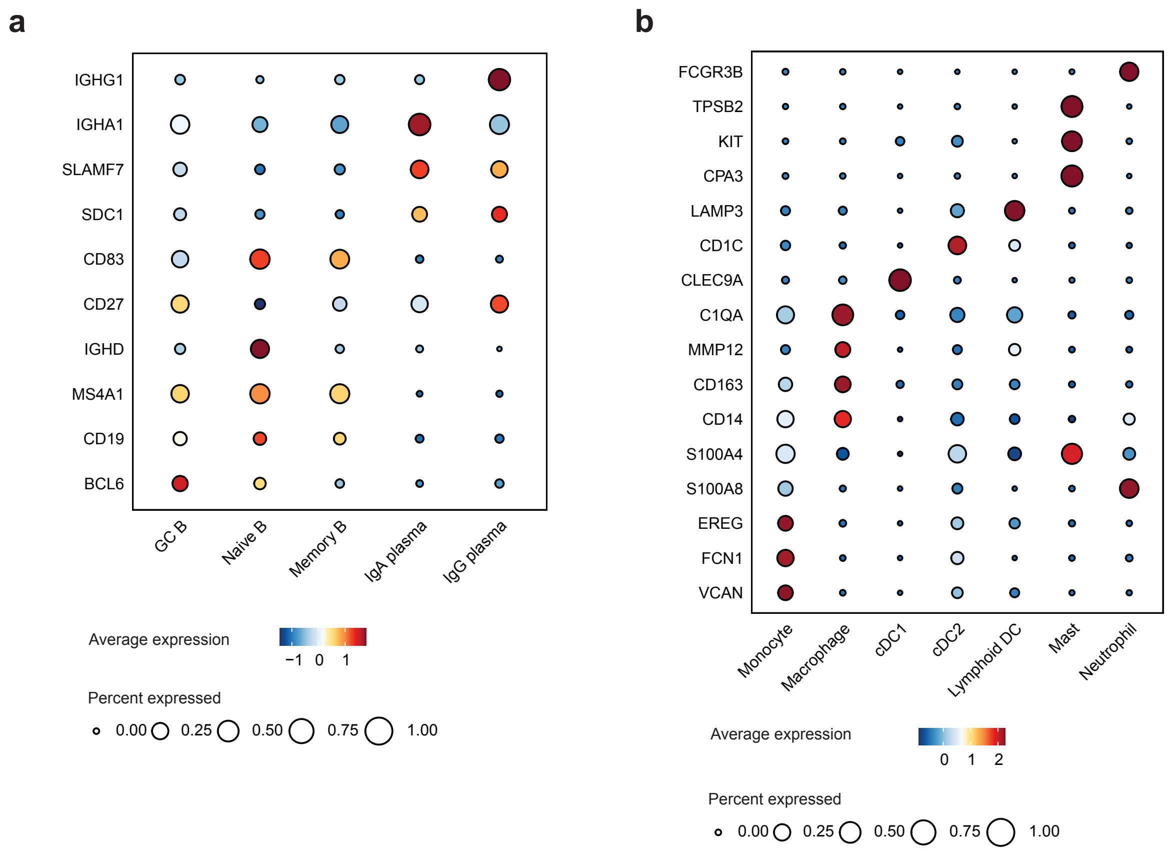

### Supplementary Figure 5

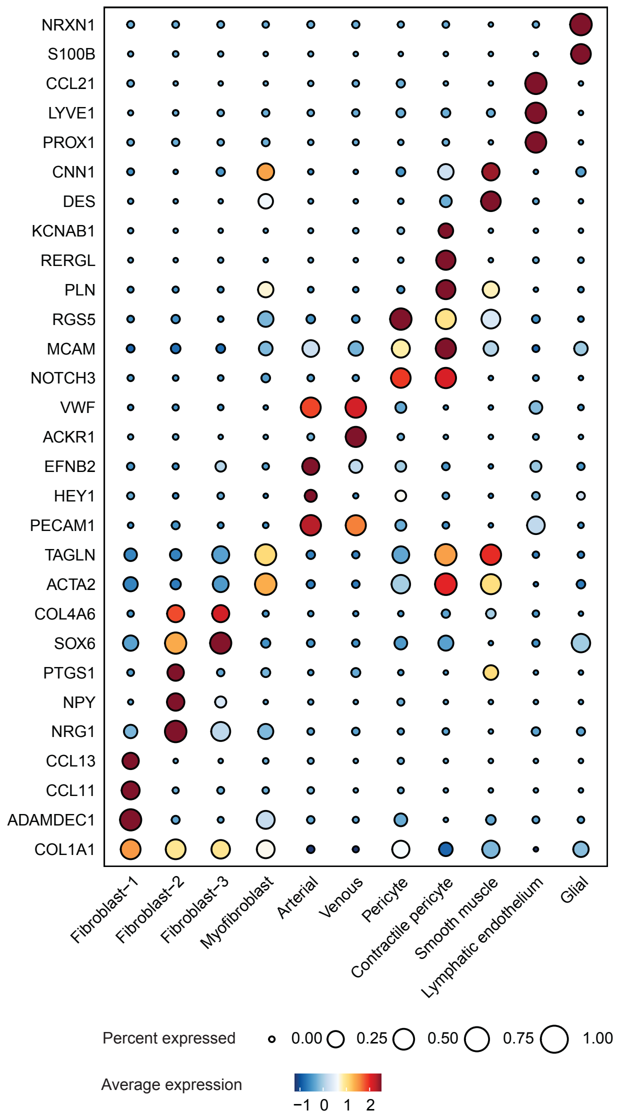

### Supplementary Figure 6

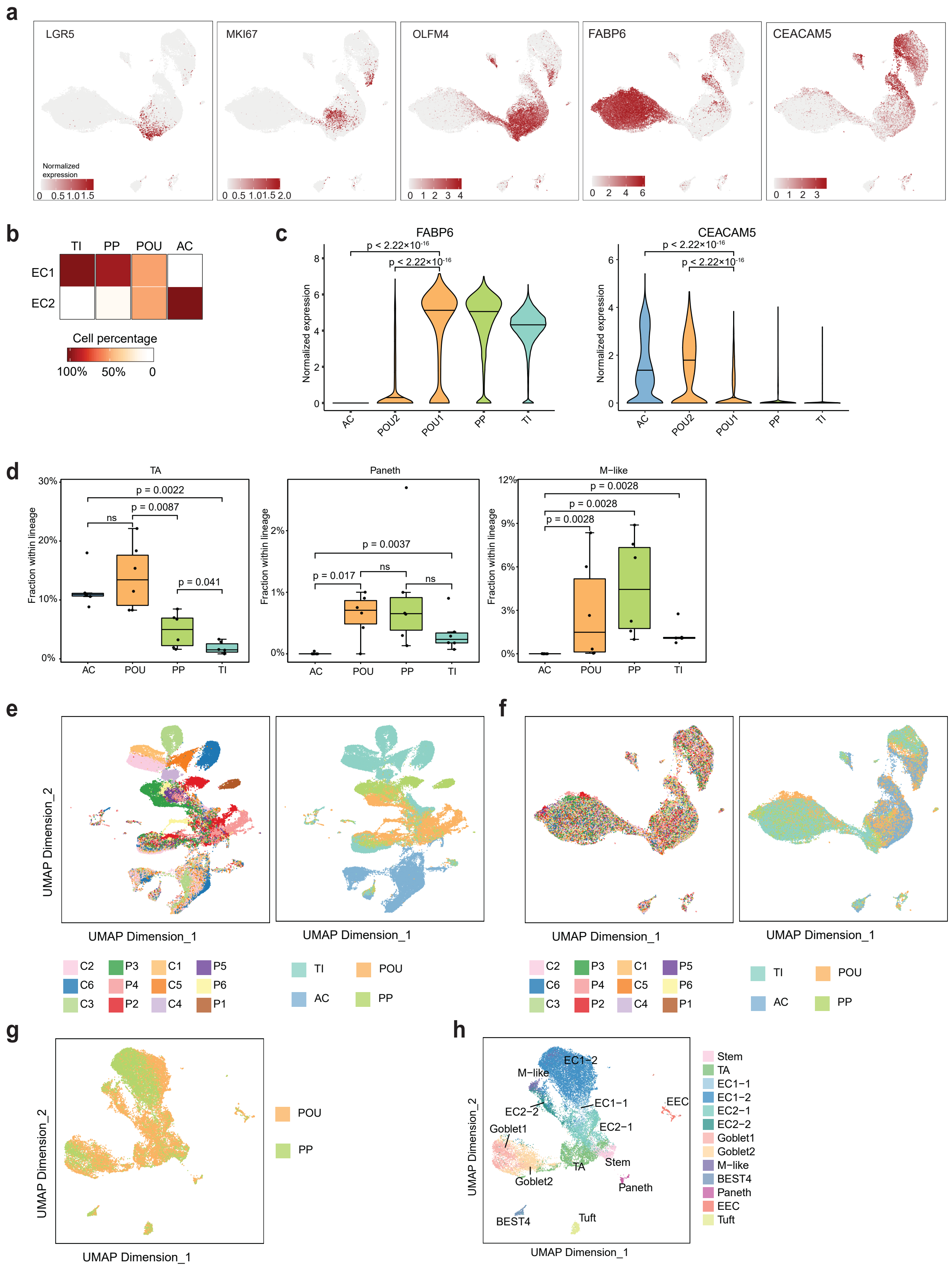

Supplementary Figure 7

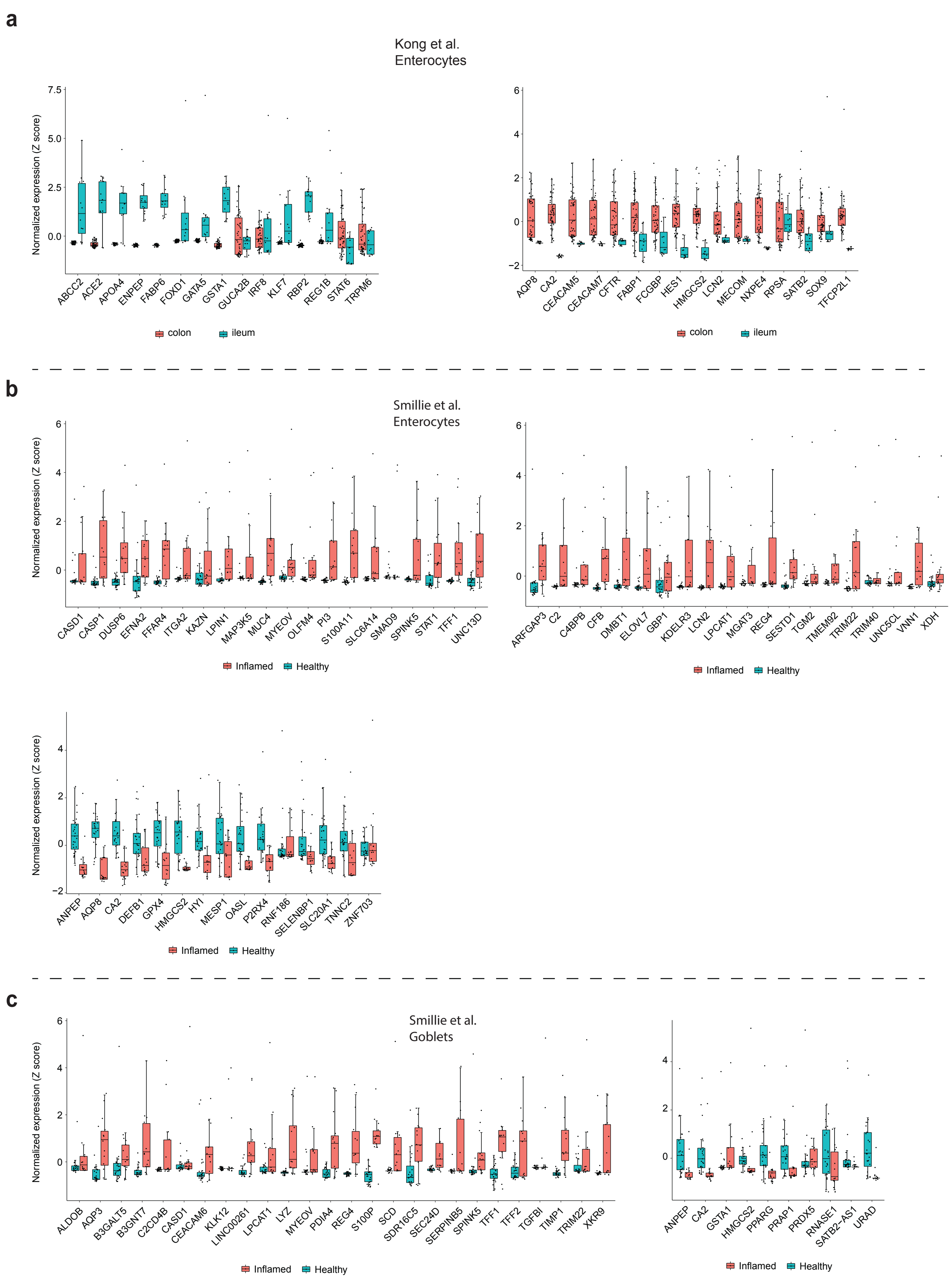

Supplementary Figure 8

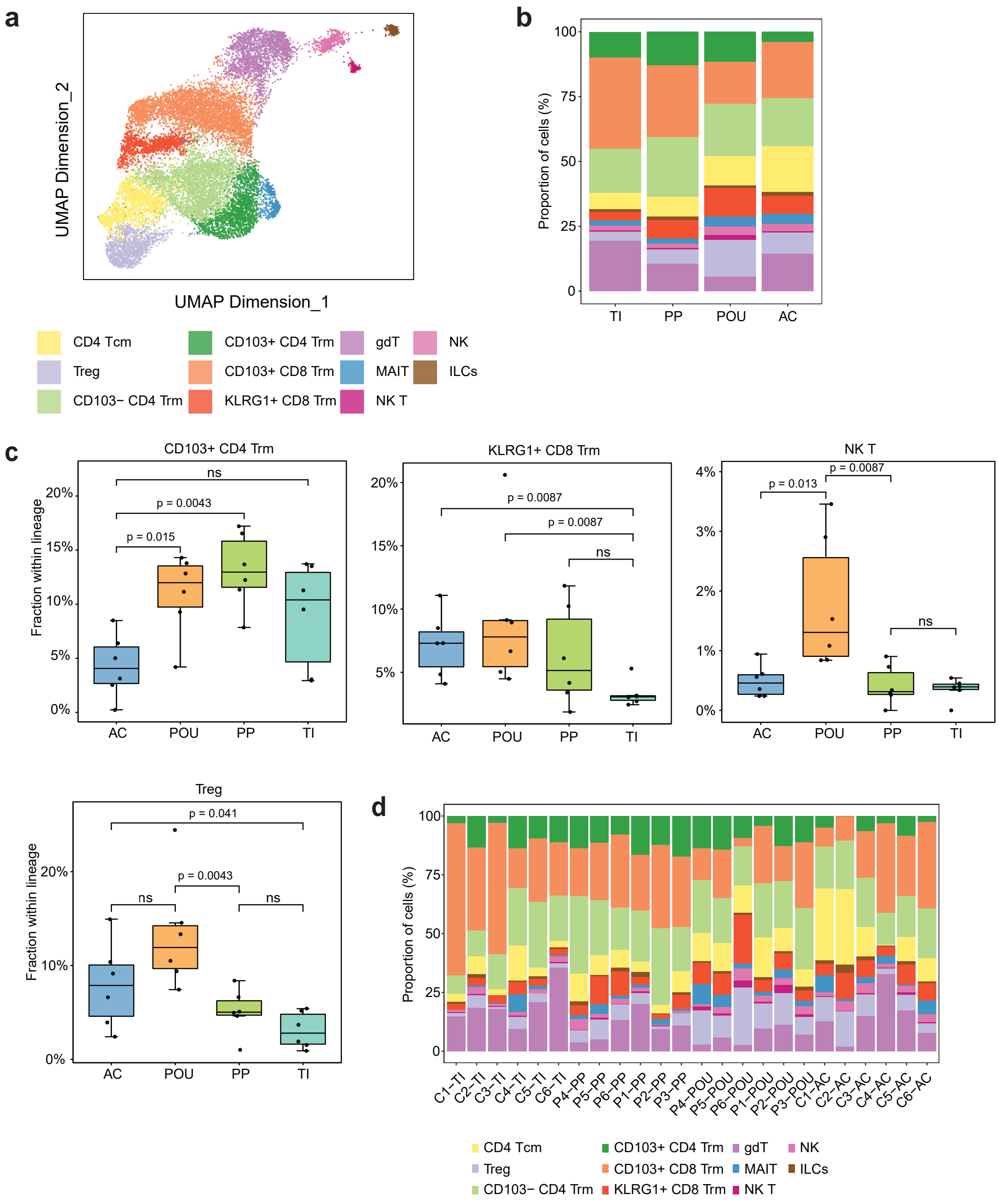

### Supplementary Figure 9

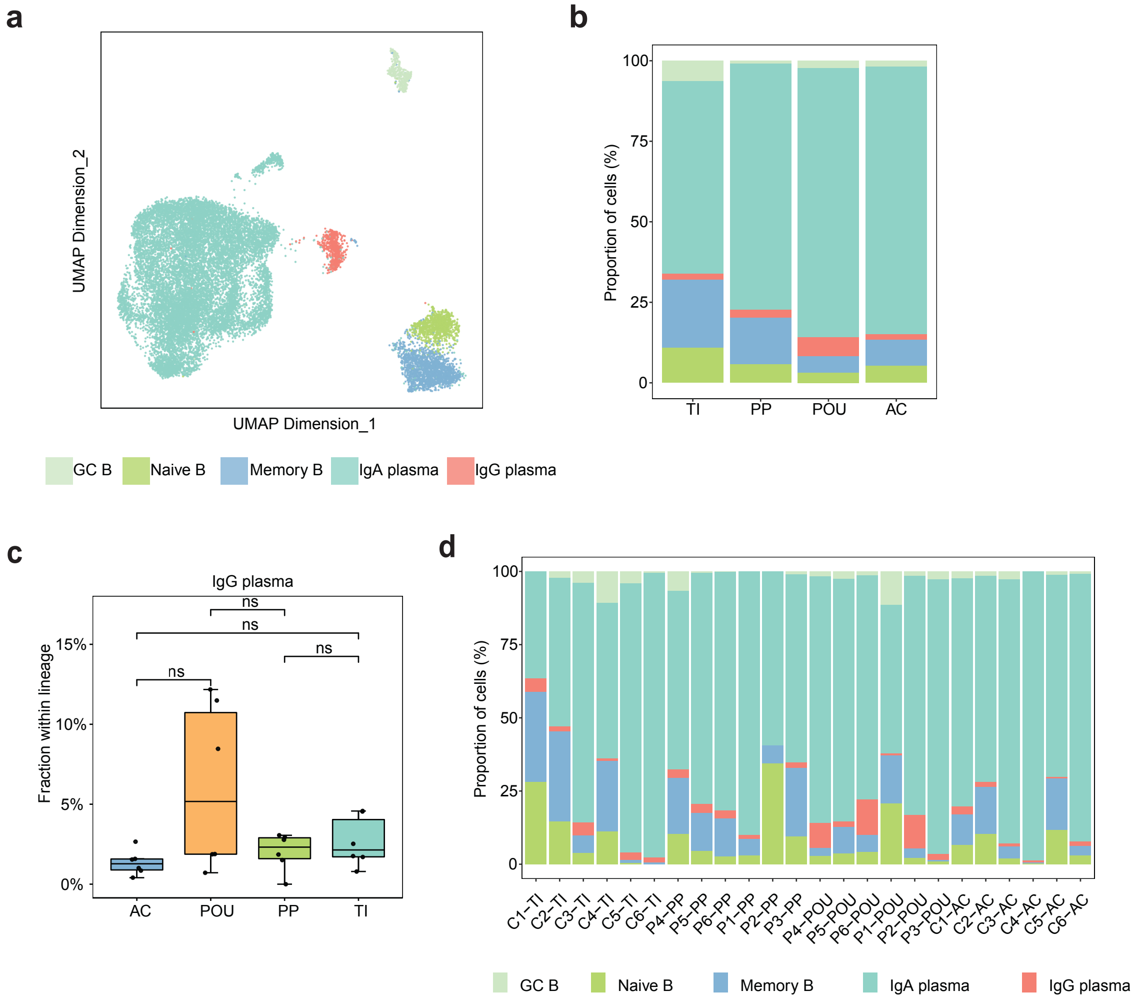

Supplementary Figure 10

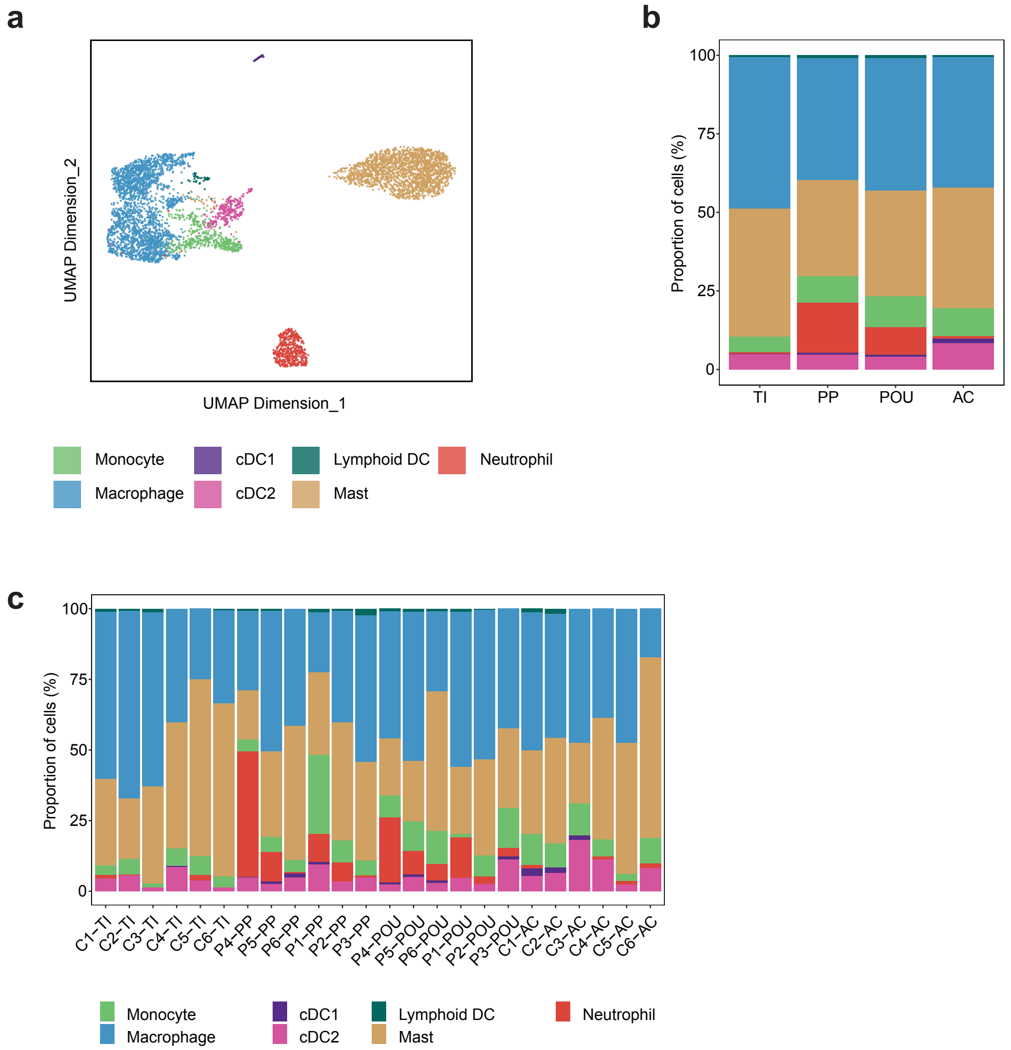

Supplementary Figure 11

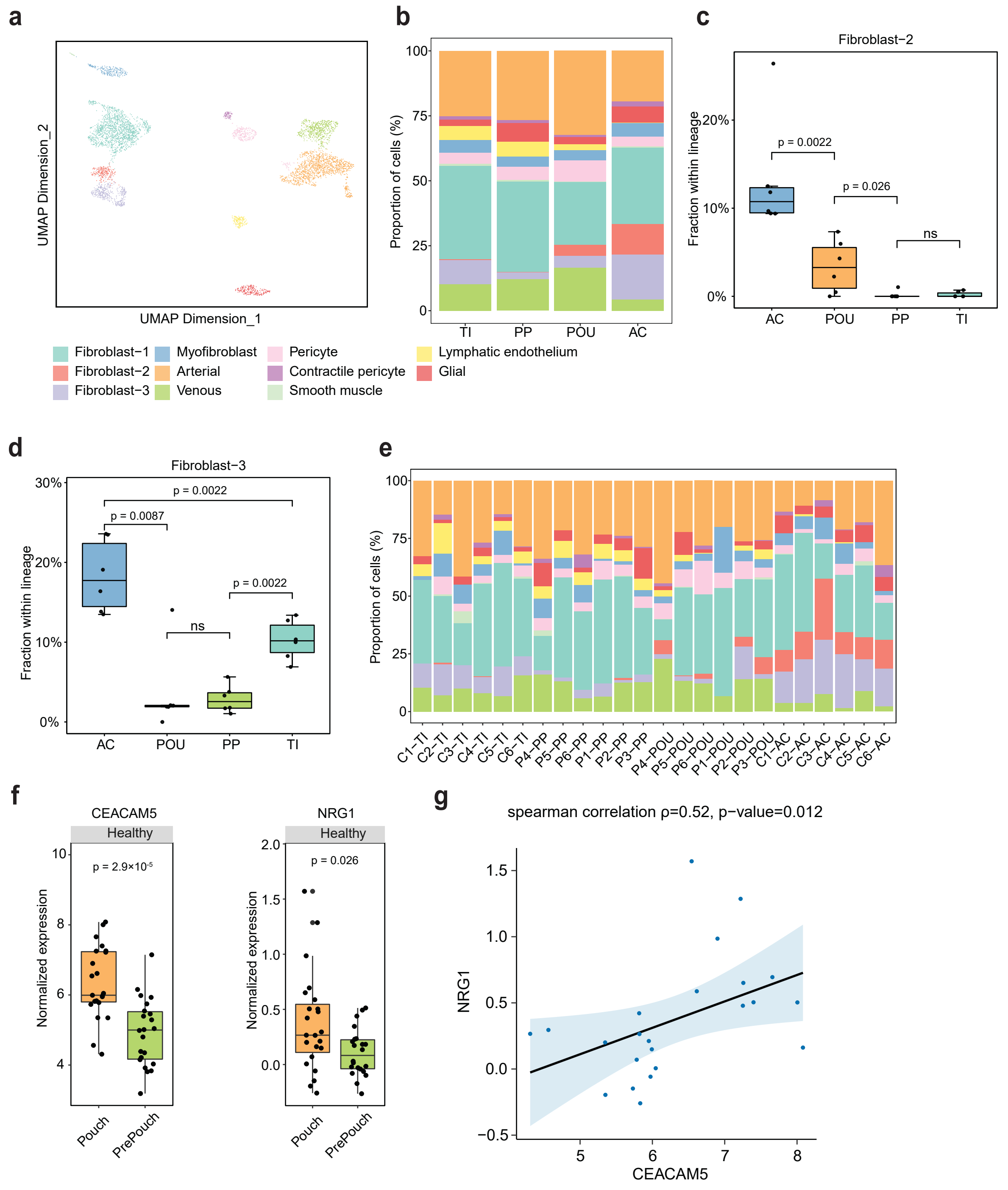

Supplementary Figure 12

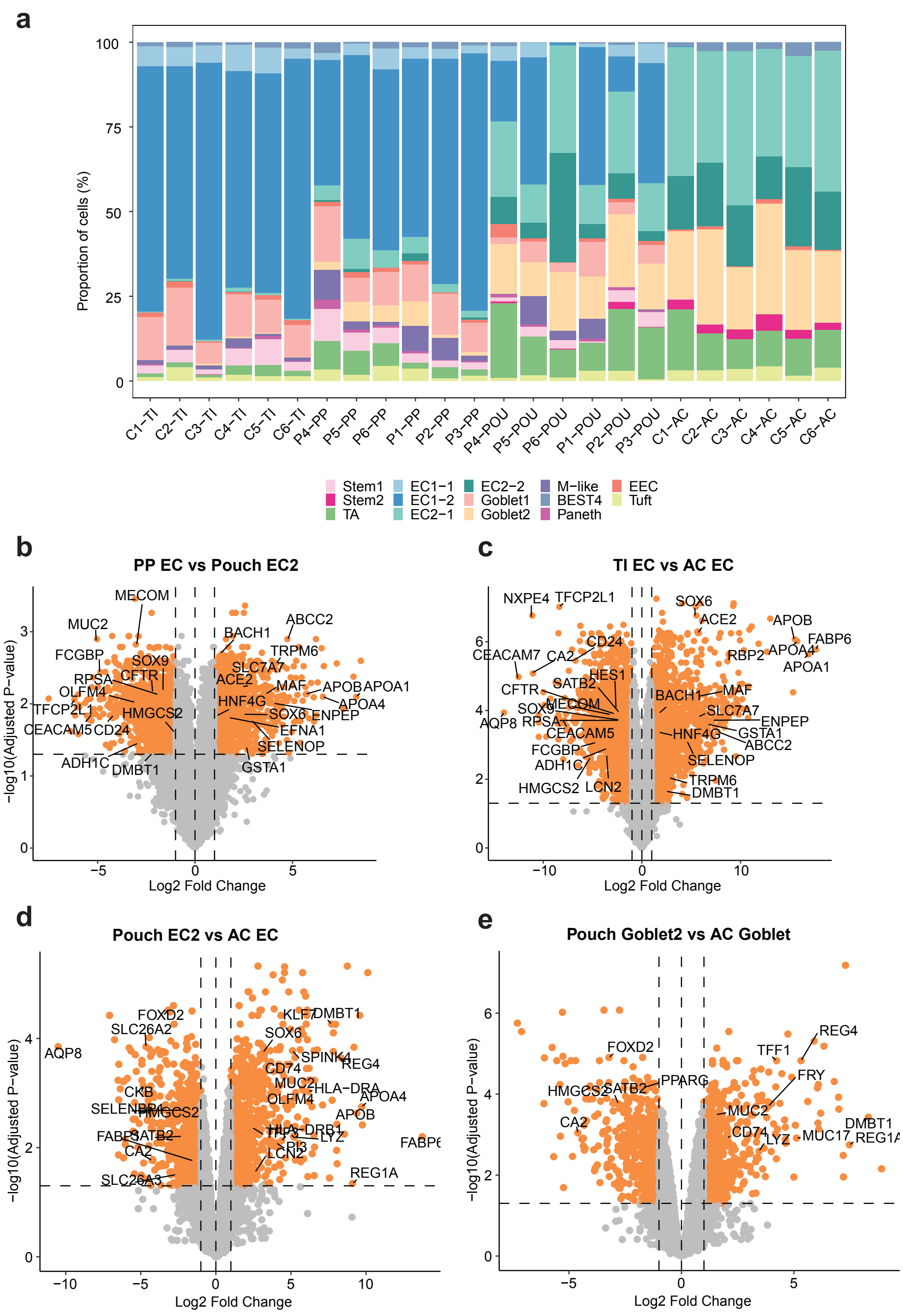

Supplementary Figure 13

**a**

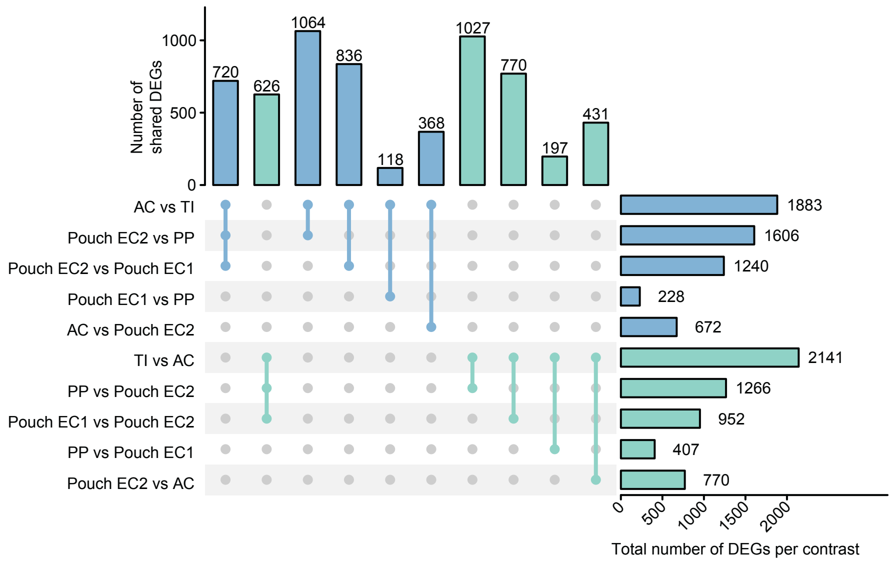


**b**

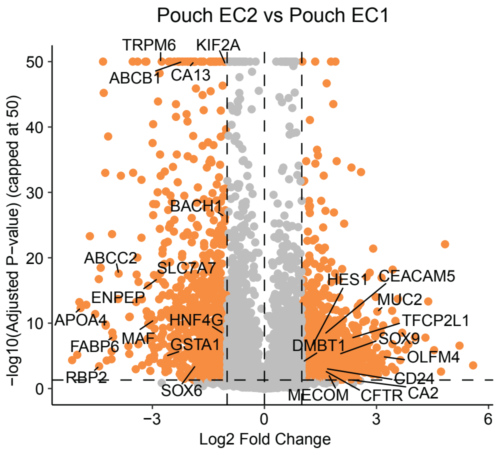

Supplementary Figure 14

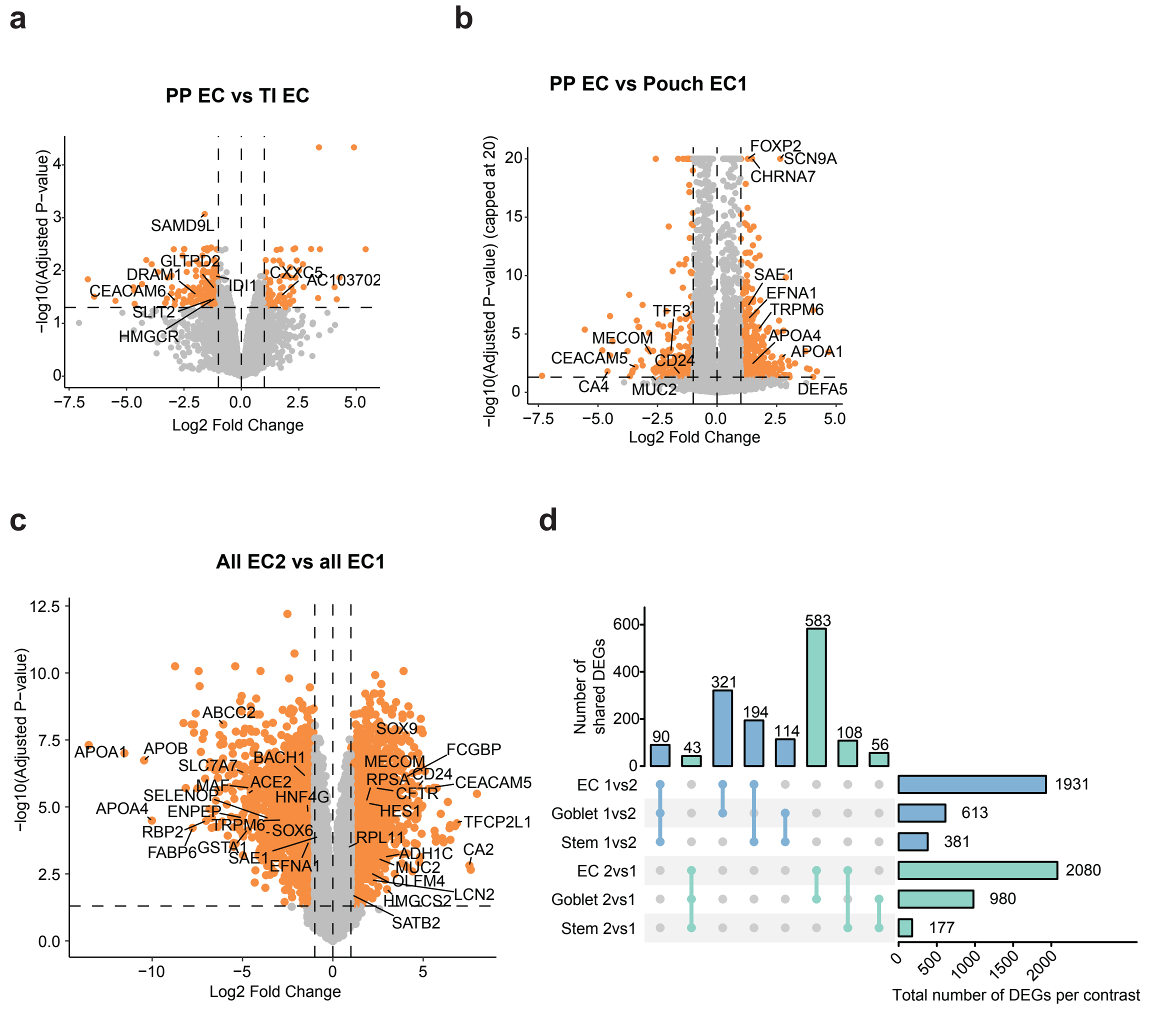

Supplementary Figure 15

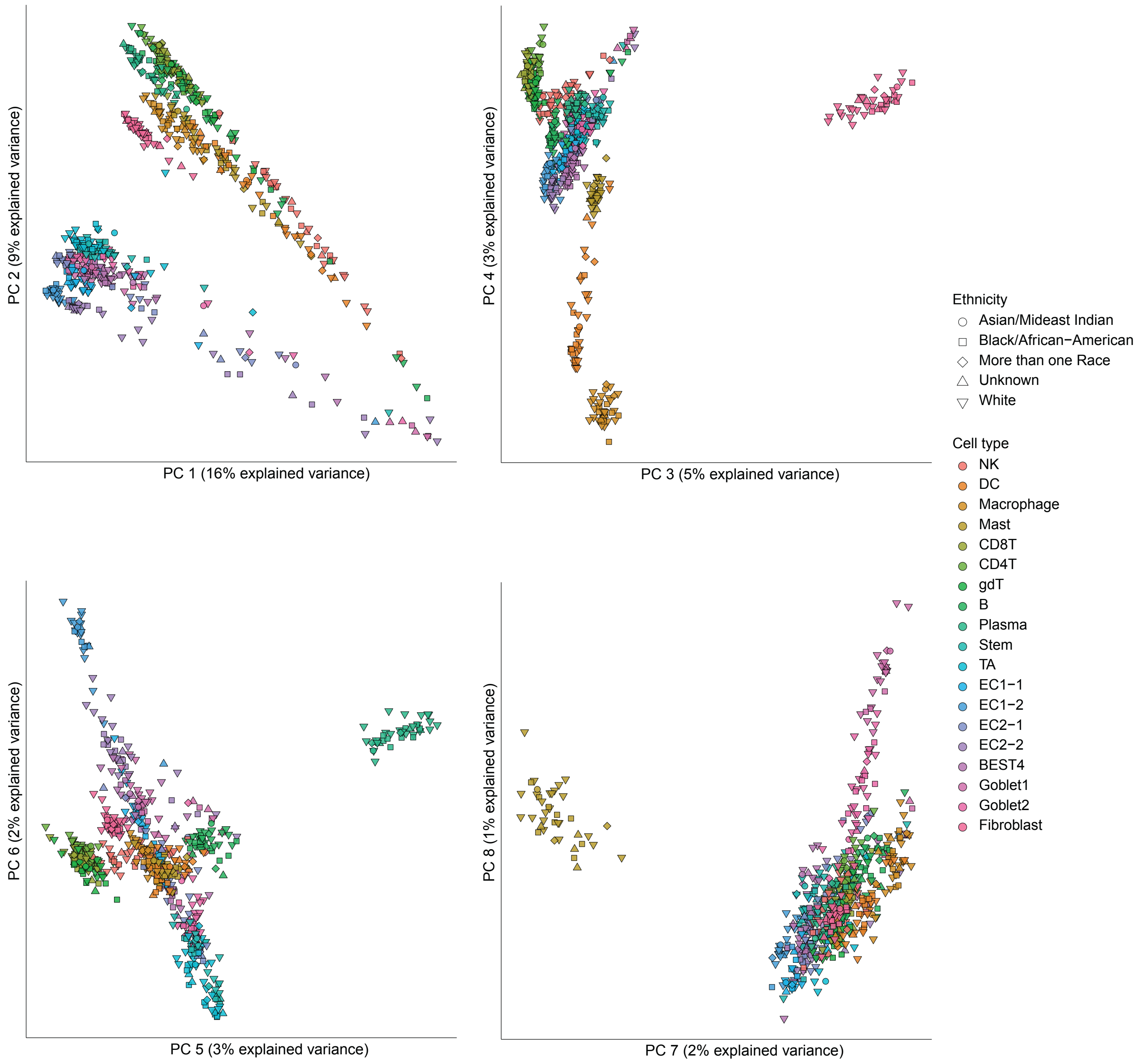

### Supplementary Figure 16

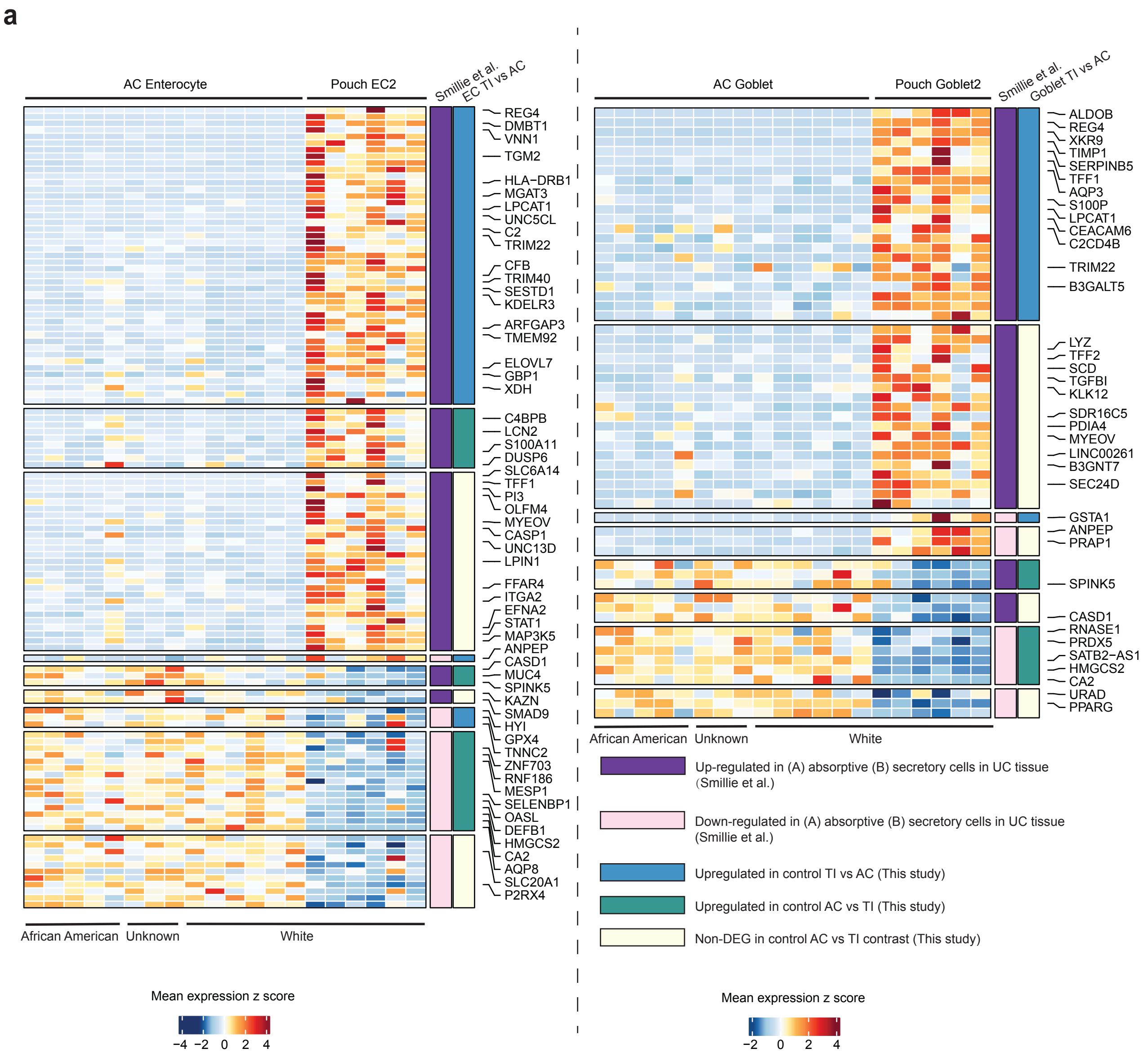


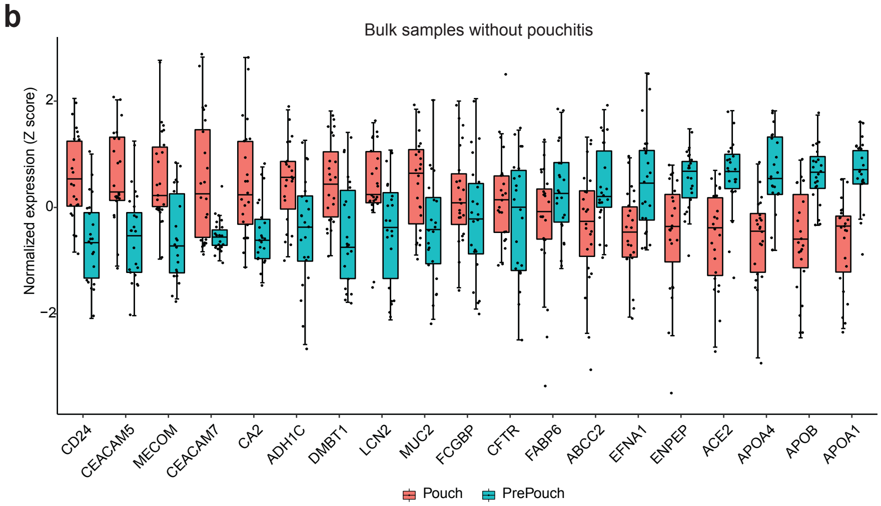

Supplementary Figure 17

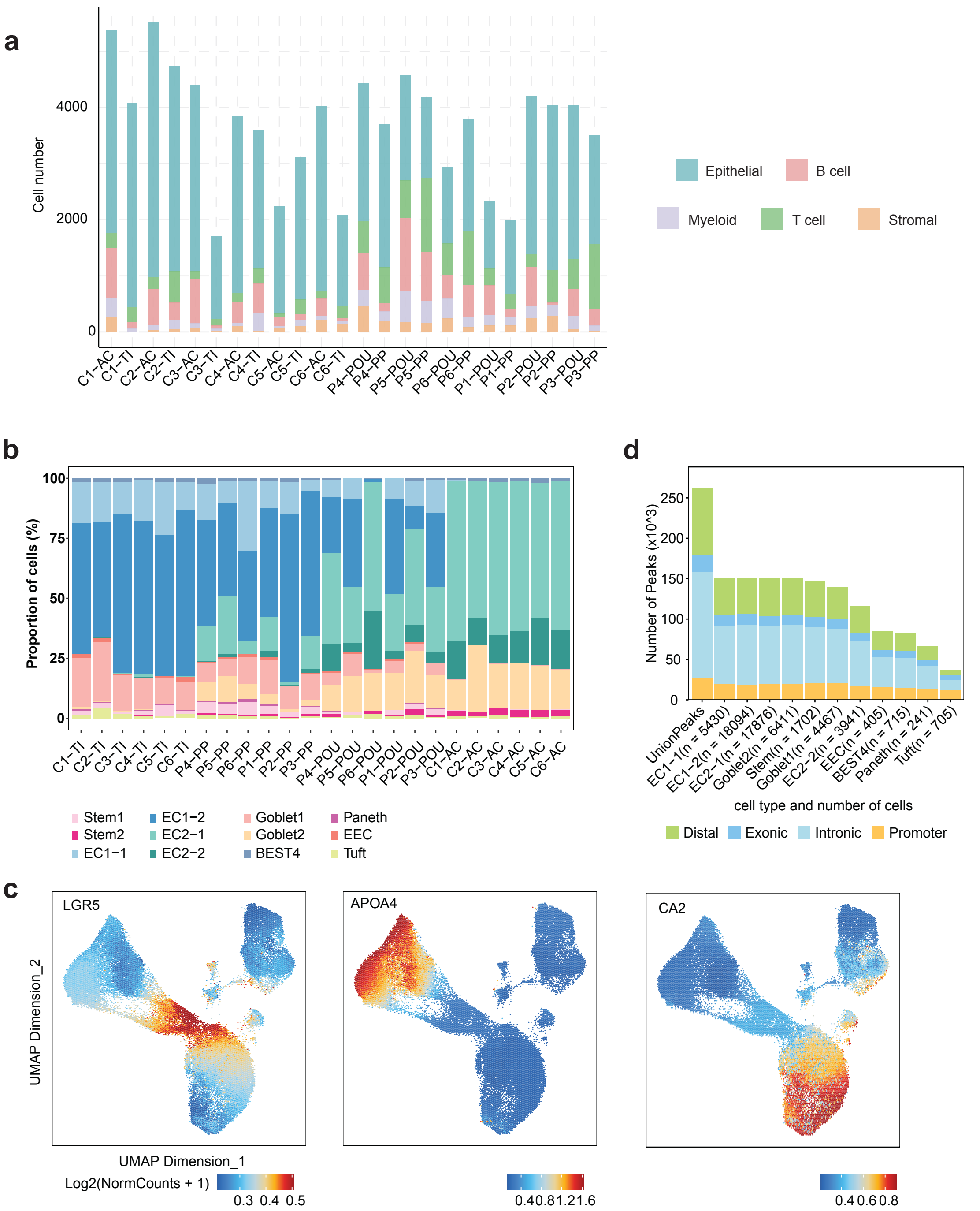

Supplementary Figure 18

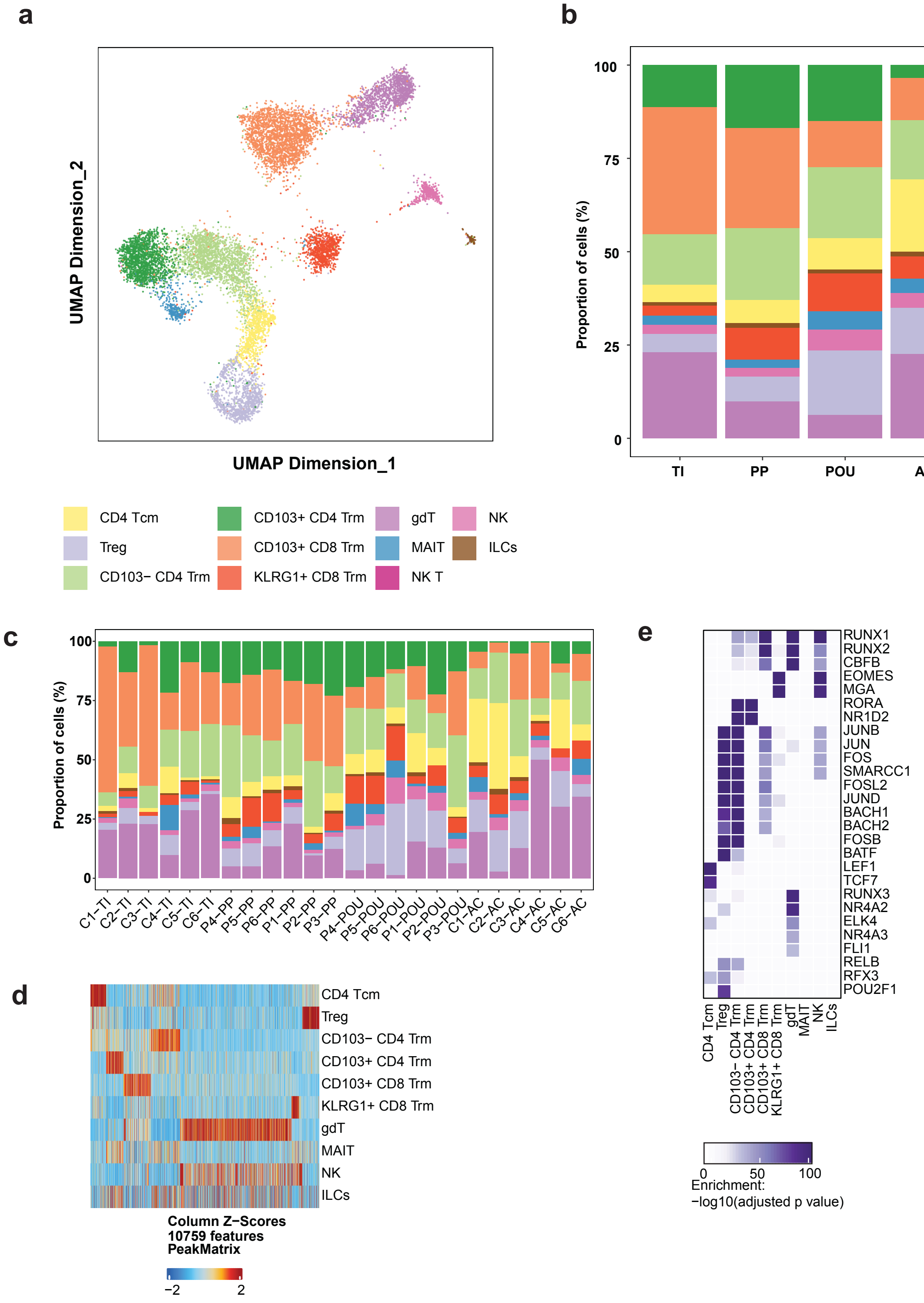

Supplementary Figure 19

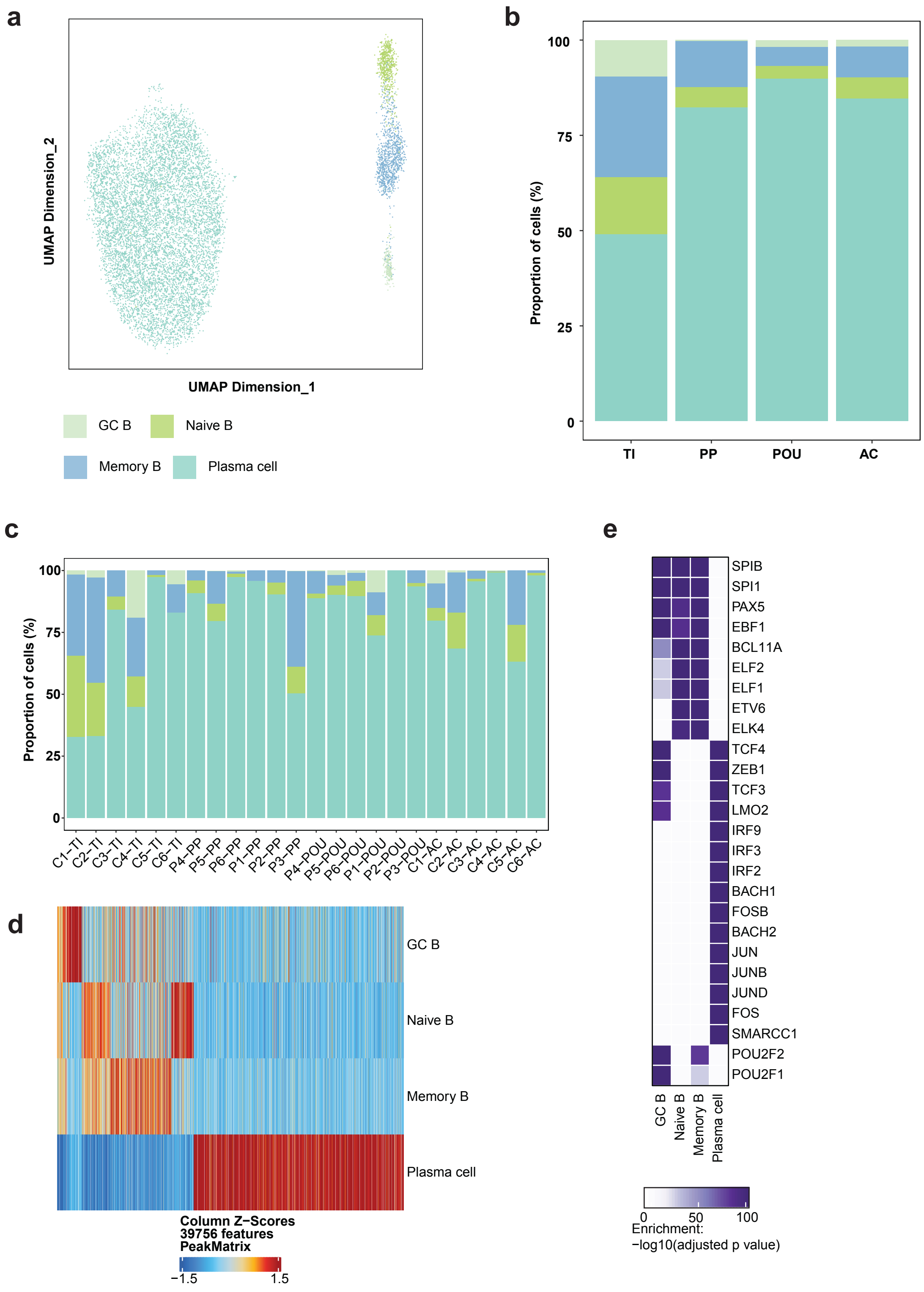

Supplementary Figure 20

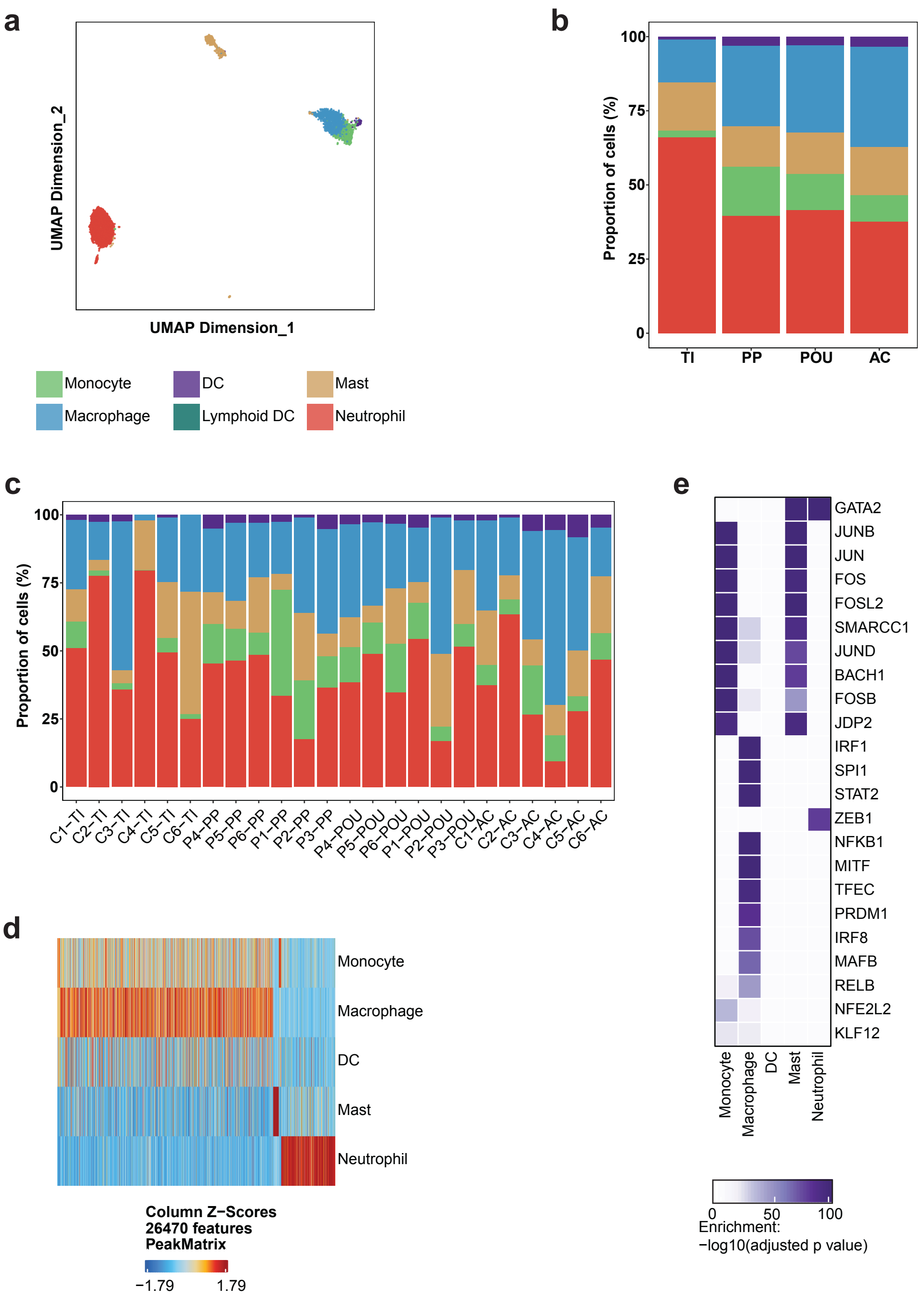

Supplementary Figure 21

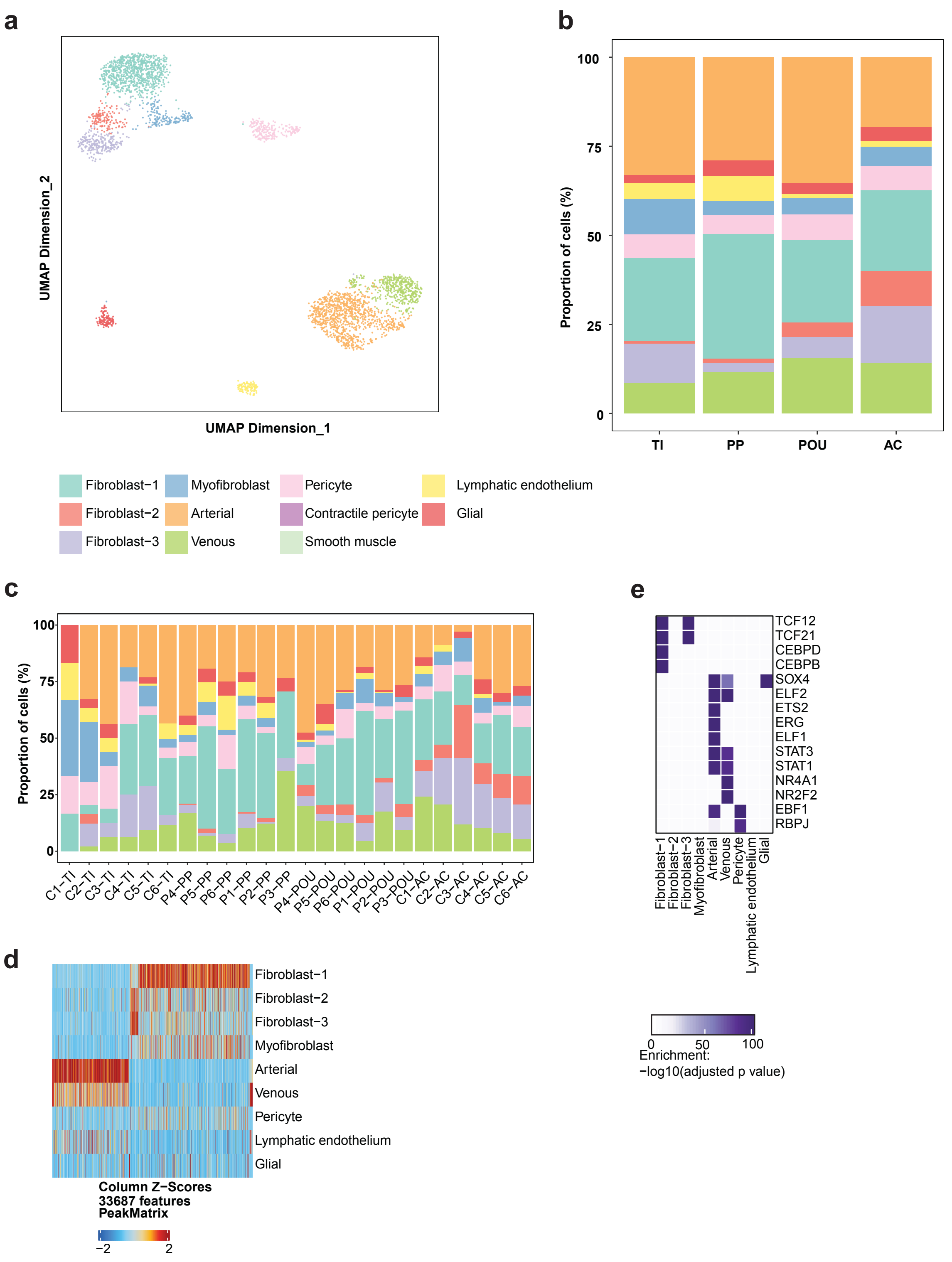

Supplementary Figure 22

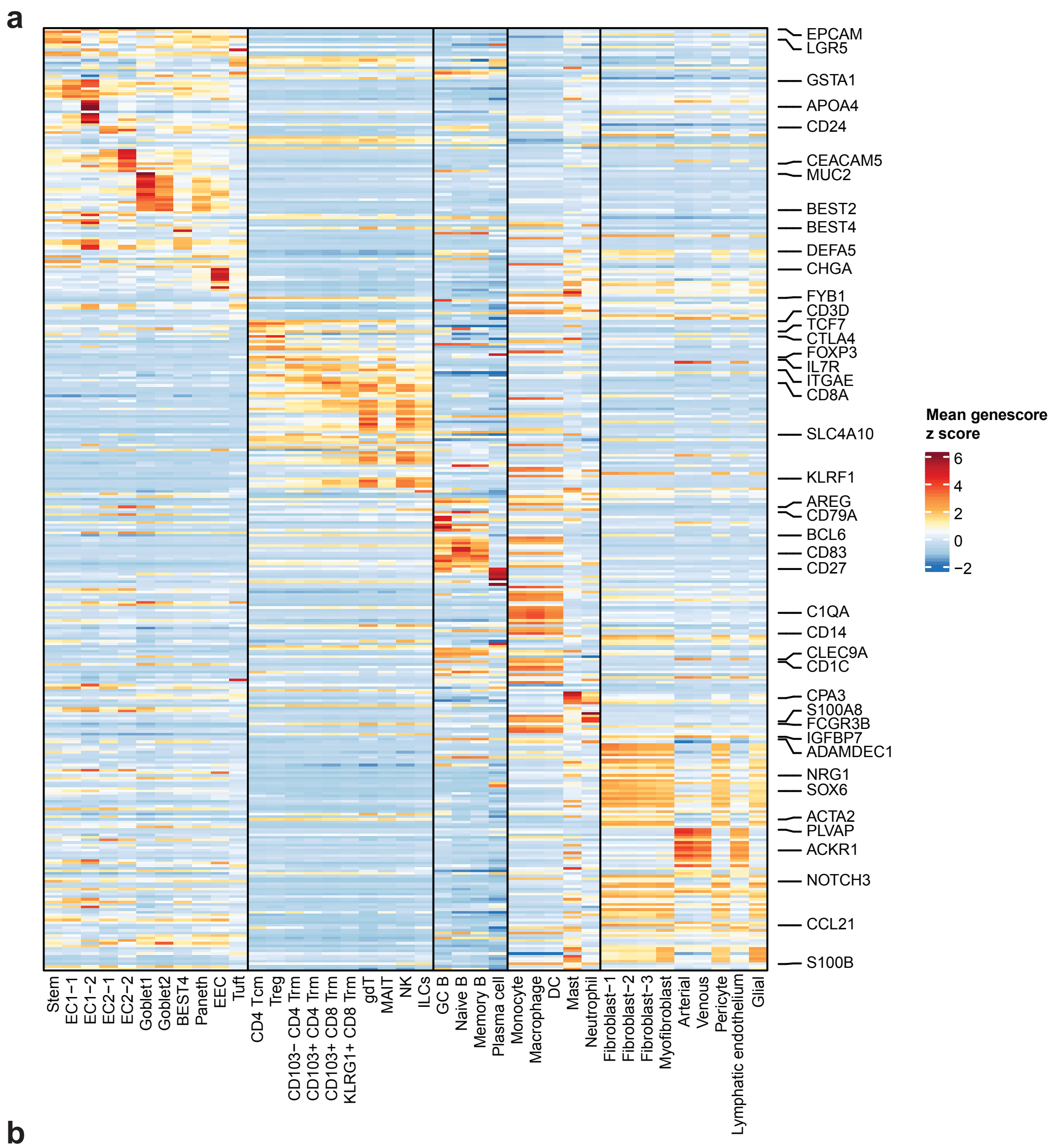


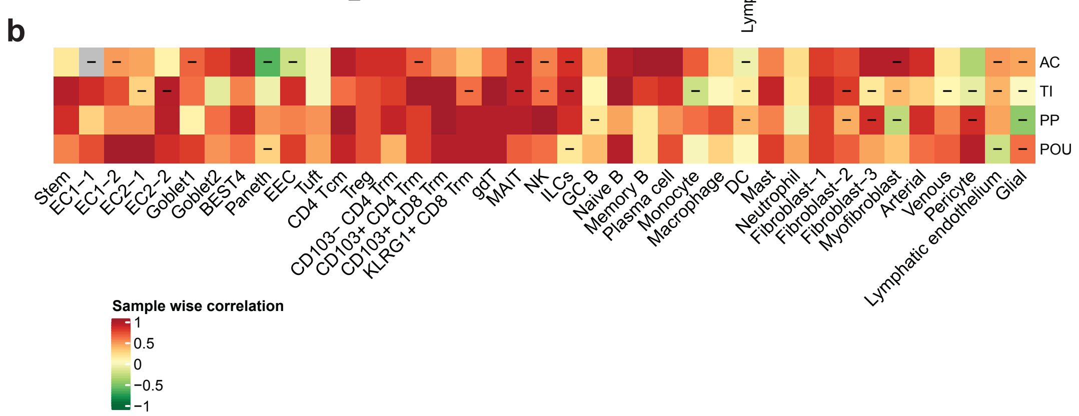

### Supplementary Figure 23

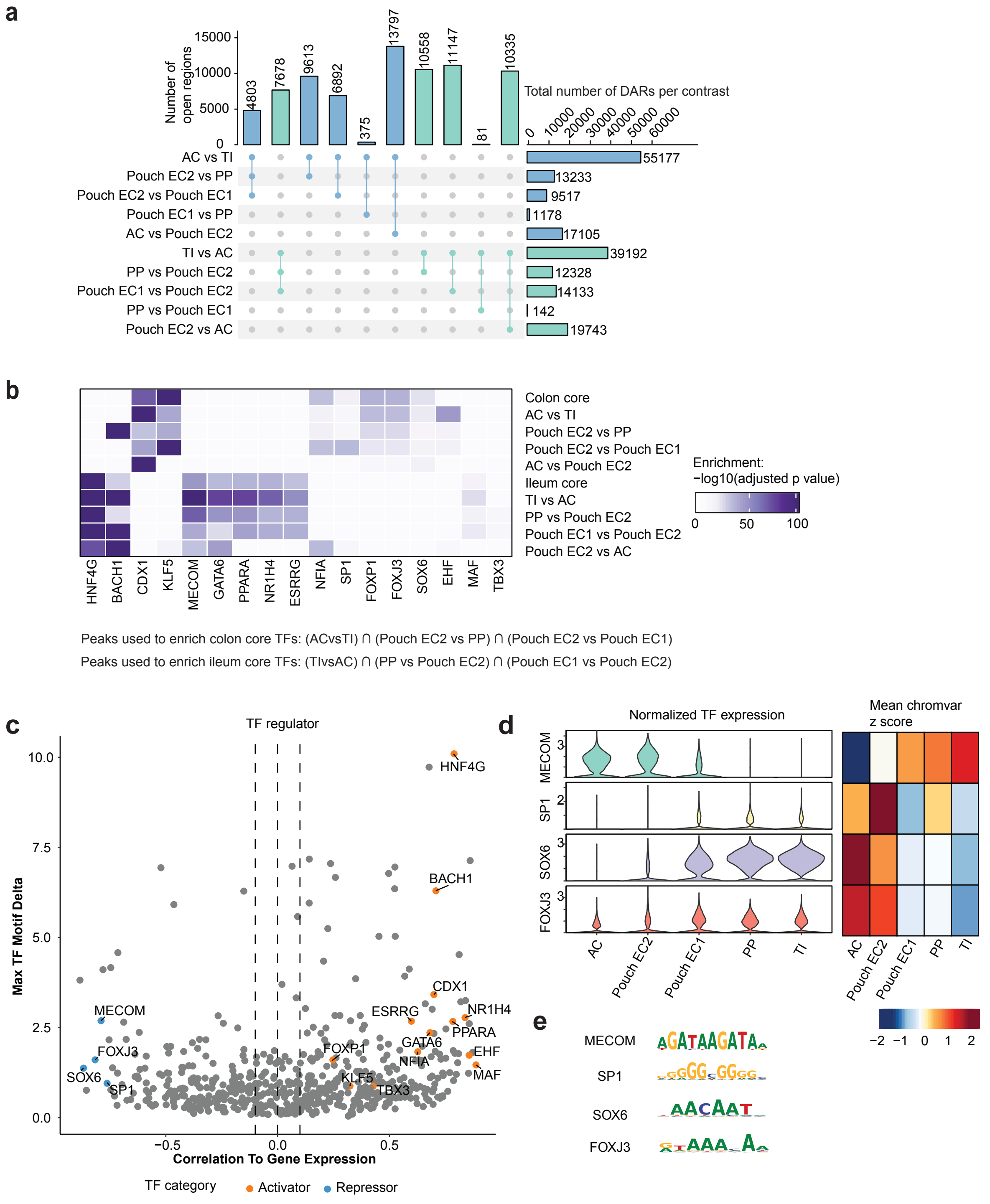

### Supplementary Figure 24

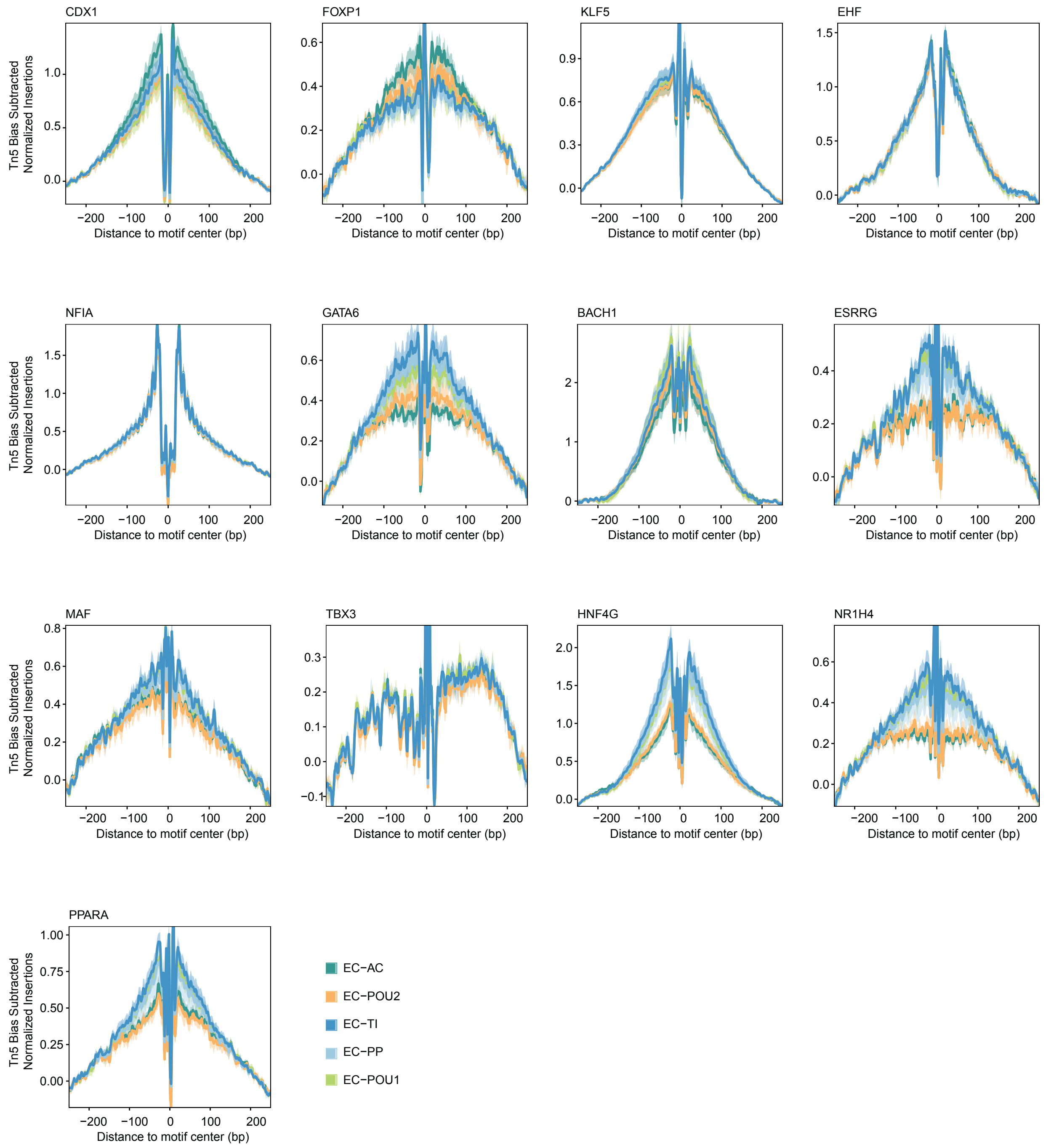

### Supplementary Figure 25

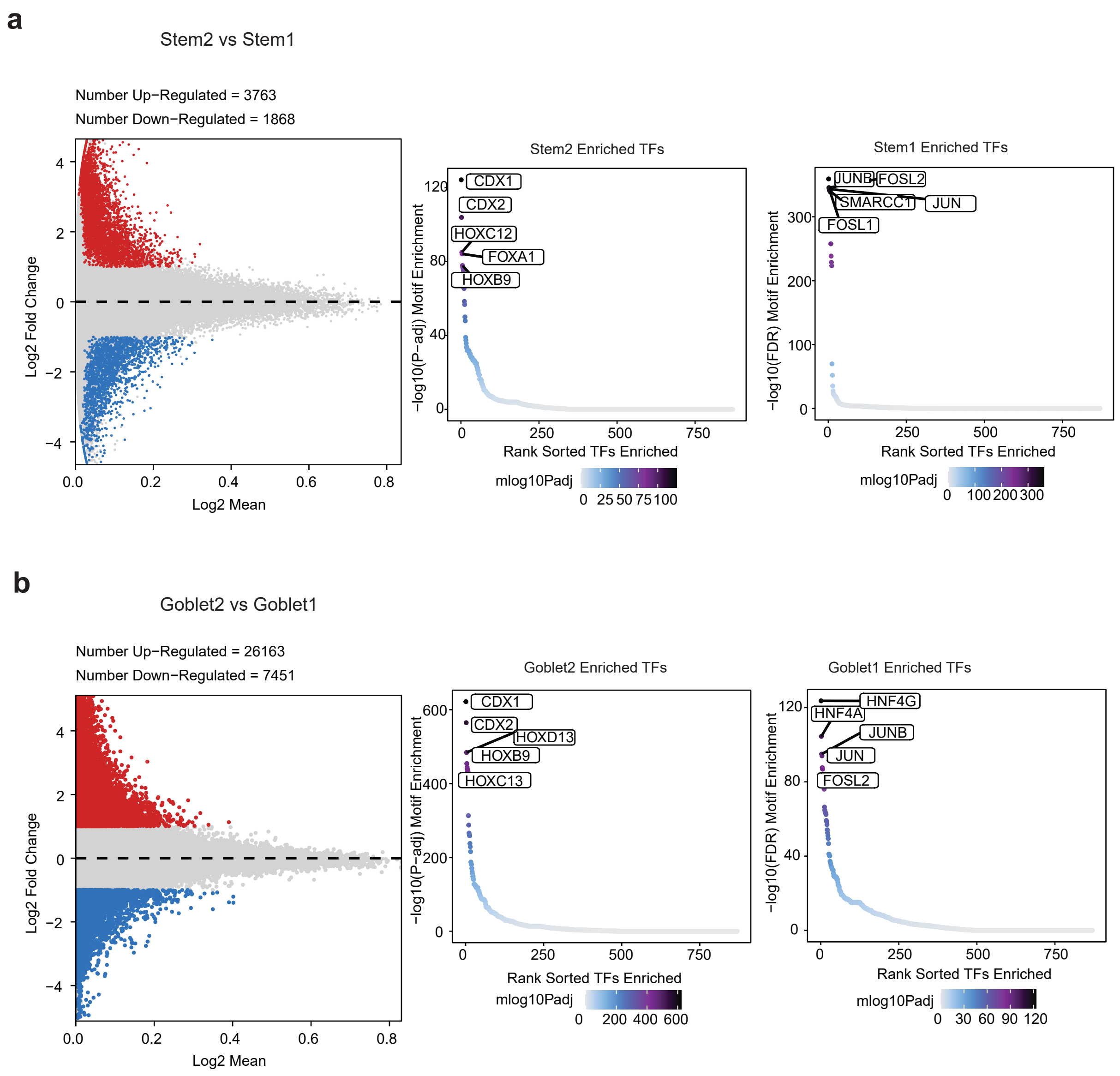

Supplementary Figure 26

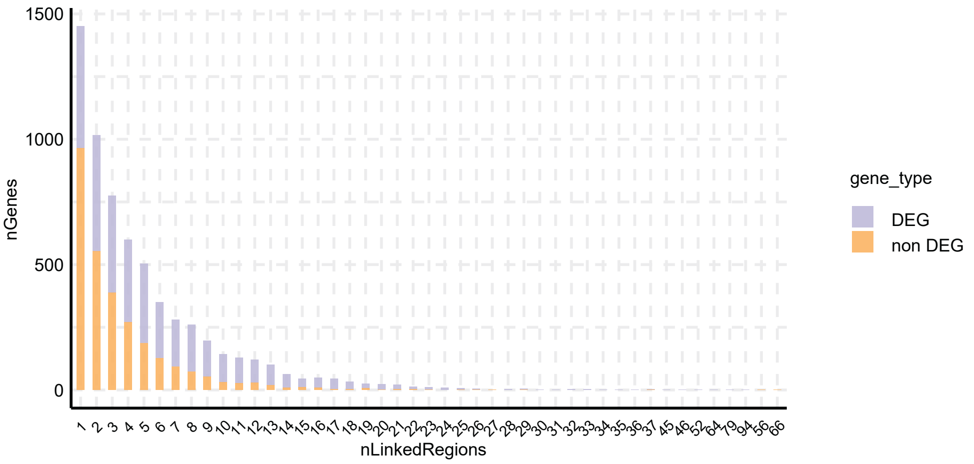

Supplementary Figure 27

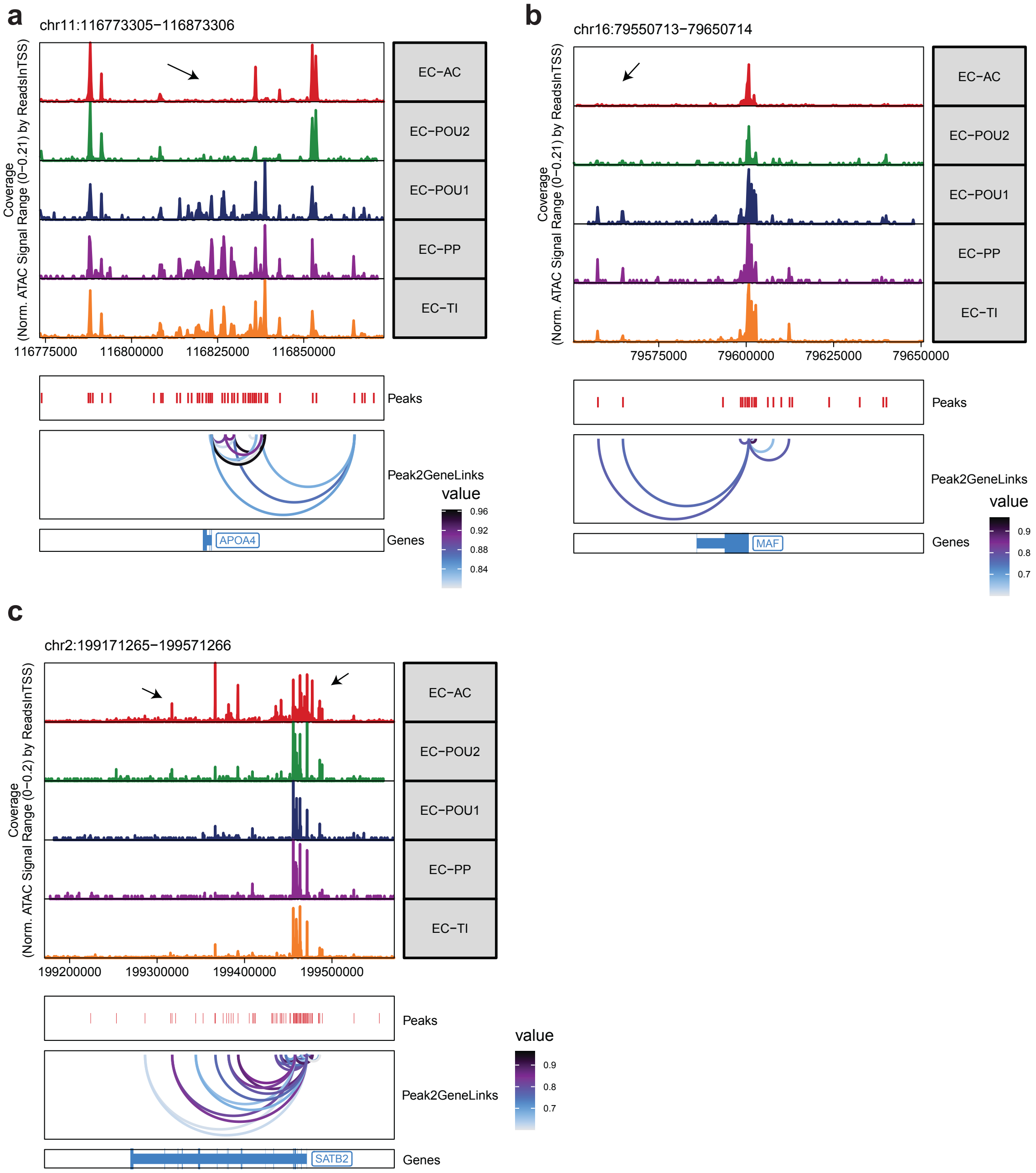
