## Supplementary material for "Multiomic analysis reveals cellular, transcriptomic and epigenetic changes in intestinal pouches of ulcerative colitis patients": description of supplementary data files

### Description of Additional Supplementary Files

**Supplementary Data 1**

**Description:** Patient metadata.

**Supplementary Data 2**

**Description:** Spatial distribution of ADAMDEC1 and NRG1 transcripts in ascending colon, terminal ileum, prepouch and pouch biopsies. Color indicates the number of transcripts detected in each segmented cell, with a maximum of three transcripts to maintain a consistent color scale.

**Supplementary Data 3**

**Description:** Comprehensive differential gene expression analysis between different enterocyte populations. A linear mixed model, accounting for patient-specific effects, was used to evaluate both upregulation and downregulation (two sided test) of genes (Methods). P-values were adjusted for multiple comparisons using the false discovery rate (FDR) method.

**Supplementary Data 4**

**Description:** Pathway enrichment analysis. A non-parametric rank-based Gene Set Enrichment Analysis (GSEA) two-sided test was used to evaluate enrichment (Methods). P-values were adjusted for multiple comparisons using the false discovery rate (FDR) method.

**Supplementary Data 5**

**Description:** Differential gene expression analysis performed between pouch EC2 and AC EC, as well as between pouch Goblet2 and AC Goblet cells. A linear mixed model accounting for patient-specific effects was used to assess gene upregulation and downregulation (two-sided test; Methods). P-values were adjusted for multiple comparisons using the false discovery rate (FDR) method. Genes identified as differentially expressed ($FDR < 0.05$, $|logFC| > 1$) in this study were compared with results published in Smillie et al[1].

**Supplementary Data 6**

**Description: a, b** Experiments to titrate antibodies and validate specificity of labeling were performed on ileal or ascending colonic biopsies collected from 2 patients. **c** Immunohistochemistry images of pouch biopsies stained for FABP6, CEACAM5, and CA2, obtained from a patient different from the one shown in Fig. 4. **d** The deconvolved cell type proportions in healthy pre-pouches from bulk RNA-seq data. **e** The deconvolved cell type proportions in inflammatory prepouches from bulk RNA-seq data.

**Supplementary Data 7-1**

**Description:** **a** Spatial domains in pouch and prepouch replicates from the same donor. **b** Heatmap to show the marker gene expression of each spatial domain. Each column represents a single cell.

**Supplementary Data 7-2**

**Description:** **a, b** Differential gene expression analysis between pouch and prepouch in epithelial domains. Positive fold change indicates gene up-regulation in epithelial domains of pouch samples. Differential expression was assessed using a two-sided Wilcoxon rank-sum test, with p-values adjusted for multiple comparisons using the Bonferroni correction.

**Supplementary Data 7-3**

**Description:** Spatial distribution of CEACAM5, CD24, CA2, FABP6 and APOA4 transcripts in ascending colon, terminal ileum, prepouch and pouch biopsies. Color indicates the number of transcripts detected in each segmented cell, with a maximum of three transcripts to maintain a consistent color scale.

**Supplementary Data 7-4**

**Description:** Spatial distribution of CEACAM5, CD24, CA2, FABP6 and APOA4 transcripts in prepouch and pouch biopsies. Color indicates the number of transcripts detected in each segmented cell, with a maximum of three transcripts to maintain a consistent color scale.

**Supplementary Data 8-1**

**Description:** Spatial distribution of CDX1, FOXP1, KLF5, EHF, NFIA and BACH1 transcripts in prepouch and pouch biopsies.

**Supplementary Data 8-2**

**Description:** Spatial distribution of ESRRG, MAF, TBX3, HNF4G, NR1H4 and PPARA transcripts in prepouch and pouch biopsies.

**Supplementary Data 9-1**

**Description: Multiomic analysis**. **a** UMAP of enterocytes and stem cells in pouch. **b** Pseudotime ordering of cells along differentiation from stem cells to EC1 cells. **c** Pseudotime ordering of cells along differentiation from stem cells to EC2 cells. (**d, e**) Gene expression changes along pseudotime in each lineage.

**Supplementary Data 9-2**

**Description: Multiomic analysis. a** UMAP of enterocytes and stem cells in AC and TI. **b** Pseudotime ordering of cells along differentiation from stem cells to EC1 cells. **c** Pseudotime ordering of cells along differentiation from stem cells to EC2 cells. **d** In AC and TI, the TF activity changes in EC1 lineage and EC2 lineage along simulated differentiation pseudotime. **e** Z scores of average gene expression from bulk RNA-seq data of healthy pouches (Huang et al.[2]) were plotted to confirm the transcription factor expression differences between the pouch and prepouch enterocytes identified in our study.

**Supplementary Data 10**

**Description:** Domain of regulatory chromatin (DORC) gene analysis.

**Supplementary Data 11**

**Description:** Additional probes for Xenium experiments.

**Supplementary Data 12**

**Description:** Accession codes for the raw data of each sample.
