## Supplementary figures and images for "Multiomic analysis reveals cellular, transcriptomic and epigenetic changes in intestinal pouches of ulcerative colitis patients"

### Supplementary Data 2

#### Supplementary Data 2

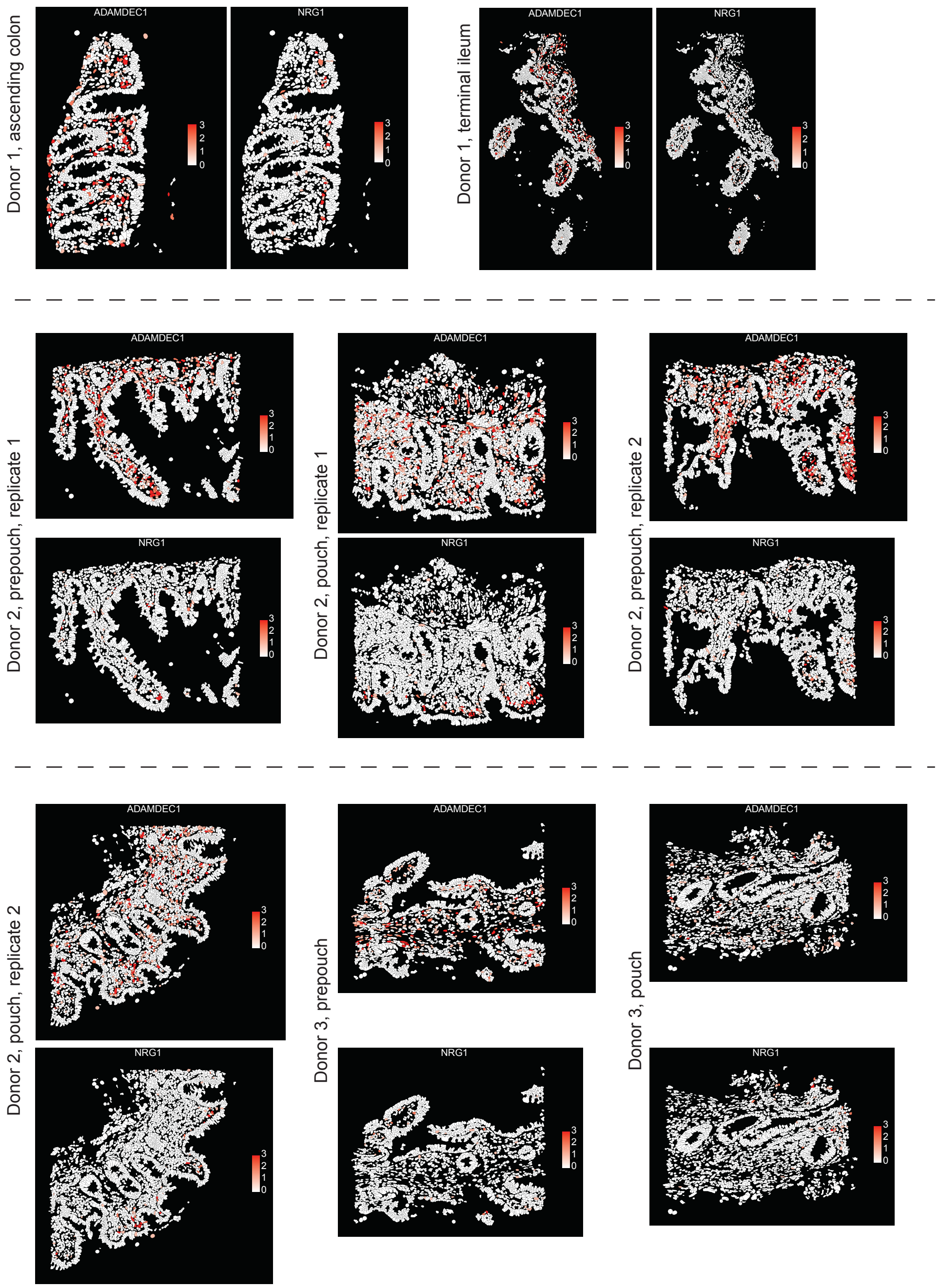

### Supplementary Data 6

Supplementary Data 6

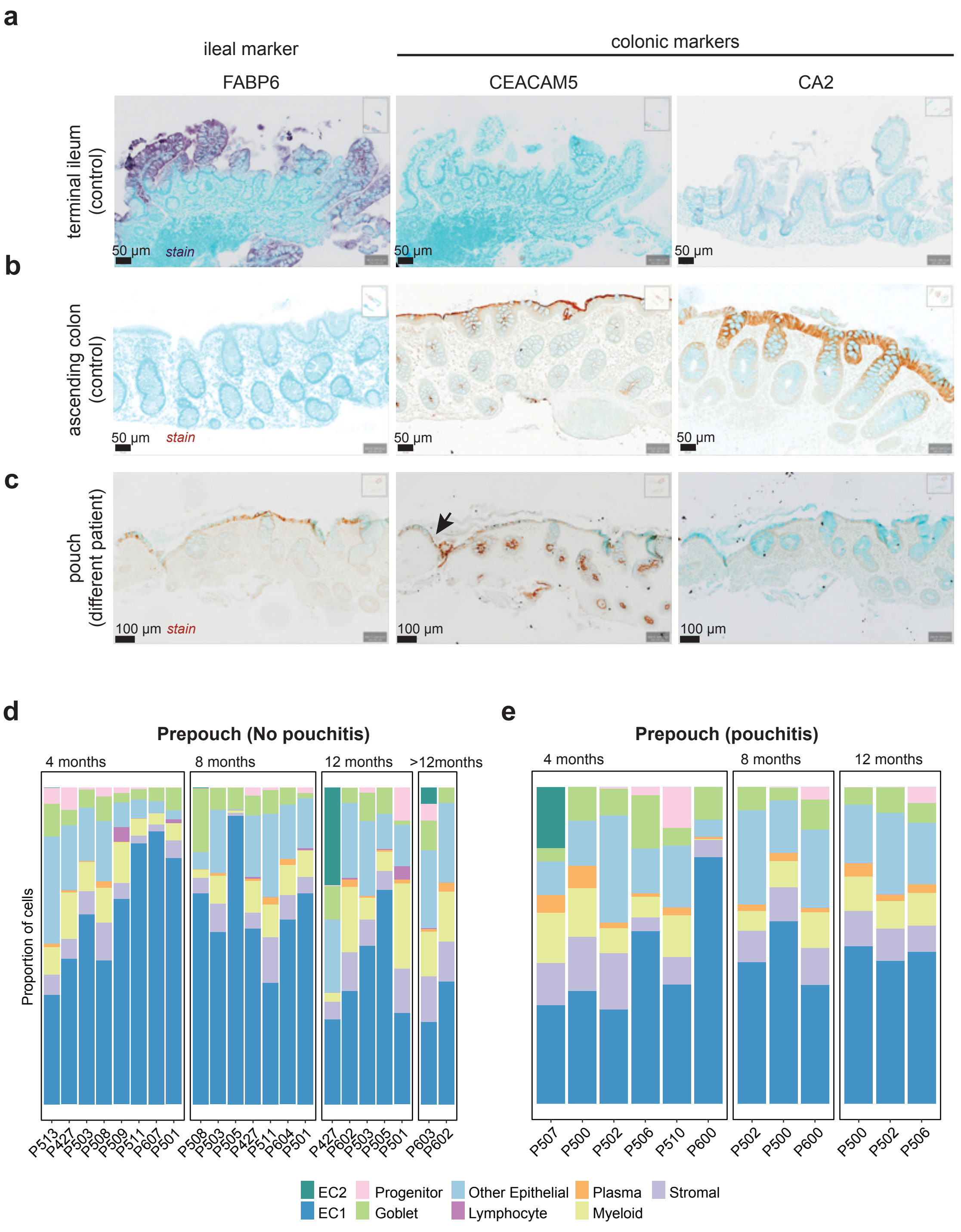

### Supplementary Data 7

Supplementary Data 7-1

**a**

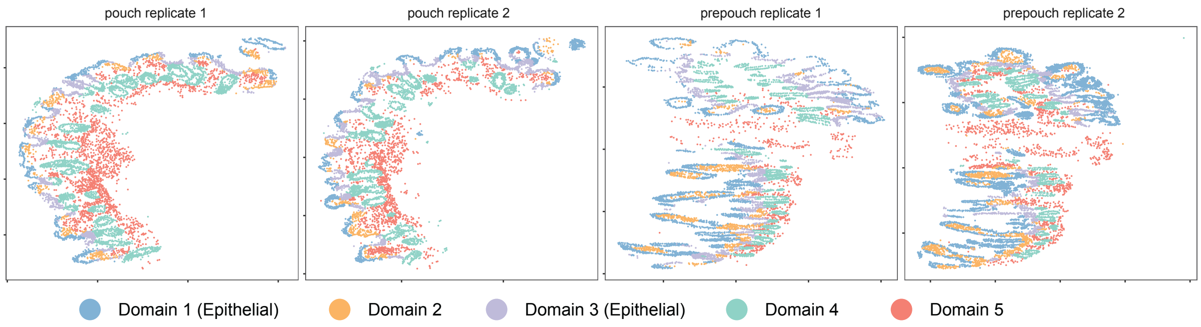


**b**

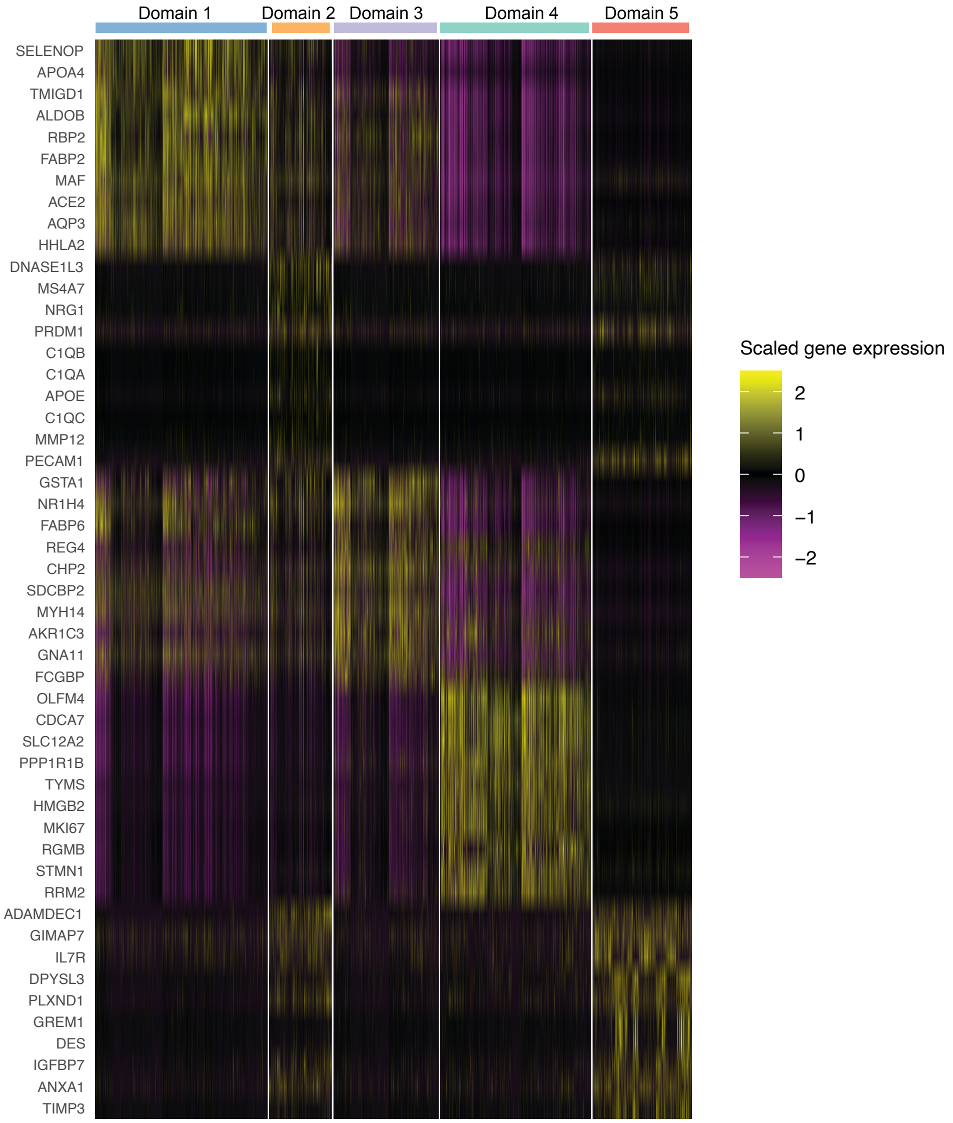

#### Supplementary Data 7-2

**a**

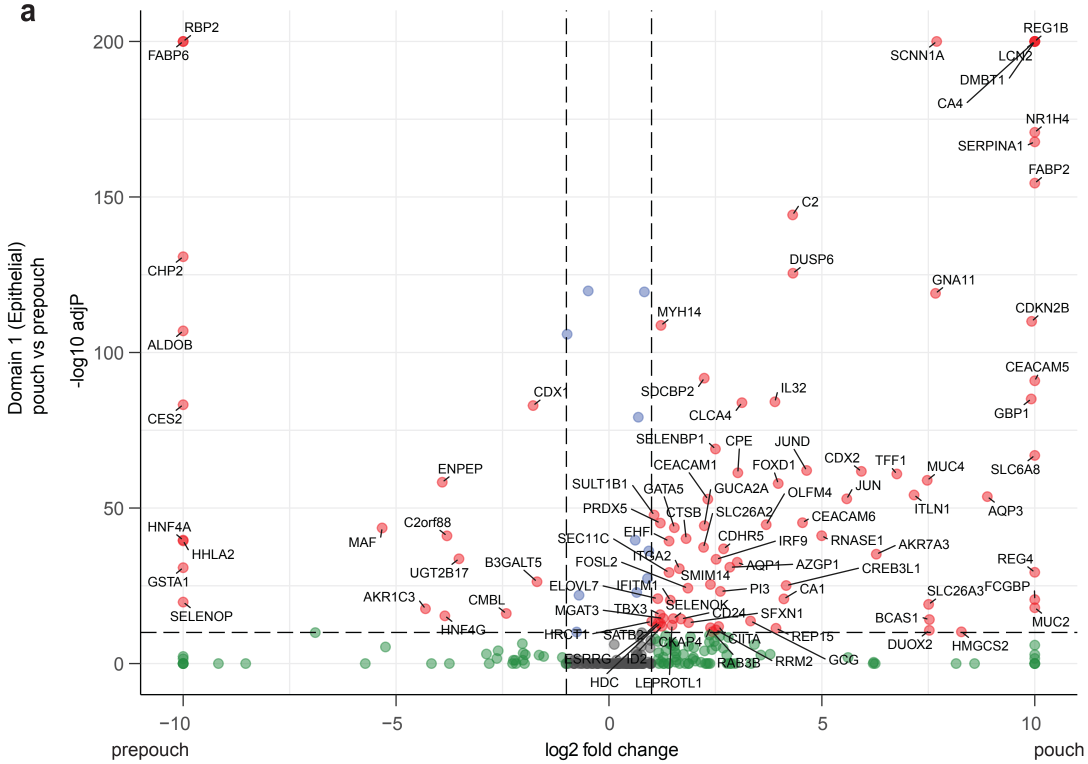


**b**

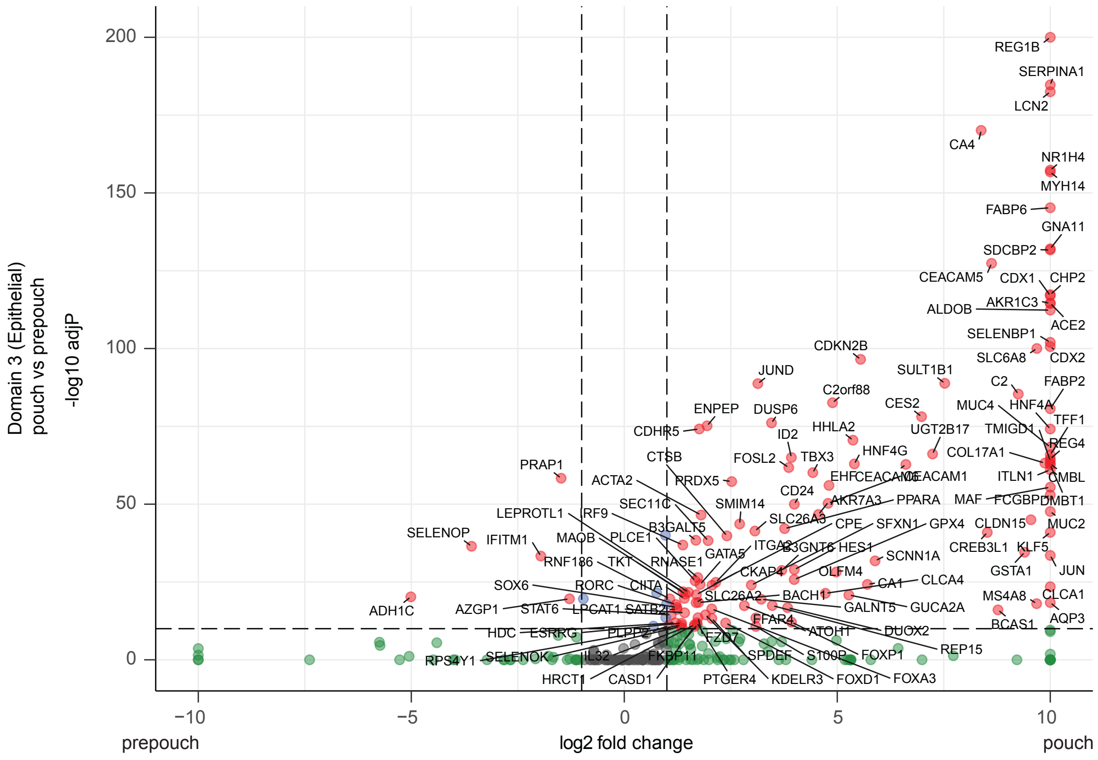

#### Supplementary Data 7-3

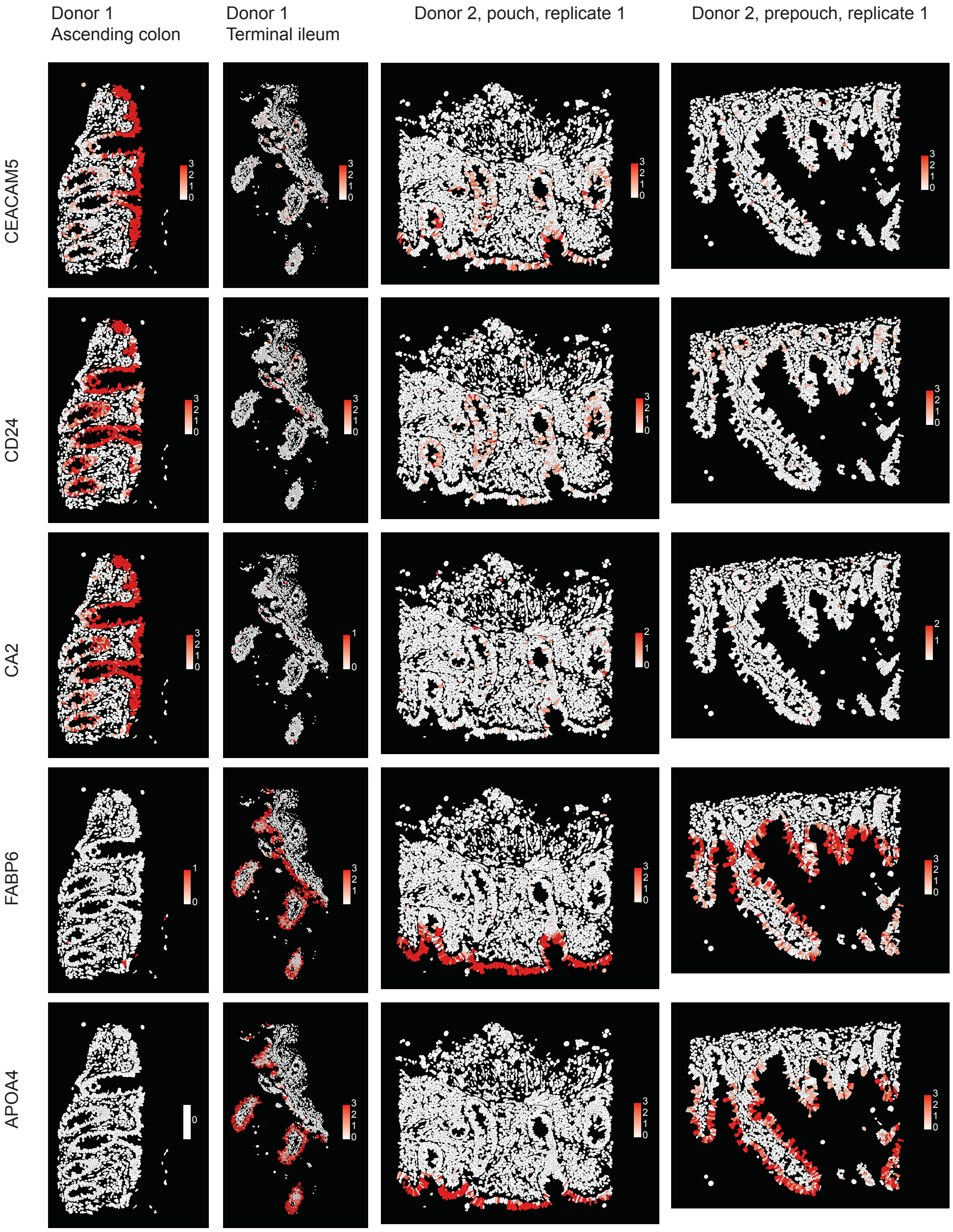

Supplementary Data 7-4

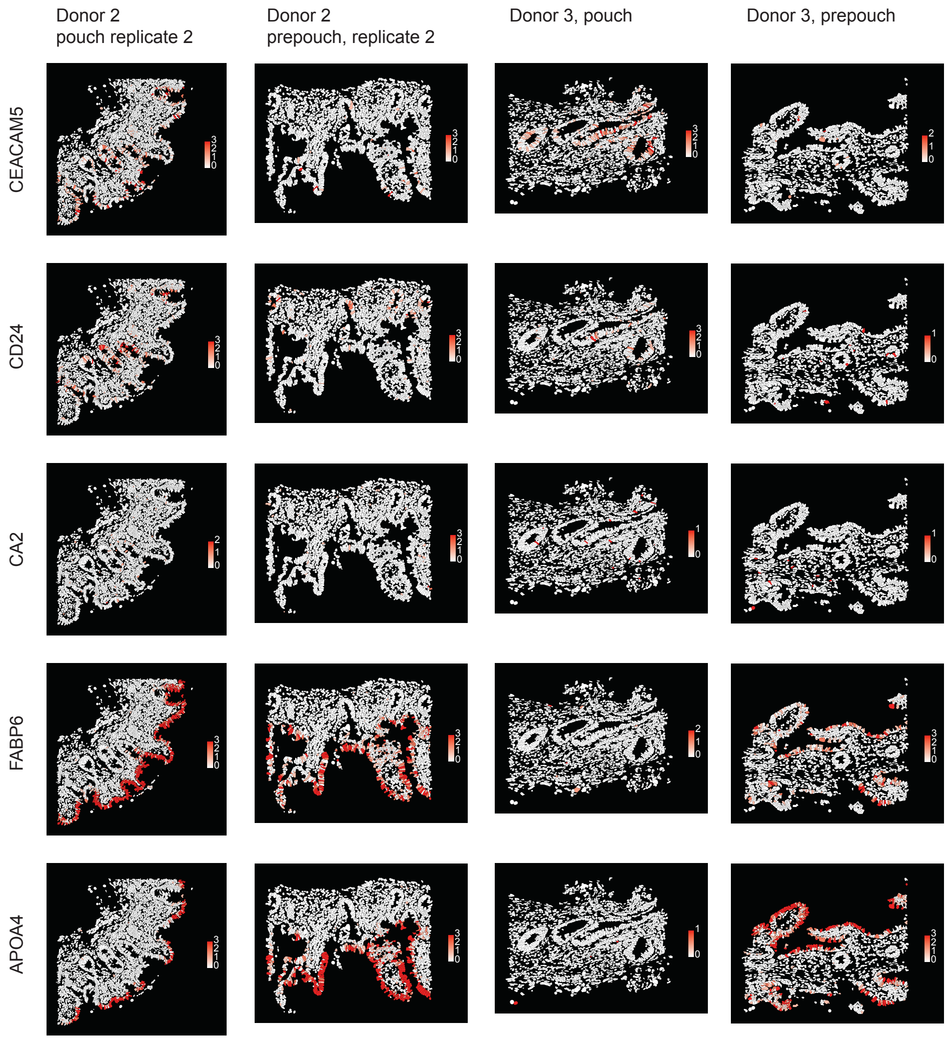

### Supplementary Data 8

Supplementary Data 8-1

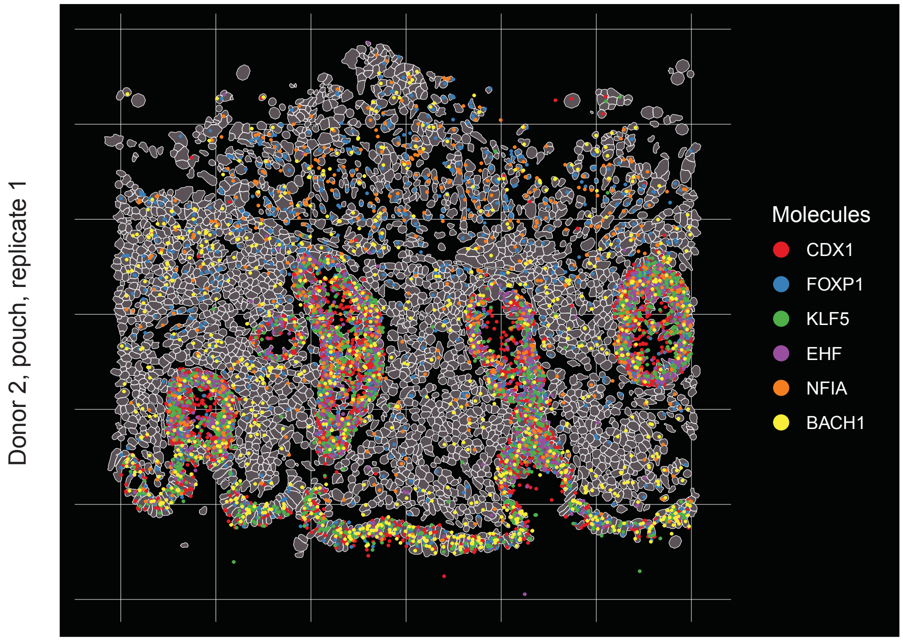


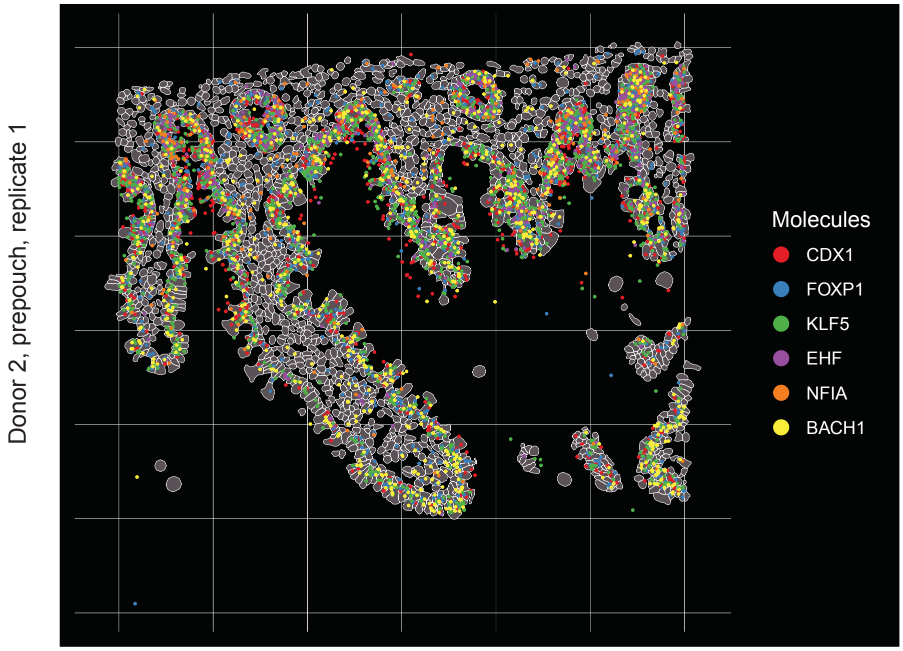

Supplementary Data 8-2

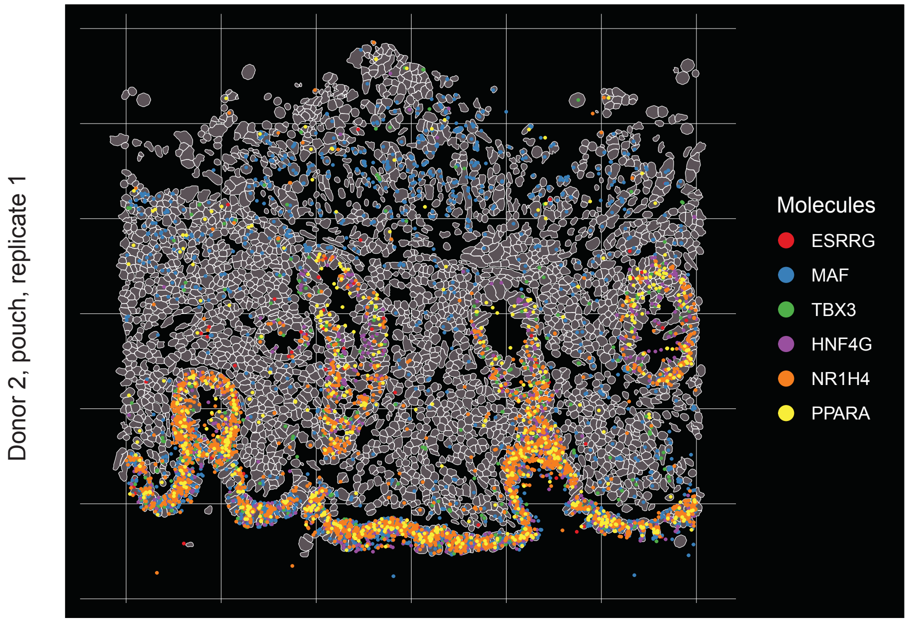


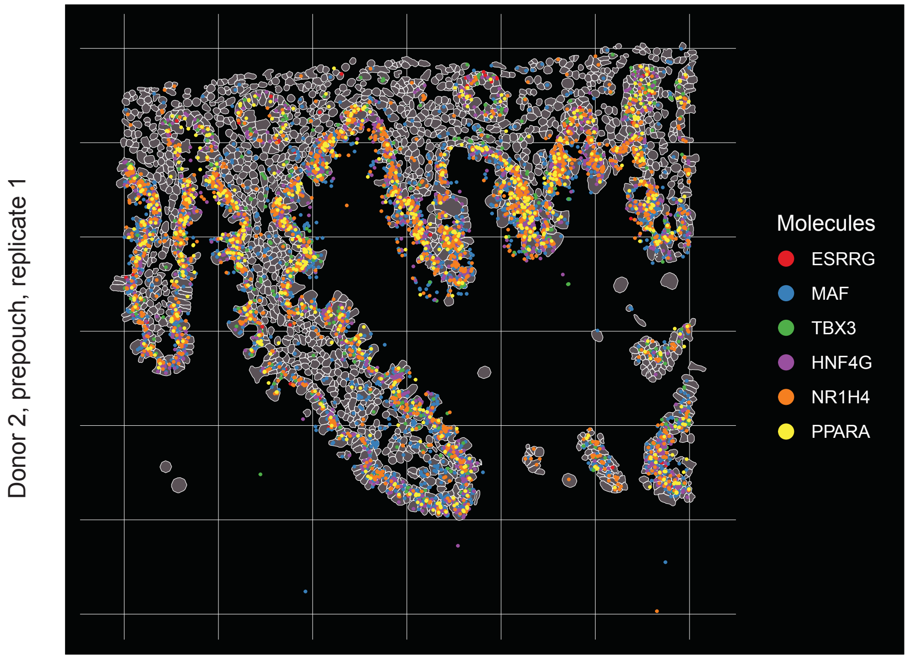

### Supplementary Data 9

Supplementary Data 9-1

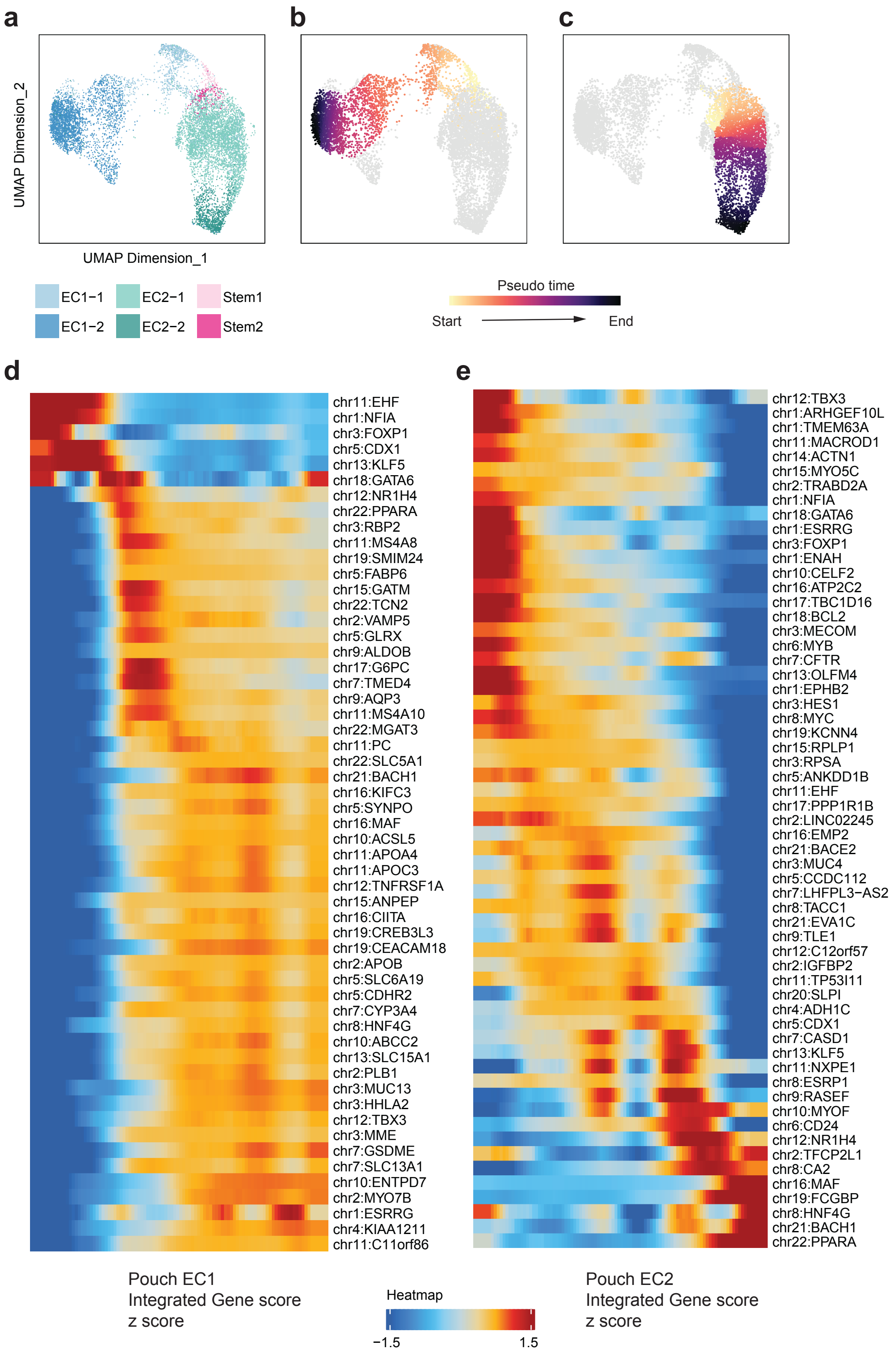

Supplementary Data 9-2

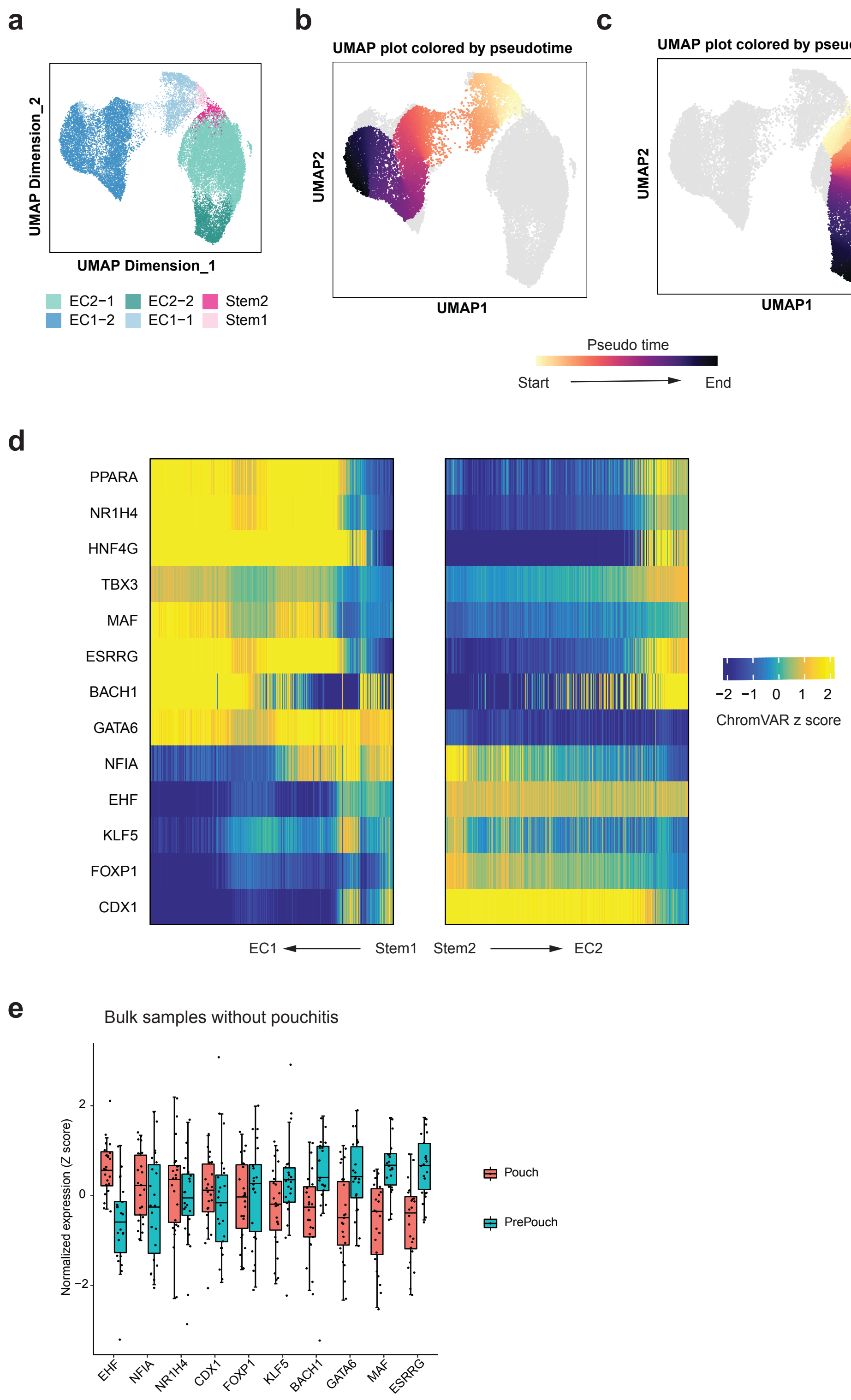
